# Noise is the signal: variability in surveillance data provide early indication of epidemic phase

**DOI:** 10.64898/2026.07.24.26358802

**Authors:** E. Joe Moran, Dustin T. Hill, Mohammed A. Alazawi, David A. Larsen

**Affiliations:** Department of Public Health, Syracuse University, Syracuse, NY, USA; Center for Environmental Health, New York State Department of Health, Albany, NY, USA

**Author notes:** Correspondence should be sent to David Larsen, 444F White Hall, Syracuse University, Syracuse, NY 13244.

## Abstract

Predicting epidemic surges is challenging and often relies on prior seasons, outbreaks in other regions, and advanced modeling of surveillance data. Using foundational principles of disease growth and spread, including trend analysis and overdispersion, we developed a surge detection algorithm for emerging pathogens that requires no historical baselines or seasonal assumptions. We used it to evaluate temporal variability (coefficient of variation, CV) and spatial clustering (global Moran’s I) as early warning indicators across three parallel SARS-CoV-2 surveillance streams: wastewater, clinical cases, and hospitalizations. All metrics showed consistent variability patterns across epidemic phases that predicted surge onset and decline approximately one month in advance. For weekly pre-surge detection, sensitivity and positive predictive value reached 75% and 54%. CV additionally signaled surge termination, with up to 70% sensitivity and 59% positive predictive value. When evaluated per surge, by requiring only a single correct signal within each phase, CV detected 86–98% of onsets and 73–85% of terminations. These signatures held across surveillance streams and geographic scales, suggesting they reflect system-level transmission dynamics rather than stream-specific artifacts, consistent with overdispersed transmission producing high variability before and after peak. Requiring only basic quantitative methods, our framework is implementable by practitioners without specialized training in dynamical systems.

## Introduction

Epidemic surges of infectious diseases begin with a small number of cases that are hard to distinguish from baseline transmission, yet by the time a surge is identifiable through conventional surveillance the window for early intervention has often closed. Early in an epidemic curve, disease occurrence is stochastic and driven by importation and spatial connectivity.^1^ As transmission increases it becomes more deterministic and spatially diffuse.^2^ Because all infectious diseases exhibit overdispersion, clustering in both space and time occurs. This clustering is most apparent during the stochastic phases that bookend a surge — the establishment phase preceding onset and the burnout phase following termination — and is least apparent at peak transmission when disease is widespread. Changes in both spatial and temporal variability may therefore signal impending surge transitions in both directions (Figure 1).

**Figure 1:**
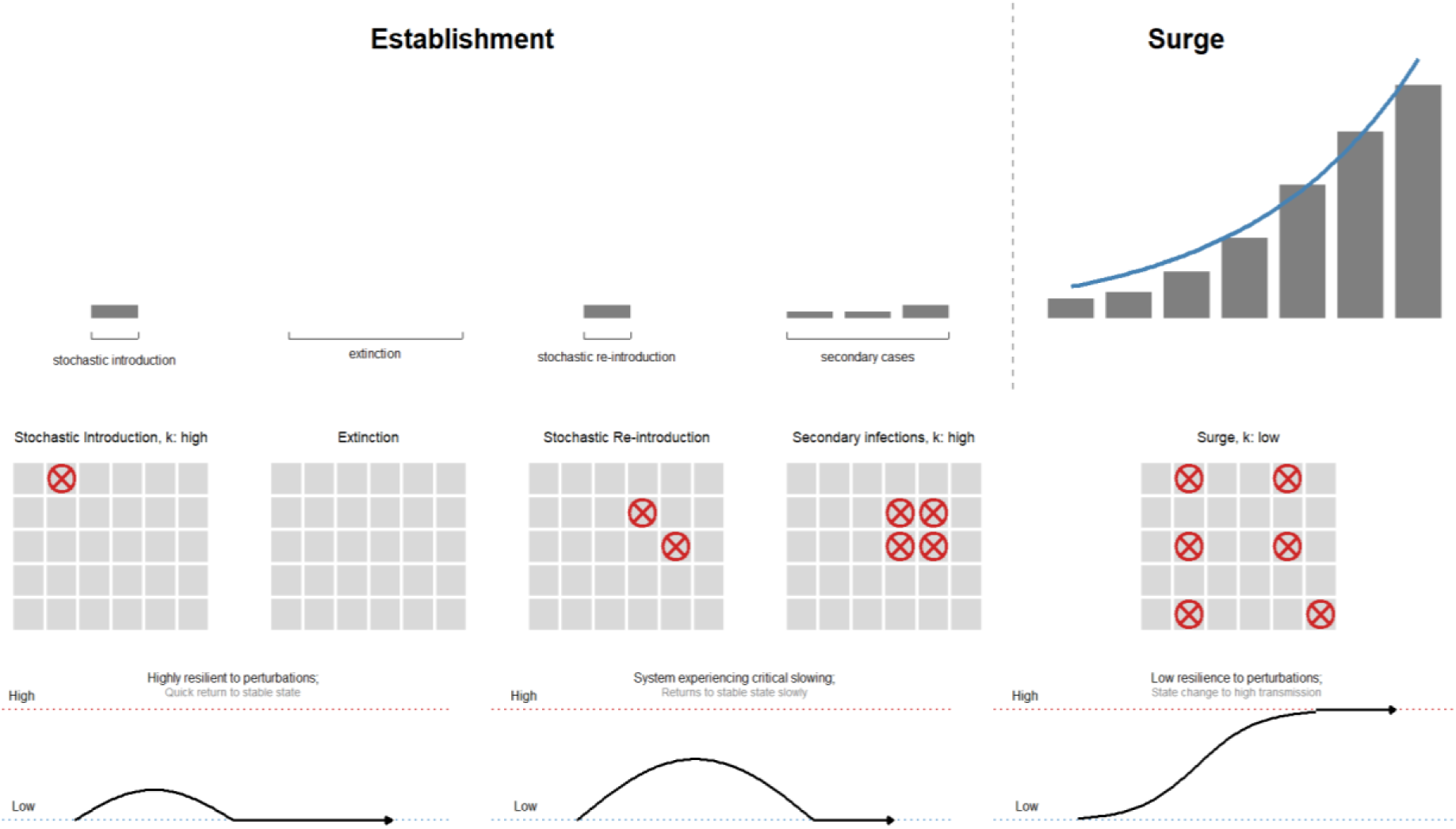
Epidemic curve and its relationship to stochasticity and overdispersion. Prior to a surge, disease incidence is stochastic, with high variance and potential for die out or establishment (establishment phase). Secondary cases are highly clustered and slow the return to a stable state (low transmission). When a surge is established, the new infections are prevalent enough that they perturb the system into a new high transmission state, where disease incidence is common and well dispersed throughout the susceptible population.

Early warning of infectious disease surges can manifest in several ways: an outcome that precedes another (e.g., a COVID case preceding hospitalization), a disease driver such as weather patterns for mosquito-borne diseases,^3^ or system characteristics that change prior to a state transition. The latter can be described through resilience indicators related to critical slowing down (CSD) theory at bifurcation points. As a dynamical system approaches a bifurcation point, it becomes less resilient to perturbations and returns to baseline more slowly.^4^ These dynamics can be observed as increasing variance and autocorrelation in a time series of measurements,^5,6^ where larger perturbations are less likely to self-correct and drive the system toward a new state. Such transitions have been documented in eutrophication,^7^ climate change,^8,9^ ecological systems,^7,10^ economics,^11^ and theoretical epidemiology.^5,6^ In applied epidemiology, we theorize this state change represents the transition from low community transmission of an infectious disease to a high-transmission surge.

Existing epidemic or surge detection methods rely on historical data, using temporal composites anchored to specific points in time. These approaches compare current observations to the same time period across multiple previous years^12^ and essentially require two parameters: the depth of historical data to be used, and the seasonal window for identifying comparable time periods. From historical baselines, normal ranges and surge thresholds can be calculated. Unfortunately, this approach does not work for emerging pathogens or pathogens that do not exhibit predictable seasonal patterns but are instead driven by other mechanisms. COVID-19 fell into this category for several reasons: (1) it was an emerging pathogen with no historical baseline;^13^ (2) social and political actions significantly influenced its spread in ways that varied unpredictably over time;^14^ (3) population immunity was initially absent and then shifted dramatically with the introduction of vaccines and infection-acquired immunity;^15^ and (4) transmission dynamics were initially driven by variant emergence rather than seasonal influences.^16^ More recent threshold-based approaches, such as the moving epidemic method (MEM), use sliding windows of historical data to establish epidemic thresholds and intensity levels, and have been applied to influenza surveillance across diverse settings.^17,18^ However, MEM still requires prior seasonal data to characterize baseline activity, and its thresholds are calibrated to magnitude alone, without capturing the directionality of change or accounting for heterogeneous noise characteristics across data streams. We developed an alternative algorithm for identifying surges that does not rely on baseline historical data and can be applied to emerging or re-emerging pathogens.

Herein we show variability as an observable characteristic before and after surges in our study of SARS-CoV-2 transmission using three separate infectious disease surveillance data streams: wastewater, clinical cases, and hospitalizations. We test whether temporal variability in epidemiological data and spatial clustering in case data can reliably identify significant pre- and post-surge transitions. Rather than asking ‘how many cases or hospitalizations are there?’, we ask ‘how is the system behaving?’. We hypothesize that the variance-based metrics coefficient of variation for temporal data and Moran’s I for spatial data derived from infectious disease surveillance data streams can provide earlier and more accurate warnings than traditional approaches. These metrics capture the system-level epidemiological characteristics and critical slowing down dynamics that surround epidemic transitions.

## Results

### Surge Detection Algorithm Development

We developed a surge detection algorithm that uses empirical cumulative distribution functions (ECDFs) of both magnitude and slope calculated from rolling windows of recent data, allowing real-time adaptation to shifting transmission dynamics and includes five steps (Figure 2). Through these five steps, we progressively incorporated slope-based detection criteria, 26-week rolling window ECDFs, minimum thresholds to prevent false positives during low-circulation periods, and temporal filtering to consolidate meaningful epidemic waves. The inclusion of slope as a detection criterion adds directional sensitivity — distinguishing accelerating transmission from sustained but stable activity — while minimum threshold and temporal filtering stages suppress false positives arising from noisy or low-signal data streams, a limitation that magnitude-only approaches do not explicitly address. By anchoring detection to recent data rather than historical baselines, this approach is suitable for emerging and re-emerging pathogens, novel variants, and contexts where historical patterns are unreliable or nonexistent.

**Figure 2.**
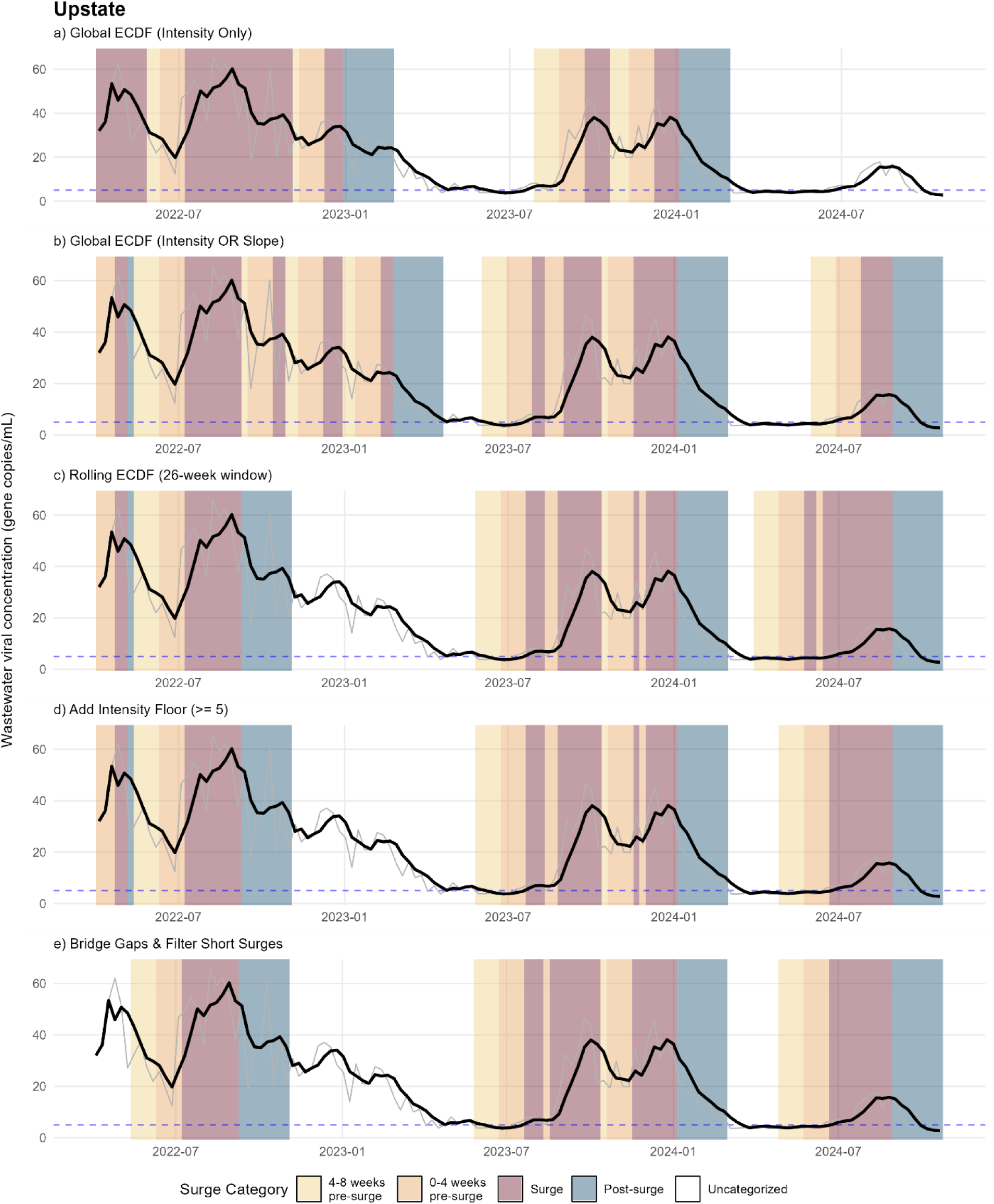
Progressive development of the surge detection algorithm showing five iterative refinement stages for Upstate New York. Each panel displays wastewater SARS-CoV-2 concentration (gene copies/mL) over time with background shading indicating classified epidemic phases, as seen in the legend. Stage 1 **(a)** only used global ECDF of intensity to classify surges above the 75^th^ percentile. Stage 2 **(b)** added ECDF slope criteria of 75% to capture rapid increases. Stage 3 **(c)** replaced global ECDFs calculated on all data points with 26-week rolling windows to adapt to evolving transmission dynamics. Stage 4 **(d)** incorporated a minimum intensity threshold (5 gene copies/ml) to eliminate false positives during low-circulation periods. Stage 5 **(e)** applied temporal filtering to bridge gaps shorter than two weeks and remove periods classified as a surge lasting less than three weeks. This method progression can be viewed for clinical cases and hospitalizations in Supplemental Figure S1-S2.

Surge category classifications demonstrated moderate to substantial agreement across the different surveillance streams using the kappa statistic, with the strongest agreement observed between lagged clinical cases and hospitalizations (statewide average κ = 0.640; highest regional average κ = 0.737, Supplemental Table 1). To evaluate category-specific agreement, observations were pooled across all regions and dichotomized for each surge phase (target category vs. all others), and kappa was calculated from the resulting 2×2 confusion matrix. Dichotomous one-vs-rest kappa analysis revealed that agreement was highest in the surge and post surge categories, agreement of κ = 0.710 and κ = 0.786, respectively, supporting the validity of our surge detection algorithm for identifying the critical transition points marking surge onset and termination for multiple data streams.

### Temporal Variability: Analysis of Coefficient of Variation Across Epidemic Phases

Analysis of the coefficient of variation (CV) across epidemic phases revealed characteristic variability patterns that distinguish both the onset and termination of SARS-CoV-2 surges (Figures 3–5). Across wastewater (Figure 3), clinical cases (Figure 4), and hospitalizations (Figure 5), the pre-surge periods exhibited elevated CV compared to uncategorized and surge periods, consistent with the theoretical expectation that disease incidence exhibits greater stochasticity during early epidemic growth phases. During active surges, CV declined and compressed to a narrow range, reflecting the transition from stochastic to sustained transmission. CV then rose again during the post-surge fadeout as incidence shifted from sustained to sporadic, before settling to baseline levels. The boxplot distributions (Figures 3c–5c) demonstrate clear separation between these epidemic phases, with the pre-surge and post-surge periods bookending a distinct low-variability surge period.

**Figure 3.**
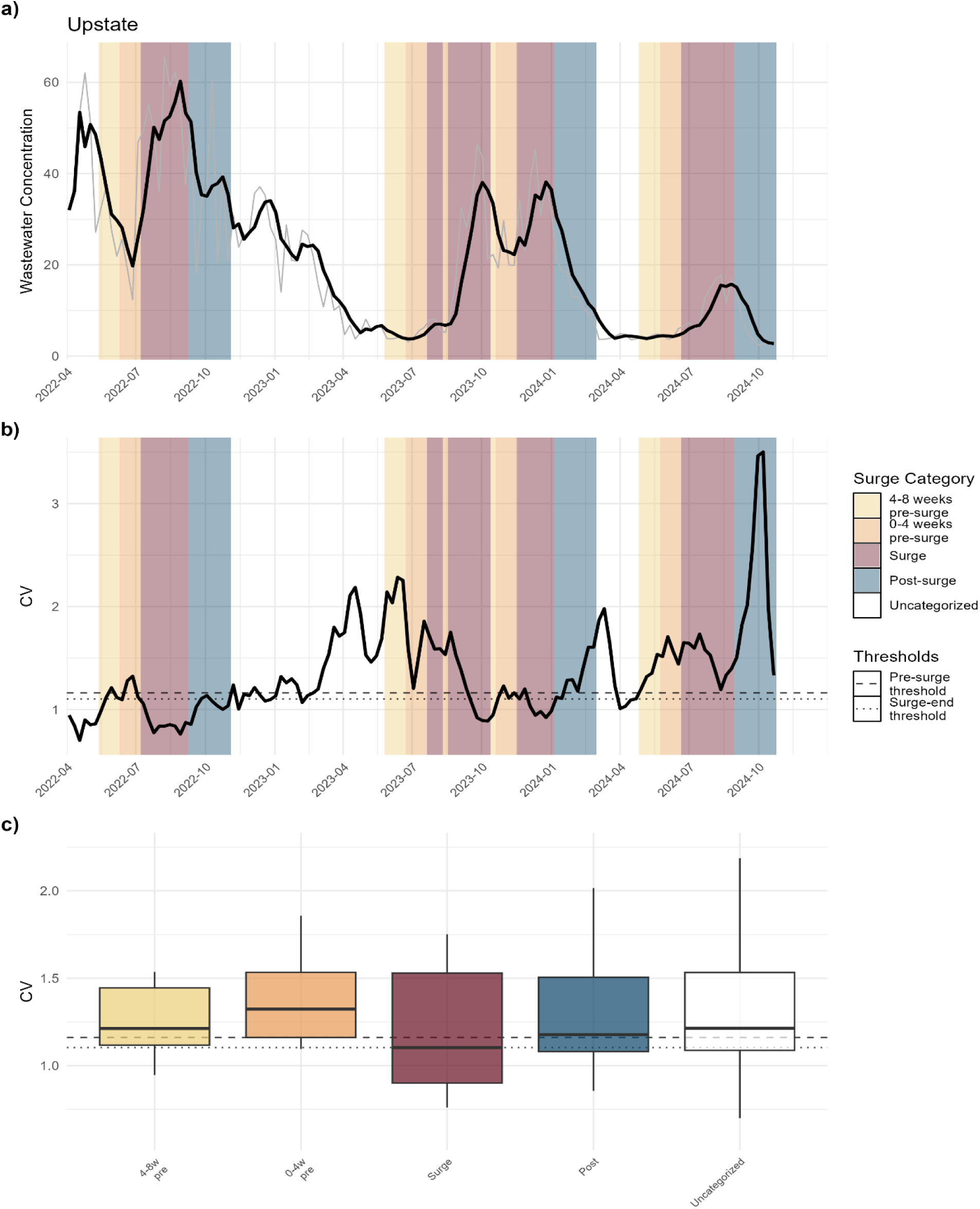
Coefficient of variation analysis for wastewater concentration in Upstate New York. Panels show: **a)** wastewater concentration time series raw values (grey) with 3-week smoothing (black), **b)** coefficient of variation over time with 25th percentile threshold of pre-surge periods (dashed) and 50th percentile threshold of surge periods (dotted), and **c)** distribution of CV values by surge category and threshold lines. Background colors indicate epidemic phases, as shown in the legend. Kruskal-Wallis tests indicated significant differences in CV amongst sequential surge categories (p < 0.001); all pairwise comparisons are provided in Supplemental Tables S3-S5.

**Figure 4.**
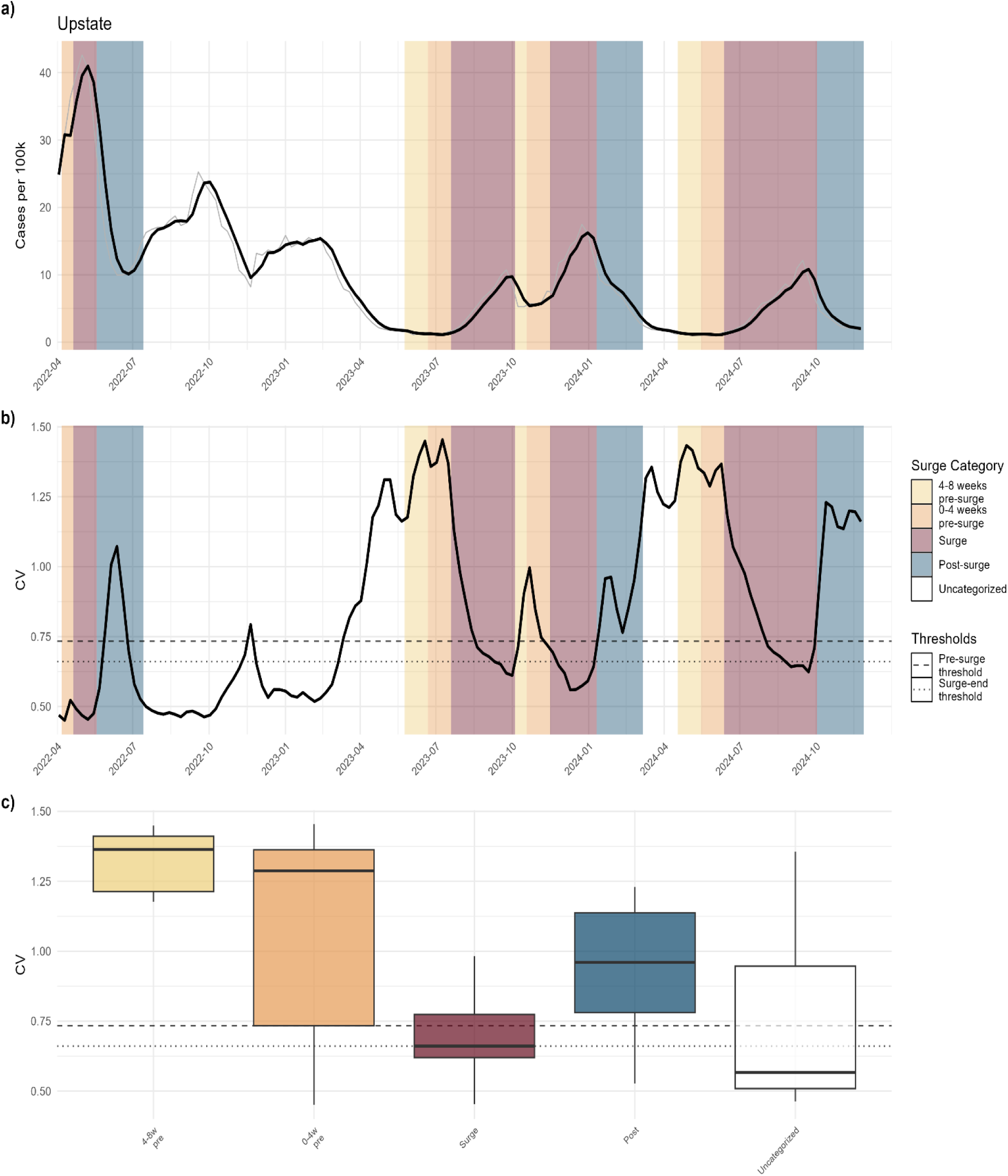
Coefficient of variation analysis for clinical COVID-1G incidence in Upstate New York. Panel structure and color scheme as in Figure 3. Incidence is expressed per 100,000 population. Kruskal-Wallis tests indicated significant differences in CV amongst sequential surge categories (p < 0.001); all pairwise comparisons are provided in Supplemental Tables S6-S8.

**Figure 5.**
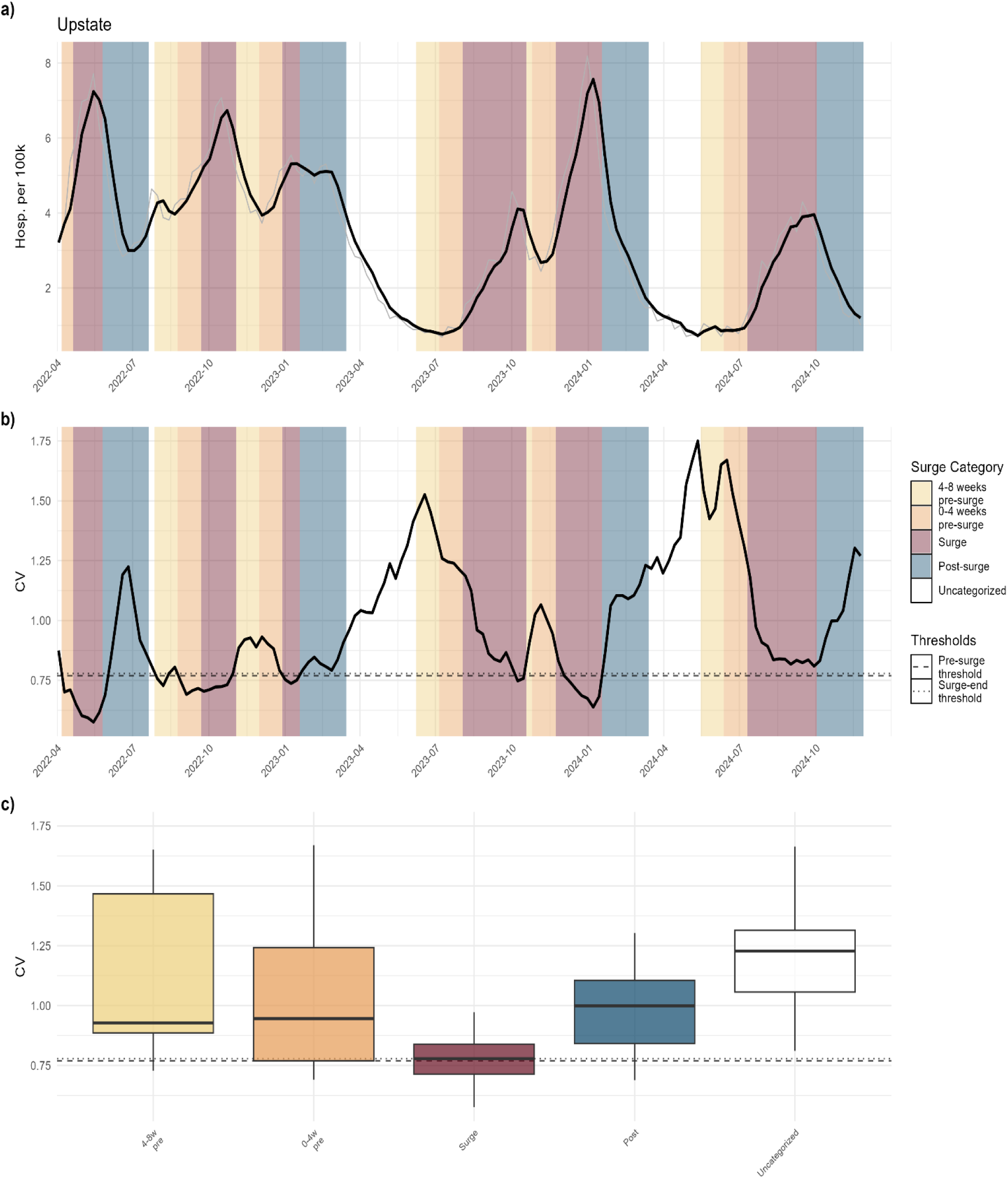
Coefficient of variation analysis for COVID-1G hospitalization incidence in Upstate New York. Panel structure and color scheme as in Figure 3. Hospitalization incidence is expressed per 100,000 population. Kruskal-Wallis tests indicated significant differences in CV amongst sequential surge categories (p < 0.001); all pairwise comparisons are provided in Supplemental Tables S9-S11.

Upstate NY (all NYS economic development regions except Mid-Hudson, New York City and Long Island), Downstate NY (Mid-Hudson, New York City and Long Island regions), and individual economic development regions exhibited distinct early warning signal characteristics. The Upstate region generally showed higher observed CV values and more pronounced elevations during pre- and post-surge periods, likely reflecting greater county-level heterogeneity in disease transmission dynamics (Figure 3). The Downstate region displayed lower and more stable CV values but maintained consistent pre- and post-surge elevation patterns relative to its regional baseline (see Supplemental Figures S3, S14, S27), likely reflecting the more densely connected New York City metropolitan area. Despite these differences in observed magnitude, all regional aggregations demonstrated the same CV patterns that form the basis for early warning signals throughout the phases of a surge.

### Spatial Variability of Clinical Cases

County-level spatial autocorrelation analysis revealed distinct patterns of geographic clustering throughout the study period (Figure 6). Global Moran’s I values showed elevated clustering during pre-surge periods, with the 4–8 and 0–4 week pre-surge windows exhibiting higher values compared to uncategorized and post-surge periods (Table S12). This pattern was consistent across all major epidemic waves, with global Moran’s I typically rising 4–8 weeks before surge onset, peaking in the immediate pre-surge period, and declining during surge phases as transmission became more geographically dispersed. Unlike CV, which exhibited a transient rise during post-surge fadeout, global Moran’s I continued to decline through the surge and post-surge periods, consistent with the expectation that transmission remains spatially dispersed during fadeout — cases decline in frequency but remain geographically dispersed, rather than re-clustering. The boxplot distributions (Figure 6, panel C) demonstrate a clear pre-surge peak followed by a continual decline. Notably, the spatial signal appeared to emerge earlier than the temporal variability captured by CV, with Moran’s I often rising in the 4–8 week window preceding surge onset, suggesting that spatial clustering may provide the earliest detectable signature of impending surges, as examined further in the performance evaluation below. Because Moran’s I is calculated at the statewide level using county-level case data (n=62 counties), it represents a single statewide metric per time point rather than the region-specific estimates generated for CV.

**Figure 6.**
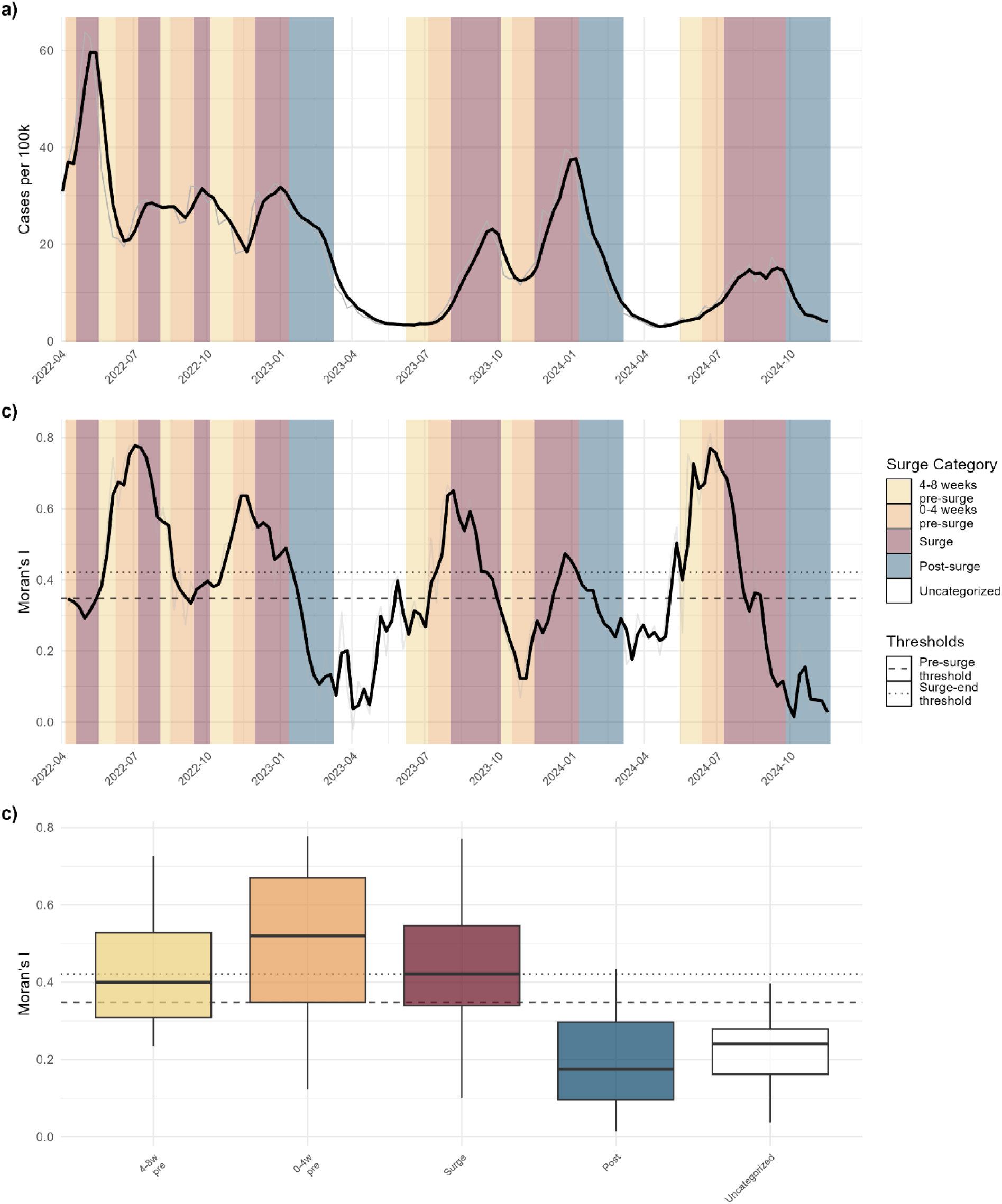
COVID-19 spatial analysis of New York State showing county-level case rates and global Moran’s I spatial autocorrelation patterns. **a)** Statewide COVID-19 case rates calculated as population weighted county-level aggregation showing epidemic waves from April 2022 through December 2024. **b)** County-level spatial autocorrelation (global Moran’s I) time series using k=4 nearest neighbors (n=62 counties), with three week rolling mean (black line) and raw values (gray line); 25th percentile threshold of pre-surge periods (dashed) and 50th percentile threshold of surge periods (dotted), and **c)** Distribution of global Moran’s I values by surge category showing elevated clustering during pre-surge periods. Background colors indicate epidemic phases, as shown in the legend Kruskal-Wallis tests indicated differences in CV amongst sequential surge categories; all pairwise comparisons are provided in Supplemental Table S12.

### Performance of Early Warning Indicators

Having established that CV and global Moran’s I exhibit characteristic patterns across epidemic phases, we evaluated the performance of alert thresholds for pre-surge and post-surge periods. For pre-surge detection we used the 25th percentile of the 0–4 weeks pre-surge category of each metric as an alarm threshold. CV pre-surge thresholds identified periods of elevated variability characteristic of stochastic introductions and overdispersed transmission events, while global Moran’s I thresholds captured periods of strong spatial clustering indicative of localized transmission before widespread diffusion. For post-surge detection, the 50th percentile of the surge category served as the CV threshold for identifying periods of low variability characteristic of uniform transmission during active surges, signaling that the surge is likely nearing termination.

For pre-surge detection, both temporal and spatial variability measures demonstrated comparable sensitivity of approximately 75% (Table 1). This consistency is expected given the threshold-setting approach: by defining the alarm threshold as the 25th percentile of values observed during 0–4 week pre-surge periods, approximately 75% of true pre-surge periods exceeded the threshold by design. Specificity was high across all methods (72–86%), indicating that most non-pre-surge periods did not trigger false alarms. The positive predictive value (PPV) ranged from 34–54%, and the negative predictive value (NPV) exceeded 93% for all methods, indicating high confidence that periods not triggering an alarm are truly not pre-surge. For post-surge detection, CV-based metrics achieved sensitivity of 55-70%, specificity of 68-81%, PPV of 42-59%, and NPV of 80-87% (Table 1). Global Moran’s I performed less well for post-surge detection (54% sensitivity, 41% specificity, 29% PPV, 67% NPV).

**Table 1:**
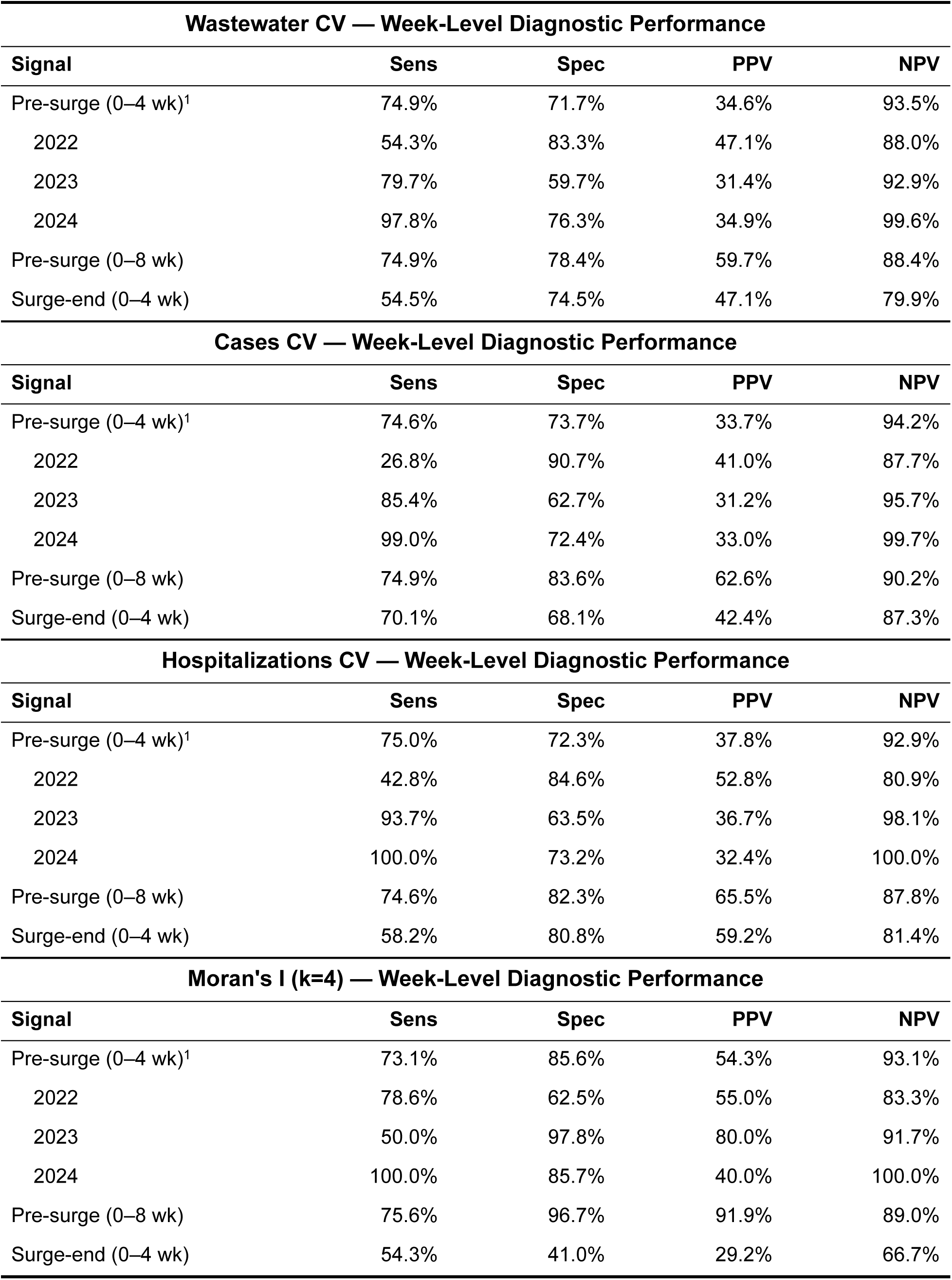
Week-level diagnostic performance of variability metrics as early warning indicators for COVID-1G surge detection in New York State. For each metric, sensitivity, specificity, positive predictive value (PPV), and negative predictive value (NPV) are shown for the 0–4 week pre-surge signal, where condition positive is defined as the metric exceeding the 25th percentile of the pre-surge distribution. Year-specific subsets (2022–2024) reflect performance across distinct pandemic eras and are nested within the overall 0–4 week pre-surge signal. An extended 0–8 week pre-surge window is also evaluated, where condition positive extends to the full 8 weeks preceding surge onset. For the 0–4 week surge-end signal, condition positive is defined as the metric falling below the 50th percentile of the surge distribution, with surge termination occurring within the following four weeks. Results are shown for coefficient of variation (CV) calculated from wastewater concentration, clinical case rates, and hospitalization rates, and for global Moran’s I spatial autocorrelation of clinical cases. All results represent average performance across New York State regions.

**Table 2.** Event-level detection performance of variability metrics as early warning indicators for COVID-1G surge detection in New York State. For each metric and signal window, event-level detection summarizes the proportion of surge events for which the signal fired at least once during the relevant window (0–4 weeks pre-surge or 0–4 weeks prior to surge end). Results are shown for coefficient of variation (CV) calculated from wastewater concentration, clinical case rates, and hospitalization rates, and for global Moran’s I spatial autocorrelation of clinical cases (k = 4 nearest neighbors). Detection rate, total surge events evaluated, and counts of detected and missed events are reported. All results represent average performance across New York State regions.

| **Wastewater CV — Event-Level Detection<sup>2</sup>** |  |  |  |  |
| --- | --- | --- | --- | --- |
| **Signal** | **Detection Rate** | **Total Surge Events** | **Detected** | **Missed** |
| Pre-surge (0–4 wk) | 98.1% | 56 | 55 | 1 |
| Surge-end (0–4 wk) | 73.2% | 56 | 41 | 15 |
| **Cases CV — Event-Level Detection<sup>2</sup>** |  |  |  |  |
| **Signal** | **Detection Rate** | **Total Surge Events** | **Detected** | **Missed** |
| Pre-surge (0–4 wk) | 85.7% | 56 | 48 | 8 |
| Surge-end (0–4 wk) | 84.9% | 56 | 47 | 9 |
| **Hospitalizations CV — Event-Level Detection<sup>2</sup>** |  |  |  |  |
| **Signal** | **Detection Rate** | **Total Surge Events** | **Detected** | **Missed** |
| Pre-surge (0–4 wk) | 92.1% | 67 | 62 | 5 |
| Surge-end (0–4 wk) | 78.2% | 67 | 52 | 15 |
| **Moran's I (k=4) — Event-Level Detection<sup>2</sup>** |  |  |  |  |
| **Signal** | **Detection Rate** | **Total Surge Events** | **Detected** | **Missed** |
| Pre-surge (0–4 wk) | 71.4% | 7 | 5 | 2 |
| Surge-end (0–4 wk) | 71.4% | 7 | 5 | 2 |

The temporal pattern of CV relative to surges is informative across the full epidemic curve. CV rises and peaks prior to a surge, then drops off as the surge begins — providing the critical early warning signal. Even in instances where the CV threshold is not crossed in the context of the performance test, the pattern of a rising peak followed by a decline into the surge is remarkably consistent. During the peak of a surge, CV declines and compresses to a narrow range, and this drop signals the end of the surge with a PPV of 42-59%, with roughly 73-85% of all surges exhibiting this signal at least once in the four weeks preceding surge termination. Careful investigation of surges that do not meet the criteria for performance testing reveals that the pattern of declining CV is still observed — it simply does not cross the threshold. As the surge ends, CV rises again as sporadic fadeout produces a transient increase in CV before settling until the next surge approaches. Spatial clustering as measured by global Moran’s I showed a more consistently elevated 0– 4 week pre-surge PPV of 54%, and week-by-week values (Supplemental Figure S38) indicate that spatial clustering is often detectable even earlier than the 0–4 week window observed with temporal variability. When both pre-surge categories are considered condition-positive above the threshold, PPV rises to 92%. Unlike CV, however, global Moran’s I does not provide a clear phase transition for post-surge detection. Once global Moran’s I peaks, it monotonically declines through the surge and post-surge period (Figure 6). This is consistent with our conceptual model of infectious disease dynamics: at the beginning of an outbreak, disease incidence is highly overdispersed and spatially clustered; during the surge, transmission becomes more evenly dispersed throughout the population; and during fadeout, remaining infections are still spatially uniform but more variable in frequency. Although spatial fadeout is evenly dispersed, CV can still detect the shift from sustained to sporadic incidence, which is why CV provides bidirectional surge detection while global Moran’s I is primarily a pre-surge indicator.

We observed differences in performance metrics across years. The years 2022 and 2024 generally performed better than 2023, which we attribute to the more sharply defined outbreaks in those years. Our data show that 2023 experienced a prolonged post-surge decline characterized by multiple minor resurgences during a period of high variant diversity, which we believe contributed to the drawn-out and irregular decline pattern. Because CV values carry directional information relative to surge timing, slope criteria can be applied alongside CV thresholds to suppress false alarms that arise when transient spikes in CV occur during prolonged decline periods rather than during genuine pre-surge acceleration. Without slope gating, a brief increase in variability during a prolonged fadeout — such as the pattern observed in 2023 — could trigger a pre-surge alarm even though the broader trajectory is downward. Requiring positive slope over a defined window ensures that alarms correspond to sustained increases in variability rather than isolated fluctuations. Using this, gains in PPV can be achieved at the expense of sensitivity; however, the magnitude of this tradeoff depends on the slope threshold selected, which is inherently system-specific. We do not present fixed performance results for slope mediated signals, as the optimal balance between false alarm suppression and sensitivity will vary by operational context. Rather, we present this conceptually as a tunable parameter that practitioners can calibrate to their specific decision-making requirements.

### Temporal Resolution

We used a 6-month rolling window to compute ECDFs, preceded by a global ECDF calculated from all available data, which was used for thresholds during the first six months of the study period. The global ECDF ensured that the first surge was assessed against an adequately large reference distribution; had we parameterized with only the initial months of data, the algorithm would have been calibrating the first surge against itself. Once sufficient data accumulated, the 6-month rolling window proved adequate for several reasons. First, six months was long enough that the window always contained a complete epidemic cycle, preventing skewed percentile distributions reflective of only one epidemic phase that would be hypersensitive to any change. Second, six months was dynamic enough to keep pace with the shifting transmission landscape driven by new variants, changing social behavior, and evolving population immunity. These characteristics are disease-specific and should be considered when adapting our methods to new pathogens and surveillance systems.

To define temporal categories for analyzing surge dynamics, we examined patterns of CV and global Moran’s I grouped by week relative to surge onset (Supplemental Figures S3, S14, S26). CV generally peaked 3–4 weeks before a surge, with this signal tending to be less pronounced and slightly earlier in the downstate region. Global Moran’s I also tended to peak slightly earlier than CV (Supplemental Figure S38). These patterns qualitatively informed our selection of 0–4 week and 4–8 week pre-surge categories, as well as an up to 8-week post-surge category. Most economic development regions showed CV peaking in the 0–4 week pre-surge window, but notable increases in CV were also observed in the 4–8 week window preceding the peak, supporting the inclusion of both categories. These groupings conveniently align with monthly increments, enabling straightforward operational language: “CV has crossed our alert threshold — a surge is likely within the next month,” or “CV has dropped below our threshold — the surge is likely to end within the next month.”

### Spatial Resolution

Our findings demonstrate robustness across multiple geographic scales of aggregation. For wastewater surveillance data, we conducted analyses at both the economic development region level and a broader upstate-downstate classification, with results remaining consistent across these spatial frameworks. County-level analysis was not feasible for wastewater data because most counties contain only one to three wastewater treatment plants sampling once or twice weekly, yielding insufficient sample sizes to reliably distinguish systematic noise patterns from stochastic variation and sampling error. Conversely, statewide aggregation introduced substantial heterogeneity in laboratory methodologies, imparting persistent analytical noise that obscured the temporal signals of interest; regional groupings largely mitigated this. Clinical case counts and hospitalization data were available at the county level. Similarly, this required regional aggregation given that a single observation, per county, per time period prevents reliable noise characterization. These constraints illustrate the importance of selecting an appropriate spatial scale for noise-based surge detection. Aggregation units that are too small lack sufficient data density to differentiate meaningful fluctuations from measurement error and true randomness, while excessively large units smooth the localized noise signatures seen during phases of epidemic waves.

Spatial autocorrelation analysis was restricted to clinical case data due to limitations in applying this framework to other metrics. Wastewater treatment plants represent discontinuous sampling units that rarely share geographic boundaries, effectively functioning as spatial islands that violates the continuity assumption of the global Moran’s I statistics. Additionally, the population served by each wastewater treatment plant varies substantially, and the proportion of county populations captured by wastewater surveillance differs between urban and rural areas, introducing spatial heterogeneity amongst the baseline data. The community-scale averaging inherent in wastewater sampling may also attenuate individual-level spatial clustering by aggregating a signal across a sewershed. Hospitalization data posed different constraints. Variant-associated severity differs and widely varying county demographics confound spatial patterns. Five NYS counties lack hospitals entirely, requiring patient migration to neighboring counties and introducing spatial misrepresentation. Additionally, declining hospitalization rates throughout the pandemic have made COVID-19 hospitalizations an increasingly rare event, making it challenging to distinguish true spatial clustering from random variation.

**Figure 7.**
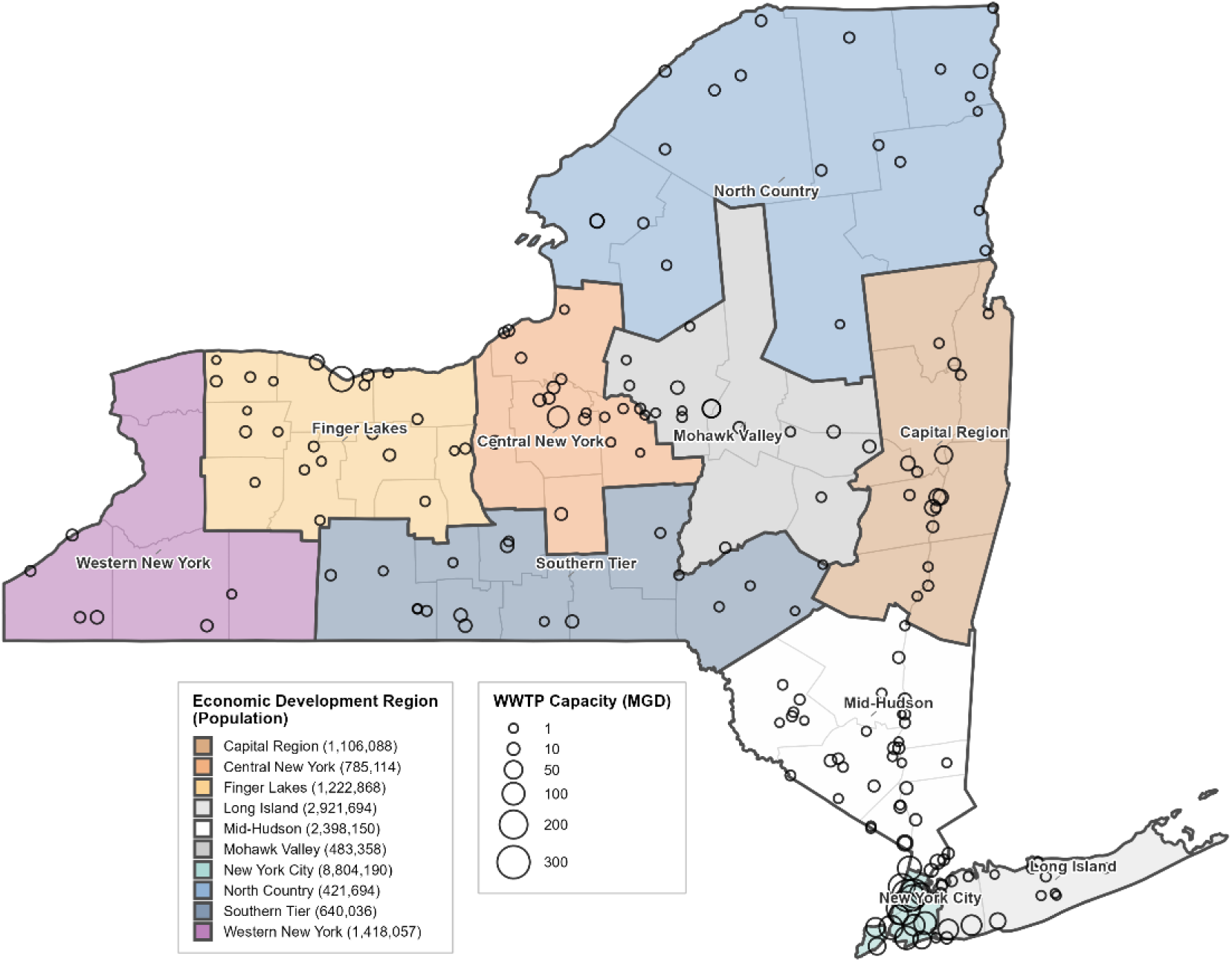
Distribution of wastewater treatment plants (WWTPs) across New York State economic development regions participating in the NYS wastewater surveillance network. Hollow circles represent individual WWTPs, scaled by treatment capacity (million gallons per day, MGD). Shaded regions are colored by economic development region with regional populations shown in parentheses. County boundaries are displayed in light grey.

## Discussion

Our findings demonstrate that variability in epidemiological surveillance data serves as a reliable indicator of epidemic phase, providing approximately one month of lead time before both surge onset and termination. We developed a novel surge detection algorithm capable of identifying epidemic waves without relying on historical baselines, a critical feature for emerging pathogens and diseases with unpredictable transmission dynamics. Using this framework, we showed that both temporal variability (coefficient of variation) and spatial clustering (global Moran’s I) exhibit characteristic patterns across epidemic phases: elevated variability during the weeks preceding a surge, declining and compressed variability during active surges, and a transient rise during post-surge fadeout. The consistency of these patterns across fundamentally different metrics — temporal variability in three surveillance streams and spatial autocorrelation of case data — suggests that the observed variability signatures reflect robust system-level dynamics rather than artifacts of any particular data source.

Among the metrics evaluated, global Moran’s I performed best for pre-surge detection, achieving 73% sensitivity, 86% specificity, 54% PPV, and 93% NPV. Widening the pre-surge window to 0–8 weeks improved global Moran’s I performance substantially, with specificity and PPV rising to 97% and 92%, respectively. CV-based metrics also performed adequately for pre-surge detection and were very consistent across surveillance streams. Surge termination was best predicted by CV, with similar performance across surveillance streams apart from hospitalizations, which had a notably higher PPV of 60%. Wastewater CV makes a particularly compelling metric for predicting surge termination, as wastewater concentration itself leads cases and hospitalizations temporally for SARS-CoV-2. Although global Moran’s I performed best for predicting surge onset, it performed worst for predicting surge termination, with a PPV of only 29%, consistent with the monotonic decline of spatial autocorrelation once transmission becomes geographically uniform. NPV was generally high across all metrics and tests at 80–94%, indicating high confidence in non-surge results. At the event level, CV detected 86–98% of surge onsets and 73–85% of surge terminations. Global Moran’s I event-level detection was lower at 71% for both surge onset and termination, though it should be noted that global Moran’s I is a single statewide measure with a considerably smaller sample size, correctly detecting 5 of 7 surge onsets and 5 of 7 surge terminations.

### Surge Detection Method

Central to these findings is the novel surge detection algorithm that we implemented, which allows for identifying epidemic waves without relying on historical baselines or equilibrium assumptions. Threshold-based approaches using fixed seasonal baselines, such as the Cullen and CDC c-sum methods, require multiple years of stable historical data and assume seasonal stationarity,^19^ conditions rarely met during pathogen emergence or epidemic re-emergence. Outbreak detection algorithms based on generalized linear models (GLMs) require assumptions about data distribution, dispersion, and seasonality that may not hold for emerging pathogens with changing transmission patterns.^20^ Previous studies have also defined epidemiological transitions based on the effective reproductive number crossing 1,^21,22^ and retrospectively via smoothing of an epidemic curve to identify the inflection points.^23^ Our approach defines surges as periods where a metric of disease transmission exceeds the 75^th^ percentile of a 26-week rolling window, with positive slope and a minimum threshold, if applicable. This non-parametric framework adapts automatically to shifting baselines without re-parameterization. This enables real-time implementation without waiting for sufficient data to confirm statistical tests; retrospective or lagged analysis when finding inflection points; or for accumulation of the multiple years of seasonal baselines. With surge periods defined, we are able to examine early warning signals that preceded them, beginning with the temporal variability patterns captured by the coefficient of variation.

### Temporal Variability

The elevated coefficient of variation we observed preceding and following surges aligns with theoretical expectations from overdispersion of infectious disease transmission. Lloyd-Smith et al. (2005) demonstrated that individual variation in infectiousness is fundamental to disease transmission, with most cases infecting few or no others, while overdispersed transmission events drive epidemic growth.^24^ During stochastic establishment phases, case counts are highly variable due to overdispersed transmission events, but as multiple transmission chains become active simultaneously, transmission becomes more uniform and predictable, reducing variability. The reverse occurs during post-surge fadeout: as susceptible hosts are depleted, transmission reverts back to sporadic chains, and variability rises again, before settling to baseline. These findings are consistent with characteristics of critical slowing down theory (CSD), which predicts elevated variance as systems approach tipping points,^4,5^ though applying CSD to epidemic systems requires reframing core assumptions about how this variability arises, and where the state change occurs. Traditional CSD theory requires that systems approach tipping points gradually with sufficient timescale separation—the system must equilibrate faster than conditions change for early warning indicators to develop.^21^ Recent attempts to apply CSD-based early warning signals to COVID-19 have repeatedly encountered these assumptions as barriers.^21–23,25^ The consistent thread across these studies is that COVID-19’s fast, re-emerging, and non-stationary dynamics violate the conditions CSD theory requires

The CSD early warning signal literature has predominantly focused on the R_T_ = 1 bifurcation point to detect either novel pathogen emergence (subcritical to supercritical transition) or confirm disease elimination (supercritical to subcritical).^6,26,27^ Theoretical work demonstrates that the coefficient of variation is mathematically uninformative for this transition, and is uninformative prior to disease emergence.^26,27^ Our finding that CV reliably signals pre-surge periods does not contradict these results, as we study a fundamentally different transition. Rather than detecting when a pathogen crosses the R_T_ = 1 threshold, we detect when an already-supercritical pathogen shifts from low to high transmission intensity. From a strict dynamical systems perspective, this is not a true bifurcation because R_T_ may not cross the threshold of 1 throughout, instead remaining above 1. Yet functionally, this transition represents exactly the state change that health systems must anticipate. We argue this transition is more operationally important than the R_T_ = 1 bifurcation for two reasons. First, detecting novel emergence requires surveillance infrastructure for pathogens not yet identified, and by the time laboratory methods, data pipelines, and monitoring systems exist, the pathogen has typically already established supercritical transmission (e.g. Covid-19). Second, elimination status is more directly assessed through vaccination coverage and herd immunity thresholds (1 - 1/R₀), than it is through bifurcation signatures. Notably, elimination has only been achieved through vaccination campaigns where immunized coverage, not CSD detection, guides decision-making.^28,29^ In contrast, surge detection for perpetually supercritical and endemic pathogens (influenza, RSV, COVID-19) represents the routine operational reality for health systems worldwide, and is what we have modified our CSD inspired framework to detect. Furthermore, our ability to identify bidirectional variability patterns means we can orient ourselves within the different stages of an epidemic, rather than focusing on a single point in time.

Considering these challenges, the dynamics and state changes underlying CSD theory may be better understood through overdispersion. In epidemic terms, the “perturbations” CSD describes are overdispersed transmission events. The “slow return to stable state” reflects transmission chains persisting rather than dying out. And the elevated variance captures the stochastic nature of transmission chains preceding and following a surge. This reframing suggests that the observable signatures of CSD can emerge from overdispersed transmission dynamics without requiring typical equilibrium conditions or bifurcation points. Overdispersion inherently produces high variability during establishment phases and pushes the system toward a state change —the transition from low to high transmission. We use CSD as a guide for the dynamical behavior we should expect to observe, while recognizing that operationalizing early warning for epidemic systems requires metrics that account for established diseases and non-stationarity, as well as mean-variance scaling. The approximately one-month lead time we observed with CV to signal surge onset may reflect the characteristic duration of stochastic establishment, the period required for overdispersed transmission dynamics to resolve through either chain extinction or successful surge establishment. Similarly, the post-surge rise in CV likely reflects the reverse process: as susceptible hosts are depleted during peak transmission, where overwhelming disease incidence transitions into sporadic transmission chains, subject to stochastic extinction and increased variability. Our approach captures the transmission characteristics at each time point without requiring quasi-equilibrium conditions or traditional bifurcation points.

### Spatial Variability (Global Moran’s I)

Our application of Global Moran’s I differs from the traditional objectives of confirming geographic clusters. Rather than asking where clustering occurs, whether case counts are anomalously high, or predicting spread to adjacent areas, we use the magnitude of spatial autocorrelation as a time-varying indicator of epidemic phase. Elevated spatial autocorrelation in our framework indicates that transmission remains localized and heterogeneous, the epidemiological signature of stochastic establishment before widespread community transmission. Declining global Moran’s I during surges reflects the transition to geographically similar transmission as the pathogen saturates susceptible networks. In epidemiological terms, the elevated global Moran’s I observed in the weeks preceding a surge likely reflects the same overdispersion dynamics captured by the CV. Overdispersed transmission events are typically localized, producing spatial clustering that diminishes as transmission chains multiply throughout the population.

Methodologically, global Moran’s I parallels the CV as a normalized metric; both measure deviation relative to a mean, but CV captures temporal variability within a time series while global Moran’s I captures spatial variability across geographic units. Recent spatial analysis supports this framing, where COVID-19 metrics showed extreme overdispersion that produced “chaotic” metapopulation dynamics.^25^ Conceptually similar work by Vega et al. (2025) showed that global Moran’s I calculated on spatially disaggregated unemployment data increased prior to economic crises in France, providing early warning signals that outperformed traditional univariate approaches. Their finding that “spatial early warning signal, global Moran’s I, is found to increase prior to the moments of economic crises”, supports the broader principle that rising spatial autocorrelation can signal impending transitions across complex adaptive systems. This reframes spatial autocorrelation as a system-state indicator, providing earlier warning by signaling that conditions favor potential surge establishment, rather than indicating that a localized outbreak has already begun.

### Limitations

Several limitations should be considered when interpreting our findings. First, our analysis was conducted retrospectively within a single state. Although the rolling-window framework is inherently prospective in its use of data, and we deliberately avoided parameter optimization to reduce overfitting risk, the parameters governing surge definition were evaluated post hoc against the full dataset, rather than through sequential application as data accumulated. Prospective validation in independent populations is needed to assess generalizability. Next, we interpret the observed variability patterns through the lens of overdispersed transmission dynamics and critical slowing down theory, but our surveillance metrics do not directly measure individual-level transmission heterogeneity or system resilience. The mechanistic link between overdispersion and the CV/Moran’s I signatures we observe remains inferential, consistent with theory but not a direct test of it. Finally, the moderate positive predictive values (34–54%) reflect our conservative week-by-week evaluation framework, which penalizes any individual week below threshold even when the broader pre or post-surge pattern is clearly present. When performance is assessed at the surge level, asking whether any week within the pre or post-surge period crosses the performance threshold, detection rates exceed 90% for pre-surge and 75% for post-surge periods. Moreover, even surges where individual weeks do not cross the threshold exhibit the characteristic directional pattern of rising CV followed by decline into the surge, suggesting the underlying signal is more consistent than week-level metrics alone indicate. The residual false positives likely reflect the stochastic nature of epidemic establishment: elevated variability signals conditions favor surge development, but whether a particular transmission chain successfully establishes or stochastically extinguishes depends on factors beyond the predictability of any surveillance-based model. In this framing, a ‘false positive’ may represent a genuine period of elevated transmission potential where chains happened to die out, rather than a failure of the indicator. This suggests that variability metrics may be most valuable when integrated with other predictors in ensemble approaches, rather than deployed as standalone decision triggers. Nevertheless, our week-level performance compares favorably to other early warning approaches: Looker et al. (2025) found that only 15% of simulations successfully signaled before critical transitions during re-emergence phases,^22^ and O’Brien and Clements (2021) reported highly variable detection across waves.^23^

## Conclusions

The consistency of these variability patterns across multiple data streams and indicator types suggests robustness to surveillance infrastructure. The approximately one-month lead time for both surge onset and termination could support healthcare surge planning, public health messaging, and resource pre-positioning. A key advantage of our approach is computational simplicity. Traditional CSD methods often require sophisticated mathematical and time series analysis, whereas our methods require only percentiles, moving averages, and standard summary statistics—calculations implementable in spreadsheet software by practitioners without training in applied math or dynamical systems. This accessibility matters: early warning systems that cannot be readily implemented do not help public health.

Our reframing of CSD early warning indicators by linking them to readily understood epidemiological characteristics provides an interpretable framework that can make early warning systems more accessible. Future work should investigate the observed variability patterns for other pathogens and should prioritize operationalizing these metrics as real-time alarm thresholds. Our work demonstrates that the overdispersed transmission dynamics preceding epidemic establishment and termination produce observable signatures in surveillance data — variability is not noise to be smoothed away, but signal.

## Methods

### Data Collection and Preparation

Wastewater surveillance data of SARS-CoV-2 were obtained from the New York State Wastewater Surveillance Network.^31^ The network analyzed samples collected from wastewater treatment plants (WWTPs) from April 2022 through December 2024. SARS-CoV-2 RNA concentrations were measured and reported as gene copies per milliliter. Samples from seven of the ten NYS economic development regions were analyzed by Quadrant Biosciences, with the remaining regions (Long Island, and New York City) analyzed by SUNY Stonybrook, and New York City, respectively. Extreme outliers were removed by excluding values above the 98th percentile within each region. For wastewater analysis, we excluded the Western New York region because samples from this region were processed by two different laboratories using distinct methodologies. These distinct methods resulted in an artificially wide range to the data and contributed to chronically high variability that was an artifact of methods rather than transmission status. Clinical COVID-19 cases and hospitalizations were obtained from the New York State Department of Health. Data were normalized to per 100,000 rates and aggregated to weekly intervals. Regional rates were calculated as population-weighted averages using county populations.

### Wastewater sample collection and processing

Wastewater samples were processed and analyzed for SARS-CoV-2 by three laboratories, each with different methods. Quadrant Laboratories processed samples using ultracentrifugation with a sucrose cushion and quantified SARS-CoV-2 concentration using reverse transcription quantitative polymerase chain reaction (RT-qPCR). Full documentation of these methods was previously published.^32^ The SUNY Stony Brook laboratory processed samples for this study from Long Island (Nassau and Suffolk counties). 24 hour composite samples of raw sewage were centrifuged at 4200 rpm for 30 min at 4°C to remove large particles and debris before polyethylene glycol (PEG) precipitation. Recovery rates were evaluated using bovine coronavirus (BCoV), which belongs to the same genus as SARS-CoV-2 and was spiked into the supernatant. The viral particles in 40 mL of samples were precipitated with PEG 8000 (Millipore Sigma, Burlington, MA) and NaCl (5 M, Millipore Sigma, Burlington, MA) and then incubated overnight at 4 °C. RNA from the PEG-precipitated wastewater was extracted by Qiagen QIAamp DSP viral RNA mini kit (Qiagen, Hilden, Germany) according to manufacturer’s instructions and eluted in 100 µL by nuclease-free water. The concentrations of RNA were measured by NanoDrop One Spectrophotometer (Thermo Fisher Scientific, Waltham, MA). All RNA samples were stored at −80 °C and subjected to cDNA synthesis within the same day of RNA extraction to avoid losses associated with storing and freezing and thawing RNA extracts.

Quantification for SUNY Stony Brook samples was done using reverse transcription by High Capacity RNA-to-cDNA Kit (Applied Biosystems, Waltham, MA) at 37 °C for 60 min, and stored at −20 °C until further analysis. The cycling condition was 95 °C for 10 min, followed by 40 cycles of 95 °C for 5 s and 55 °C for 40 s, and 98 °C for 10 min. The total volume of each reaction was 14.5 µL containing 7.25 µL of QuantStudio 3D Digital PCR Master mix v2 (Applied Biosystems, Massachusetts, USA), 0.725 µL of primer and probe (N1/ BCoV), 0.725 µL of TaqMan® Copy Number Reference Assay RNase P (as an internal control, Applied Biosystems, Waltham, MA), 4.8 µL of nuclease-free water, and 1 µL of cDNA template. Digital PCR was performed using N1 primers and probe set from 2019-nCoV CDC EUA Kit (IDT # 10006606) and BCoV set against the BCoV gene as an external reference on a QuantStudio 3D Digital PCR (Applied Biosystems, Massachusetts, USA). Nuclease-free water was used as non-template control (NTC) and plasmids containing the complete nucleocapsid gene from 2019-nCoV (IDT # 10006625) were used as a positive control. Data analysis was performed with the online version of the QuantStudio 3D AnalysisSuite Cloud Software.

New York City Department of Public Health (NYC DOH) analysed samples collected in the five NYC Burroughs. For extraction, 40 mL aliquots of the 24 h composite samples were first pasteurized (60 °C, 90 min), and then centrifuged (5000 × g, 4 °C, 10 min) to remove solids. The supernatant was filtered (0.22 μm, cellulose acetate) and then subjected to virus con-centration using polyethylene glycol (PEG) precipitation (addition of 4.0 g PEG and 0.9 g NaCl followed by overnight incubation at 4 °C, and centrifugation at 12000 × g at 4 °C for 120 min to pellet viruses). The supernatant was discarded and RNA (along with any DNA present) was extracted from the concentrated PEG pellet using the Qiagen QiaAmp Viral RNA Mini Kit with modifications (described in the ESI†).

From March 2020-May 2022: Concentrations of the SARS-CoV-2 N1 gene in wastewater samples (N1 gene copies/liter) were determined with RT-qPCR, with the calculation of the concentration of the SARS-CoV-2 RNA based on the comparison with a synthetic RNA standard. Calculation of the concentrations of the SARS-CoV-2 N1 gene in wastewater samples (N1 gene copies/liter) required: 1-reverse transcription quantitative polymerase chain reaction (RT-qPCR) of the samples; 2-comparison of values obtained in 1 with a synthetic SARS-CoV-2 standard. From May 2022-April 2023: N1 gene concentrations were calculated using the exact concentration of the synthetic RNA standard measured using reverse transcription droplet digital PCR (RT-ddPCR). All data from months prior to this dataset were recalculated using this method. From April 2023-Present: SARS-CoV-2 viral N1 gene copy determination was replaced by dPCR on a Qiagen (Qiagen, USA) QIAcuity. dPCR does not utilize a standard curve and values reported by the instrument are considered absolute. Samples from November 1,2022-April 11, 2023, were rerun using dPCR, the values determined by dPCR were ∼10-20 times higher those determined by RT-qPCR. Increases in viral load between RT-qPCR and dPCR were retroactively scaled to match existing methods.

### Regional Groupings

Analyses for temporal variability were conducted at two geographic scales: (1) NYS economic development regions, and (2) two broad regions designated as “Upstate” and “Downstate” to capture major epidemiological patterns across the state. Downstate was defined by the Mid-Hudson, Long Island, and New York City regions of New York, with Upstate being comprised of all remaining regions. Spatial clustering was analyzed for New York state as a whole, using county level data.

### Steps for defining surges

We developed a generalizable surge detection algorithm applicable to most epidemiological surveillance metrics through five iterative refinement stages. The algorithm uses a dynamic framework making it both suitable for emerging pathogens and adaptable to shifting transmission dynamics. First, we calculated the global empirical cumulative distribution function (ECDF) across all available data for each region. We classified time points as surge periods when values exceeded the 75^th^ percentile of the regional distribution.

Second, we expanded detection criteria to incorporate the rate of change. After calculating 30-day rolling slopes using linear regression, we computed global ECDFs for both observed values and slopes. Surge periods were identified when either observed values exceeded the 75^th^ percentile, or the slope exceeded the 75^th^ percentile (with a positive slope required), capturing both sustained high transmission and rapid increases.

Third, to account for temporal variability, seasonal patterns, and host-virus coevolution, we replaced global ECDFs with 26-week rolling window calculations. For each time point, ECDFs were computed using the preceding 26 weeks of data, with global ECDFs serving as a fallback when insufficient historical data existed. Surge criteria required either: (1) observed values exceeding the 75^th^ percentile, (2) slope exceeding the 75^th^ percentile with positive values, or (3) global percentiles of either metric above 75%.

Fourth, we applied a metric-specific minimum threshold representing a level below which transmission is considered minimal. We used the 25^th^ percentile as a minimum threshold, however, metric specific values that reflect low transmission status could also be substituted here. This modification prevented a false positive surge classifications triggered by slope criteria during periods of low transmission, where small changes can produce steep relative slopes despite negligible epidemiological significance. Both statistical (ECDF-based) and minimum threshold criteria were required for surge designation.

Fifth, the final refinements addressed temporal fragmentation. We bridged gaps of two weeks or less between surge periods, recognizing these interruptions as likely sampling artifacts rather than true epidemiological transitions. We also removed surge periods lasting three weeks or less to focus analysis on sustained transmission events. When filtering and bridging surges, the order of operations is important: bridging gaps before removing short surges prevents the erroneous removal of what are functionally continuous surge periods interrupted by noise in the data or brief sampling gaps. This two-step temporal filtering produced consolidated surge periods representing meaningful epidemic waves.

We chose to use the 75^th^ percentile of actual values and slopes to be the main defining criteria for defining the surge category. This was a simple number that worked well for our algorithm and performance metrics. This value may change with outbreak frequency and duration; therefore, it is likely to be pathogen specific. For the highest performance, a data mining strategy may optimize this parameter, such as the delta parameter used by MEM methods.^17,18^ We found that our general use of the 75^th^ percentile performed adequately well for all scenarios, and we did not pursue the minimal gains we may have gotten from a parameter optimization method. We felt this was a worthwhile tradeoff so that we did not subject this analysis to overfitting and the other challenges that come along with certain parameter optimization techniques.

### Epidemic Phase Classification

Following surge identification via our 5-stage algorithm, we classified each time point into five mutually exclusive categories representing different epidemic phases. The surge was defined above. Pre-surge periods were divided into two windows: 4-8 weeks before surge onset, and 0-4 weeks before surge onset. “Post-surge” periods begin immediately after a surge and can last a maximum of 8 weeks. If another surge is experienced less than 8 weeks after the prior surge, then the post surge category is overwritten as the respective pre-surge category. Time points not meeting criteria for surge or pre/post-surge periods were marked as “uncategorized”. This classification scheme enabled compartmentalized examination of time series behavior throughout complete epidemic cycles.

### Temporal Variability Calculation

To examine temporal variability across three different epidemiology surveillance measures, i.e., wastewater concentration (gene copies/ml wastewater), clinical case incidence (per 100,000), and hospitalization incidence (per 100,000), we calculated rolling standard deviations using a 30-day window for each region. For each time point, the standard deviation was computed using all measurements within the preceding 30 days for each epidemiology measure we were tracking, in each region. This approach captured local variability while maintaining sufficient sample size. We then computed the coefficient of variation (CV) as the ratio of standard deviation to the regional weekly population-weighted means for each epidemiology measure, providing a normalized estimate of relative variability comparable across the different units and magnitudes for the three data streams. CV was calculated for all three measures (wastewater intensity, clinical case incidence, and hospitalization incidence).

### Spatial Variability (Autocorrelation) Calculation

Spatial autocorrelation methods have been widely applied in infectious disease epidemiology, typically either confirming that a disease exhibits spatial clustering or predicting the geographic location of future outbreaks based on proximity to current clusters.^33^ The global Moran’s I is perhaps the most common tool to confirm that cases cluster geographically in order to subsequently identify hotspots for targeted intervention such as through a local Moran’s I or a spatial scan statistic.^34^ We conducted spatial autocorrelation analysis using the global Moran’s I statistic to evaluate geographic clustering of COVID-19 clinical cases across NYS counties. County boundaries were obtained from the 2020 US Census from the tigris package,^35^ and centroids were calculated for spatial analysis. For each week, we calculated global Moran’s I using k-nearest neighbor spatial weights (k=4) implemented through the spdep package.^36^ The k-nearest neighbor approach was chosen to ensure consistent neighbor relationships regardless of county size or shape. Two-week smoothing was applied to the resulting global Moran’s I time series to reduce noise while preserving temporal patterns. Global Moran’s I was only calculated for the clinical case incidence measure because wastewater data were too spatially disjointed, and the hospitalizations were too few.

### Statistical Analysis

We compared the distributions of CV and global Moran’s I for each epidemic phase using Kruskal-Wallis tests to assess overall differences among the categories. We test sequential comparisons between pairwise temporally adjacent categories to examine transitions through epidemic cycles. Wilcoxon rank-sum tests with Bonferroni correction were used to assess statistical significance for all pairwise comparisons.

To evaluate agreement of surge classifications across the three surveillance data streams, we calculated Cohen’s kappa statistics comparing the time series for wastewater, clinical cases, and hospitalizations at regional scales. We examined synchronous comparisons as well as temporally lagged comparisons for wastewater (lagged 1-2 weeks behind both clinical indicators) and for cases (lagged 1 week behind hospitalizations) to account for the expected temporal asynchrony of disease detection across the data streams. To assess category-specific agreement, we performed binary one-vs-rest kappa analyses, where each surge category from every region (“uncategorized”, “4-8 weeks pre-surge”, “0-4 weeks pre-surge”, “Surge”, “Post-surge”) was binarized against all other categories, and agreement was calculated on the resulting 2×2 classification. To obtain statewide Kappa values, we averaged region-specific estimates.

### Performance Evaluation

We evaluated the performance of CV and spatial autocorrelation (global Moran’s I) as early warning indicators for COVID-19 surges. For each metric, we calculated diagnostic test characteristics comparing the 0–4 week pre-surge period (condition positive) against the uncategorized and 4–8 week pre-surge periods (condition negative). The alarm threshold was set at the 25th percentile of values observed during 0–4 week pre-surge periods, such that approximately 75% of true pre-surge periods would exceed the threshold by design. Surge and post-surge periods were excluded from the pre-surge diagnostic evaluation because the algorithm already reliably identifies these phases once they occur, and the purpose of investigating variability metrics was to detect impending surges before they became identifiable by the surge algorithm. By excluding these periods, the evaluation focuses specifically on the ability of each metric to distinguish the immediate pre-surge window from uncategorized and early pre-surge periods.

We additionally evaluated an extended 0–8 week pre-surge window to assess whether the variability signal emerges earlier than four weeks before surge onset. Year-specific subsets (2022, 2023, 2024) were also evaluated to examine how diagnostic performance varied across distinct pandemic eras characterized by different variants, immunity levels, and transmission dynamics.

For surge-end detection, we evaluated CV as an indicator of impending surge termination. The condition positive was defined as the metric falling below the 50th percentile of values observed during the surge category, and the surge ending within the following four weeks. This tests whether low variability during an active surge reliably signals that the surge is approaching termination.

Beyond week-level diagnostics, we assessed event-level detection rates by asking whether each signal fired at least once during the relevant pre-surge or surge-end window for each surge cycle. This complements the week-level analysis, which penalizes any individual week below threshold even when the broader pattern across the period is clearly present.

For each data stream and evaluation framework, we calculated sensitivity, specificity, positive predictive value (PPV), and negative predictive value (NPV).

All analyses were performed using R version 4.3.2 ^37^. Data, data preparation scripts, analysis code, and figure generation scripts are publicly available at: https://github.com/moranjoe/Variability---Surge-Prediction.

## Data Availability

All data is publicly available, and code is provided via a public github repository

https://health.data.ny.gov/Health/New-York-State-Statewide-COVID-19-Wastewater-Surve/hdxs-icuh/about_data

https://health.data.ny.gov/Health/New-York-State-Statewide-COVID-19-Testing/jvfi-ffup/about_data

https://health.data.ny.gov/Health/New-York-State-Statewide-COVID-19-Hospitalizations/jw46-jpb7/about_data

https://github.com/moranjoe/Variability---Surge-Prediction

## Author Contributions

**E. Joe Moran:** Conceptualization, Methodology, Formal analysis, Writing – original draft, Writing – review & editing.

**Dustin T. Hill:** Conceptualization, Methodology, Writing – review & editing.

**Mohammed A. Alazawi:** Conceptualization, Methodology, Writing – review & editing.

**David A. Larsen:** Conceptualization, Methodology, Writing – review C editing.

