## Supplemental material for "Noise is the signal: variability in surveillance data provide early indication of epidemic phase"

#### Supplemental Figures: Surge Detection Algorithm

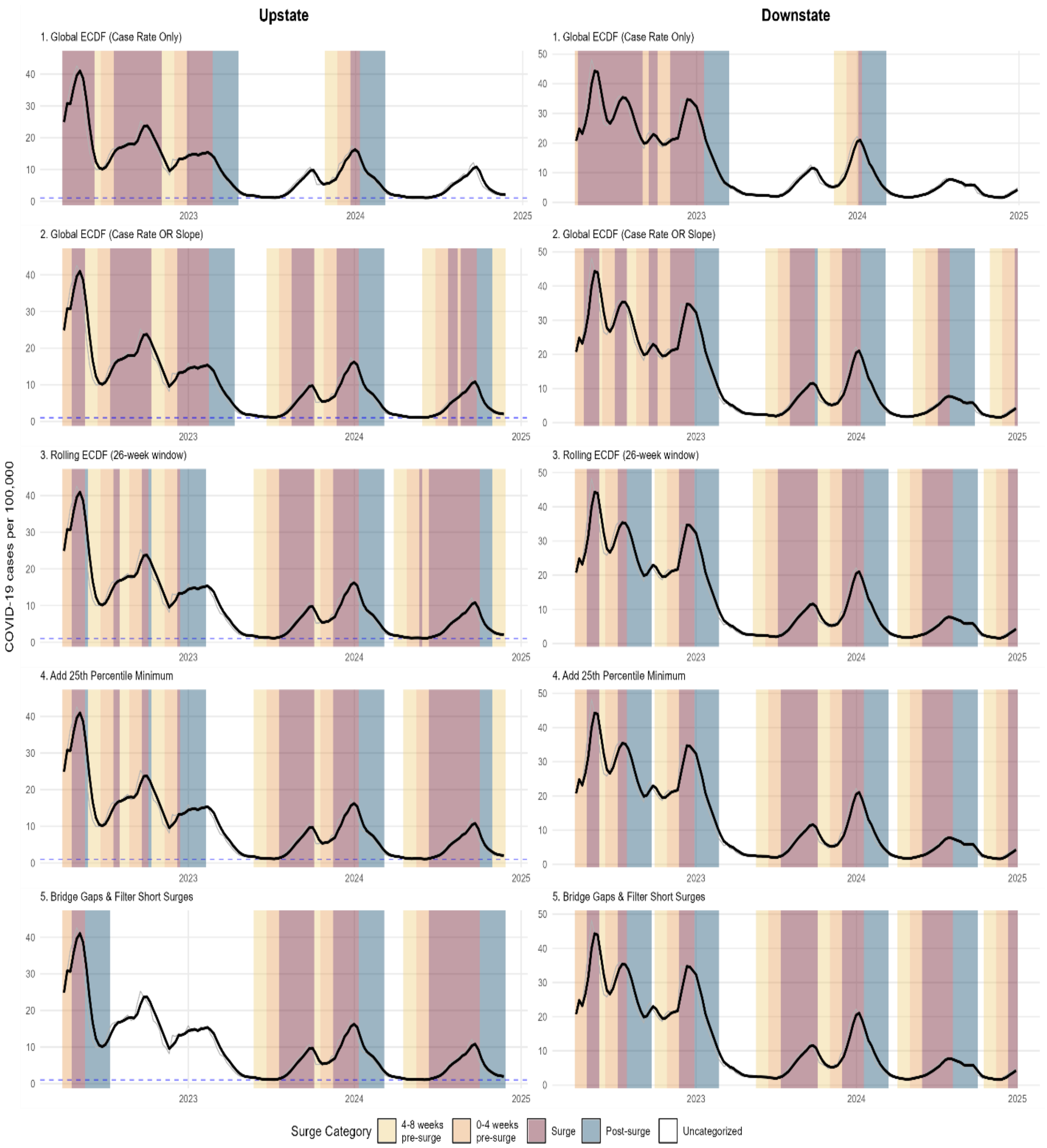


**Figure S1. Progressive development of the surge detection algorithm for clinical case data, showing five iterative refinement stages for Upstate and Downstate New York.**

Each panel displays Sars-Cov-2 clinical cases over time with background shading indicating classified epidemic phases, as seen in the legend. Stage 1 only used global ECDF of intensity to classify surges above the 75th percentile. Stage 2 added ECDF slope criteria of 75% to capture rapid increases. Stage 3 replaced global ECDFs calculated on all data points with 26-week rolling windows to adapt to evolving transmission dynamics. Stage 4 incorporated a minimum intensity threshold (25$^{th}$ percentile) to eliminate false positives during low-circulation periods. Stage 5 applied temporal filtering to bridge gaps shorter than two weeks and remove periods classified as a surge lasting less than three weeks.

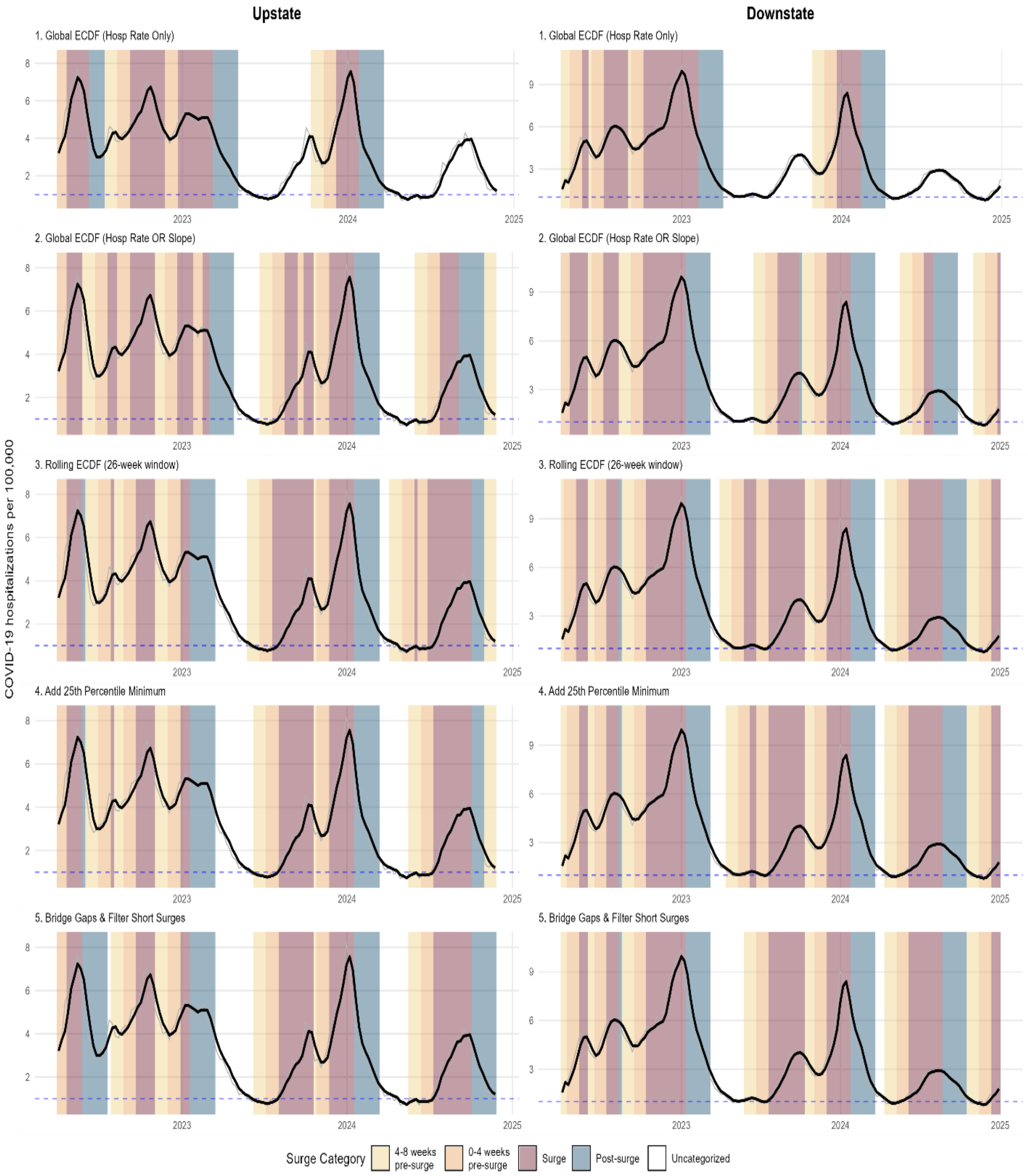


**Figure S2. Progressive development of the surge detection algorithm for hospitalizations, showing five iterative refinement stages for Upstate and Downstate New York.** Each panel displays Sars-CoV-2 hospitalizations over time with background shading indicating classified epidemic phases, as seen in the legend. Stage 1 only used global ECDF of intensity to classify surges above the 75th percentile. Stage 2 added ECDF slope criteria of 75% to capture rapid increases. Stage 3 replaced global ECDFs calculated on all data points with 26-week rolling windows to adapt to evolving transmission dynamics. Stage 4 incorporated a minimum intensity threshold (25th percentile) to eliminate false positives during low-circulation periods. Stage 5 applied temporal filtering to bridge gaps shorter than two weeks and remove periods classified as a surge lasting less than three weeks.

#### Supplemental Figures: Kappa Statistic

**Table S1. Standard Cohen's Kappa by Region.** This table shows agreement in surge category classification between wastewater, clinical case, and hospitalization data streams across New York State economic development regions. Kappa values were calculated for each region independently, with Statewide representing the mean across regions. Lag structures (1–2 weeks) were applied to wastewater and clinical cases to account for expected temporal delays between surveillance signals. Values are interpreted as slight (0.00–0.20), fair (0.21–0.40), moderate (0.41–0.60), substantial (0.61–0.80), or almost perfect (0.81–1.00) agreement. Lagged cases vs. hospitalizations showed the highest agreement across most regions, while wastewater vs. hospitalizations showed the weakest agreement.

| Comparison | Statewide | Central New York | Finger Lakes | Long Island | Mid-Hudson | Mohawk Valley | New York City | North Country | Southern Tier |
|---|---|---|---|---|---|---|---|---|---|
| WW vs Cases | **0.547** | 0.506 | 0.270 | 0.698 | 0.586 | 0.509 | 0.603 | 0.477 | 0.526 |
| WW vs Hosp | **0.355** | 0.110 | 0.247 | 0.623 | 0.583 | 0.185 | 0.555 | 0.290 | 0.099 |
| Cases vs Hosp | **0.558** | 0.458 | 0.547 | 0.682 | 0.586 | 0.276 | 0.688 | 0.500 | 0.524 |
| WW (lag 1wk) vs Cases | **0.511** | 0.543 | 0.221 | 0.666 | 0.433 | 0.366 | 0.640 | 0.450 | 0.639 |
| WW (lag 1wk) vs Hosp | **0.424** | 0.220 | 0.290 | 0.648 | 0.599 | 0.224 | 0.662 | 0.414 | 0.128 |
| WW (lag 2wk) vs Hosp | **0.440** | 0.391 | 0.301 | 0.578 | 0.482 | 0.239 | 0.636 | 0.508 | 0.233 |
| Cases (lag 1wk) vs Hosp | **0.640** | 0.633 | 0.710 | 0.737 | 0.536 | 0.411 | 0.699 | 0.564 | 0.560 |

**Table S2. Dichotomized Cohen's Kappa by Surge Category.** This table shows one-vs-rest agreement for each surge phase across data stream pairs. For each category, observations were dichotomized (target category vs. all others) and kappa was calculated from the resulting 2×2 confusion matrix, pooled across all regions. Pre-surge categories showed fair to moderate agreement ($\kappa$ = 0.263–0.542), while the surge and post surge periods showed stronger agreement ($\kappa$ = 0.395–0.786), indicating greater classification agreement for the different data streams once surge onset commences.

| Comparison | 4-8 weeks pre-surge | 0-4 weeks pre-surge | Surge | Post-surge |
|---|---|---|---|---|
| WW vs Cases | 0.528 | 0.429 | 0.600 | 0.635 |
| WW (lag 1wk) vs Cases | 0.441 | 0.367 | 0.569 | 0.651 |
| WW vs Hosp | 0.314 | 0.263 | 0.395 | 0.533 |
| WW (lag 2wk) vs Hosp | 0.324 | 0.324 | 0.485 | 0.619 |
| Cases vs Hosp | 0.447 | 0.447 | 0.623 | 0.737 |
| Cases (lag 1wk) vs Hosp | 0.521 | 0.542 | 0.710 | 0.786 |

### Supplemental Figures: Wastewater Surveillance

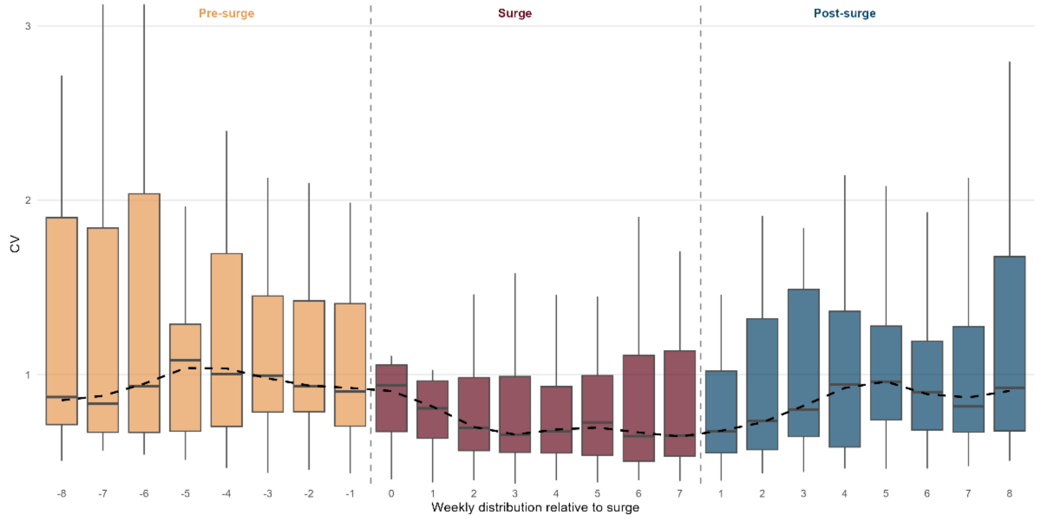


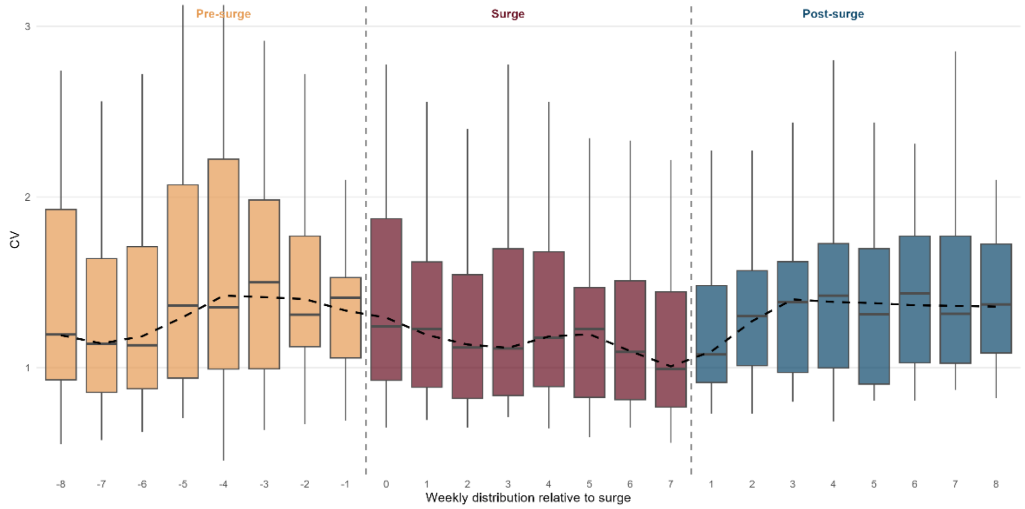


**Figure S3. Distribution of coefficient of variation (CV) by week relative to surge onset and termination for wastewater concentration for Downstate (top) and Upstate (bottom).** Boxplots show the distribution of CV values at each weekly position, pooled across all surges and regions, with weeks aligned relative to surge onset (left dashed line) and surge termination (right dashed line). Surge duration varies across events; the surge panel displays weeks 0 through 7 as a representative range encompassing observed surge lengths. Pre-surge weeks show elevated CV values peaking approximately 3–5 weeks before surge onset, followed by a decline into the surge. Surge weeks exhibit lower CV with a declining trend through active transmission. Post-surge weeks show a transient rise in CV during early fadeout before stabilizing. The dashed black line traces the median CV across weekly positions. This pattern illustrates the full bidirectional early warning signals present on either side of the surge.

Figures below show: Coefficient of variation analysis for wastewater concentration in NYS regions. Panels show: **A)** wastewater concentration time series raw values (grey) with 3-week smoothing (black), **B)** coefficient of variation over time with 25th percentile threshold of pre-surge periods (dashed) and 50th percentile threshold of surge periods (dotted), and **C)** distribution of CV values by surge category and threshold lines. Background colors indicate epidemic phases, as shown in the legend.

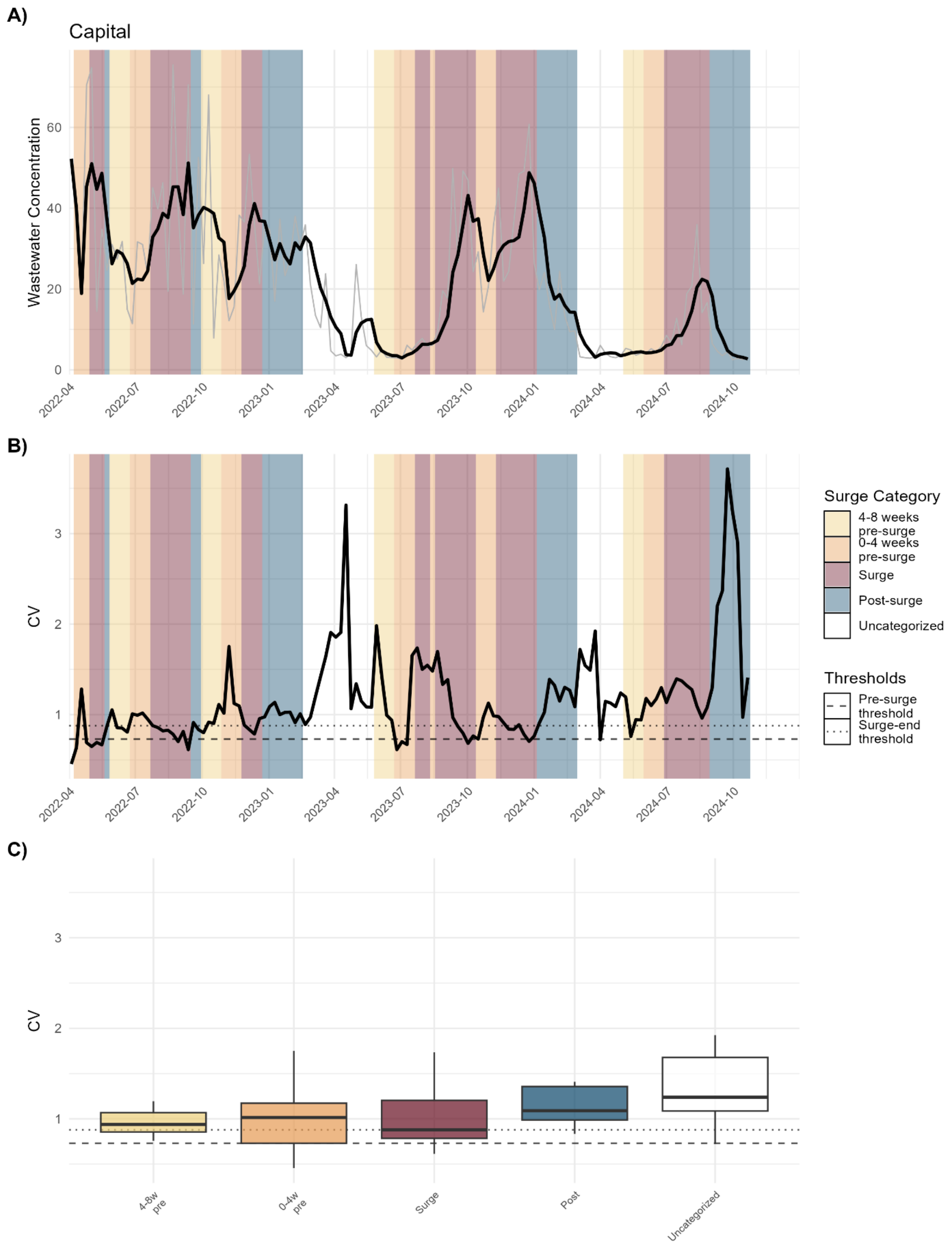


**Figure S4.** Wastewater CV of Capital Region

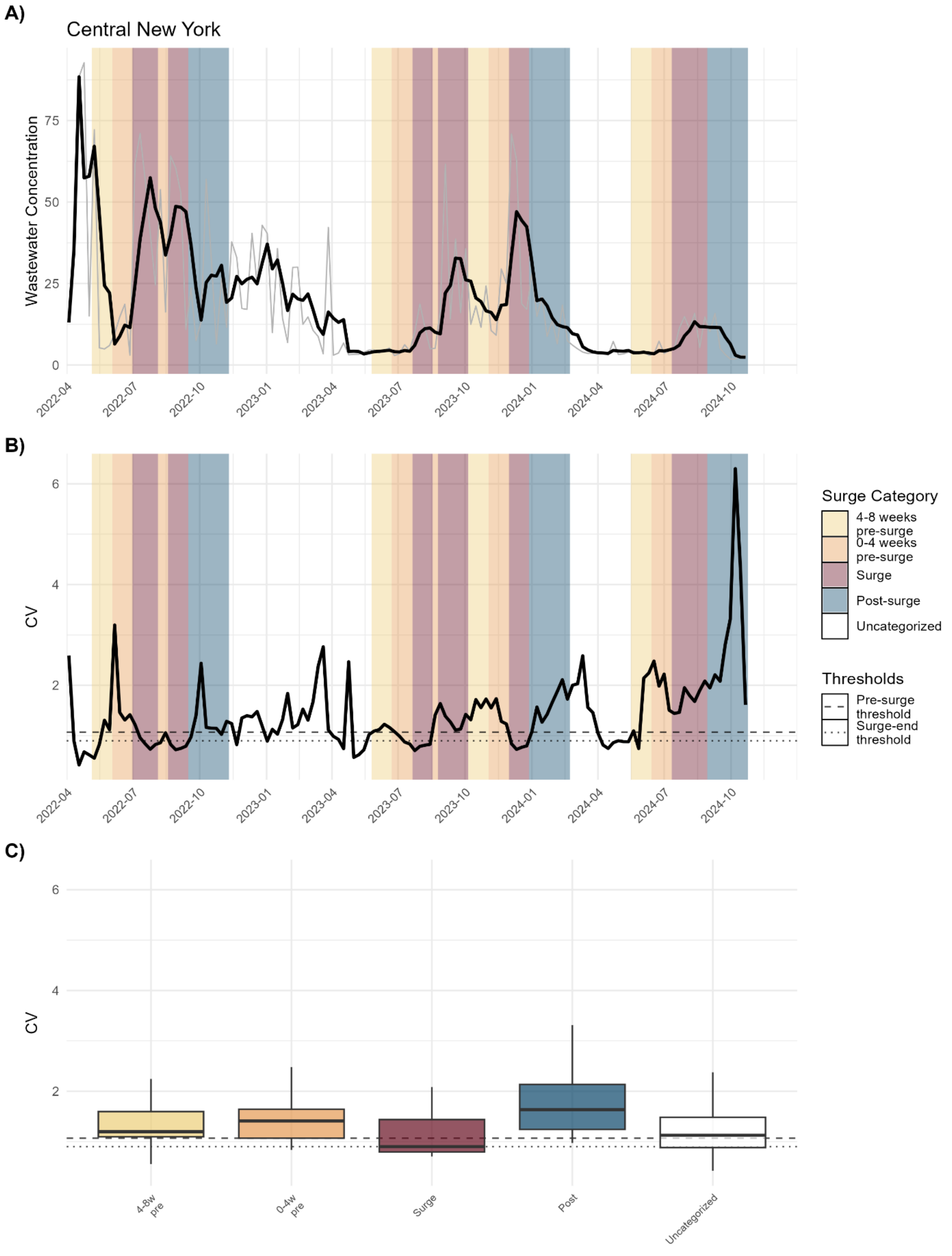


**Figure S5.** Wastewater CV of Central New York

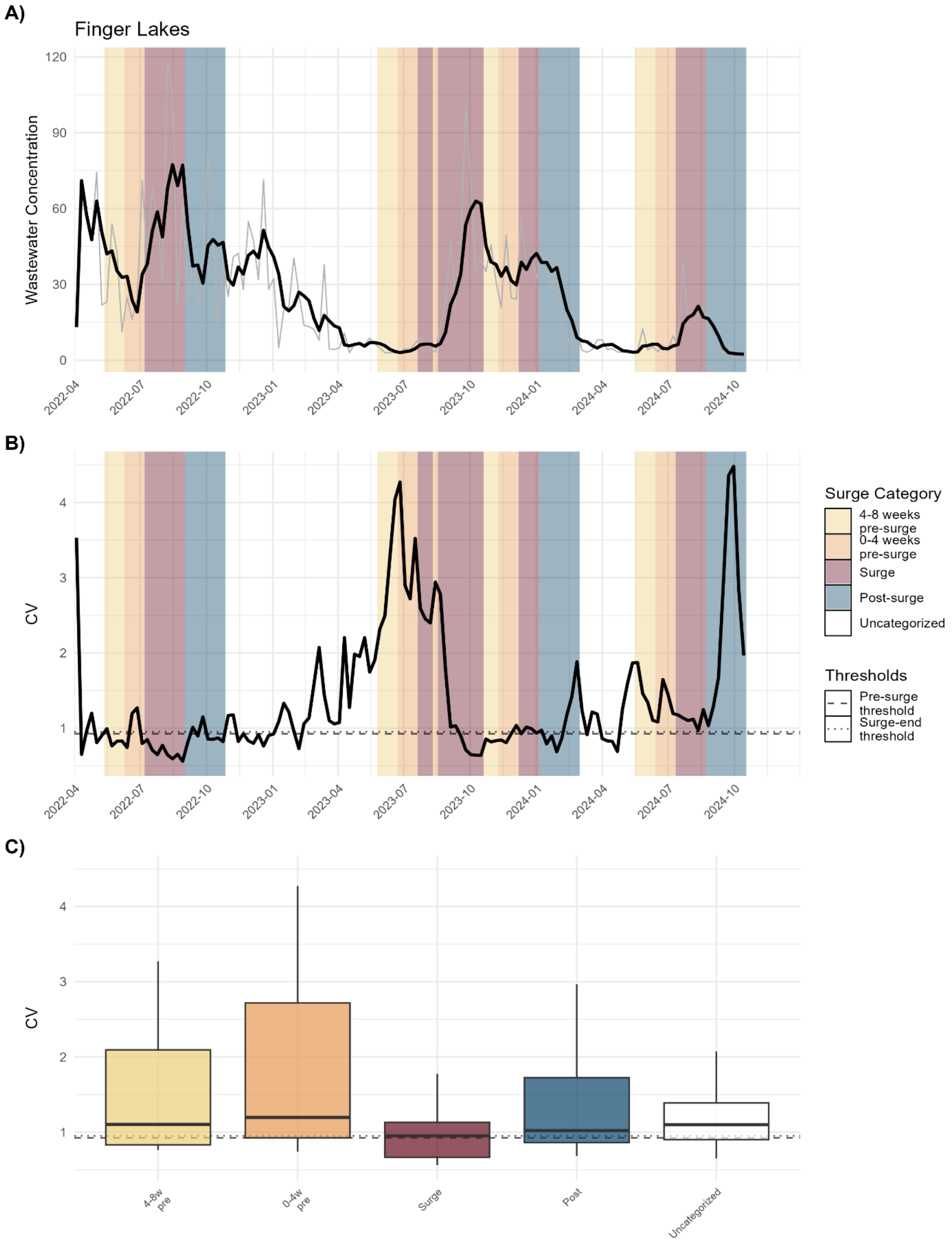


**Figure S6.** Wastewater CV of Finger Lakes

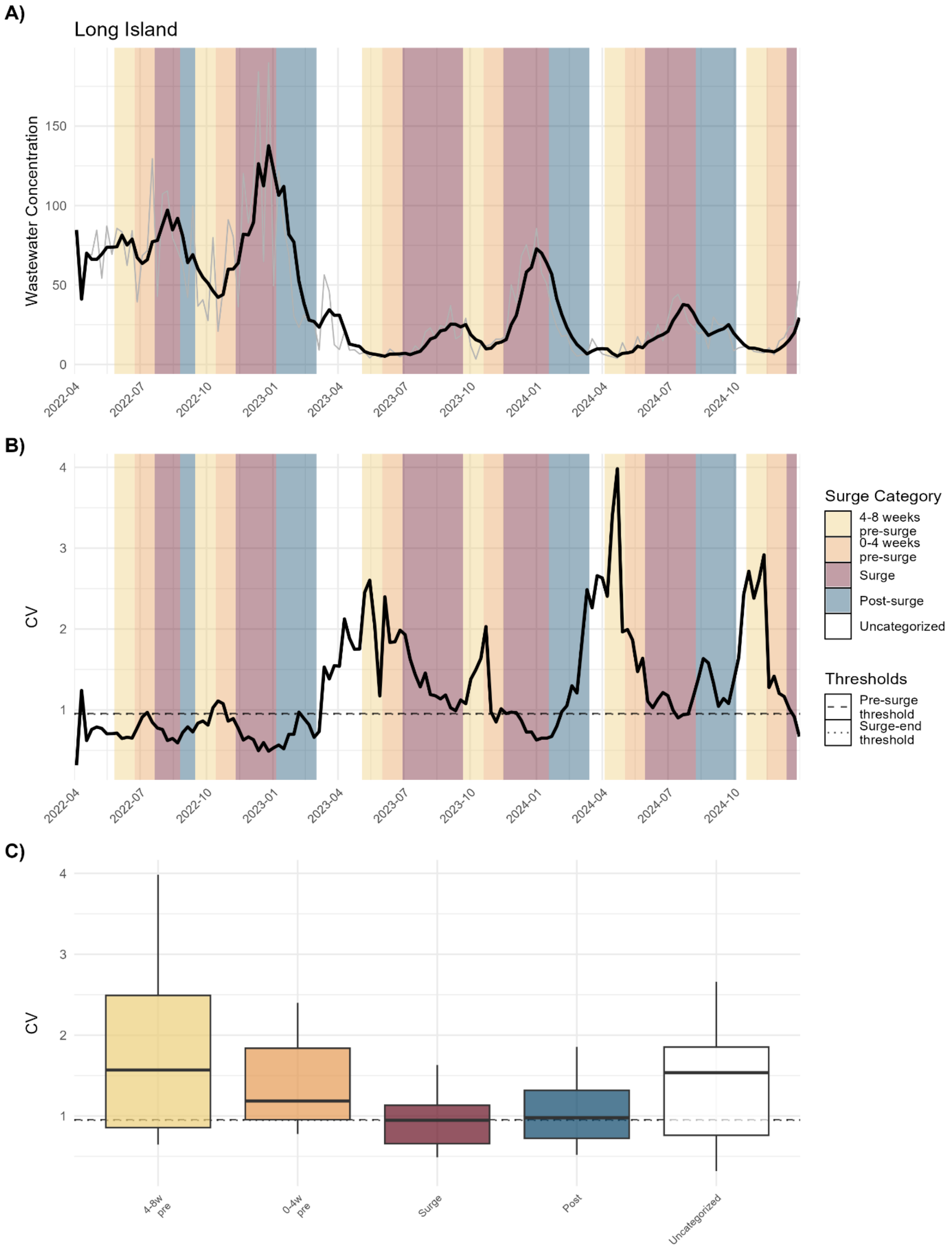


**Figure S7.** Wastewater CV of Long Island

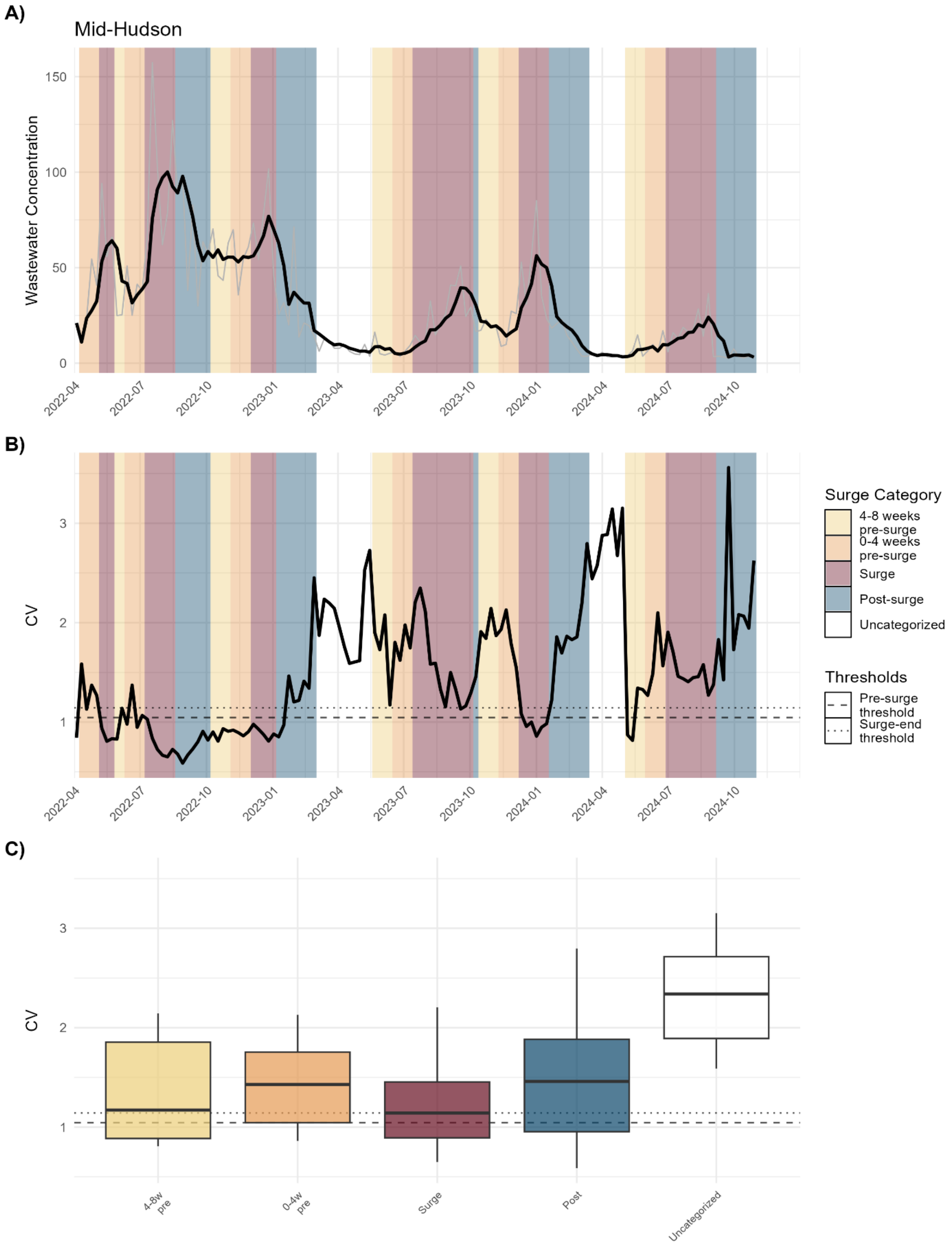


**Figure S8.** Wastewater CV of Mid-Hudson

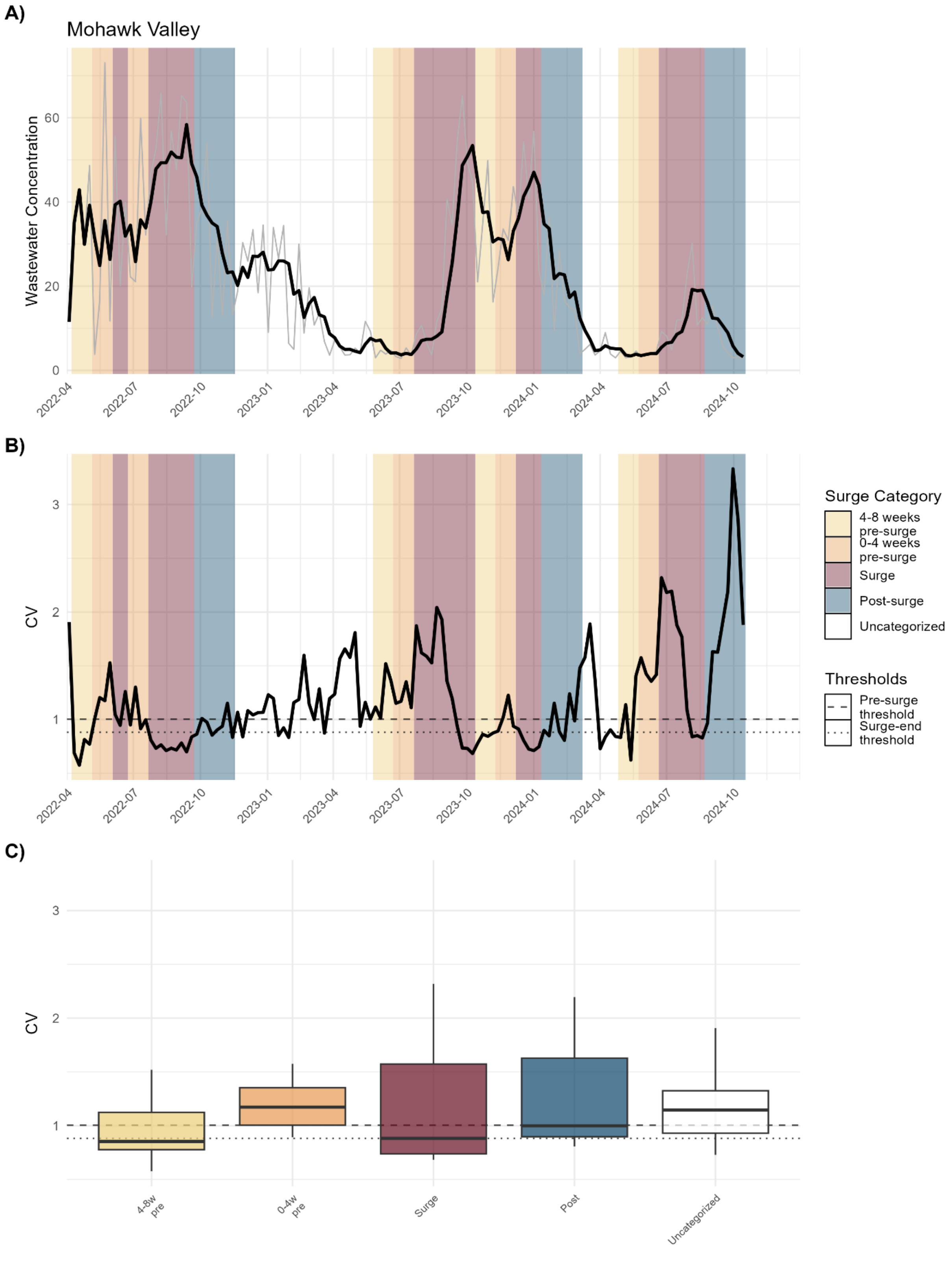


**Figure S9.** Wastewater CV of Mohawk Valley

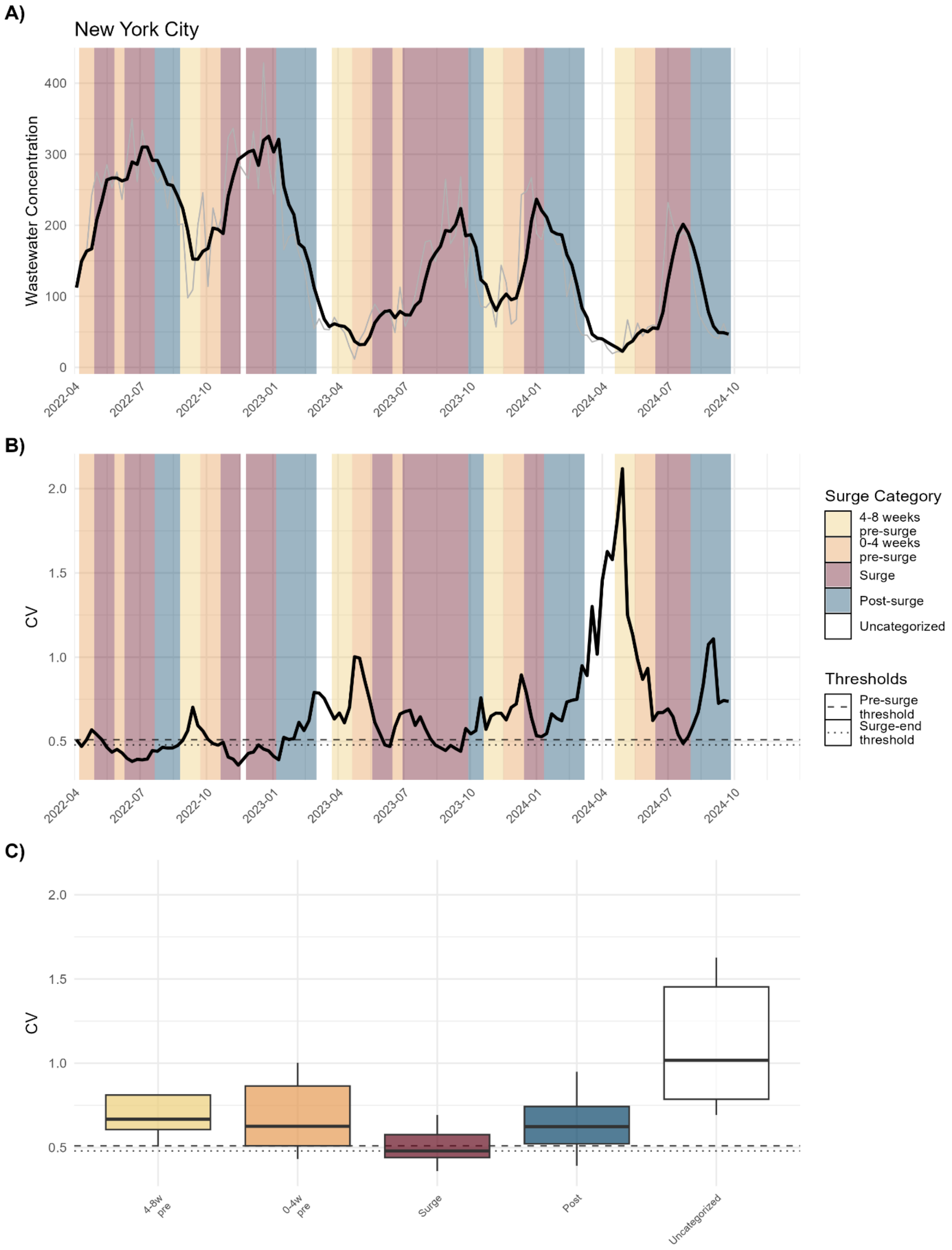


**Figure S10.** Wastewater CV of New York City

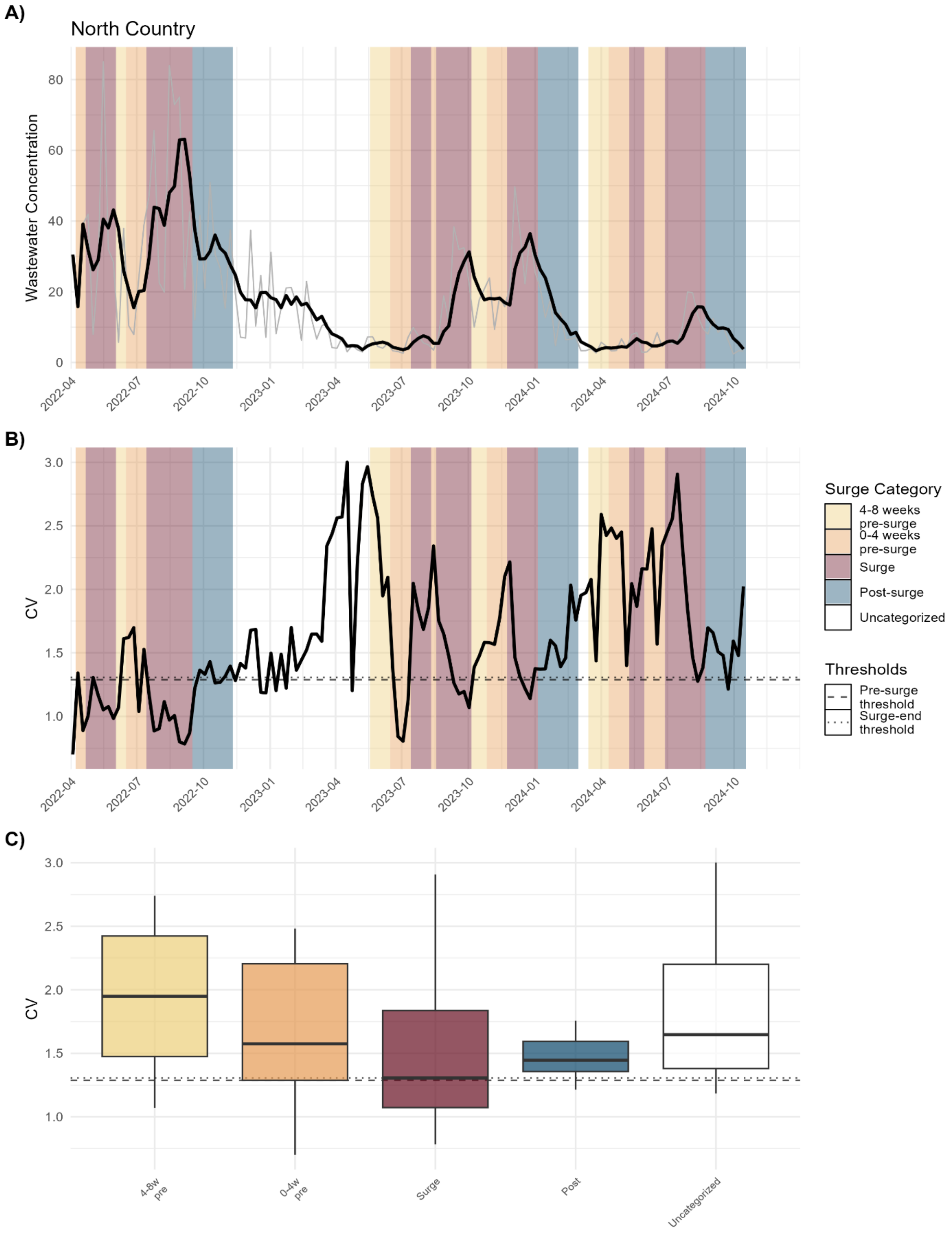


**Figure S11.** Wastewater CV of North Country

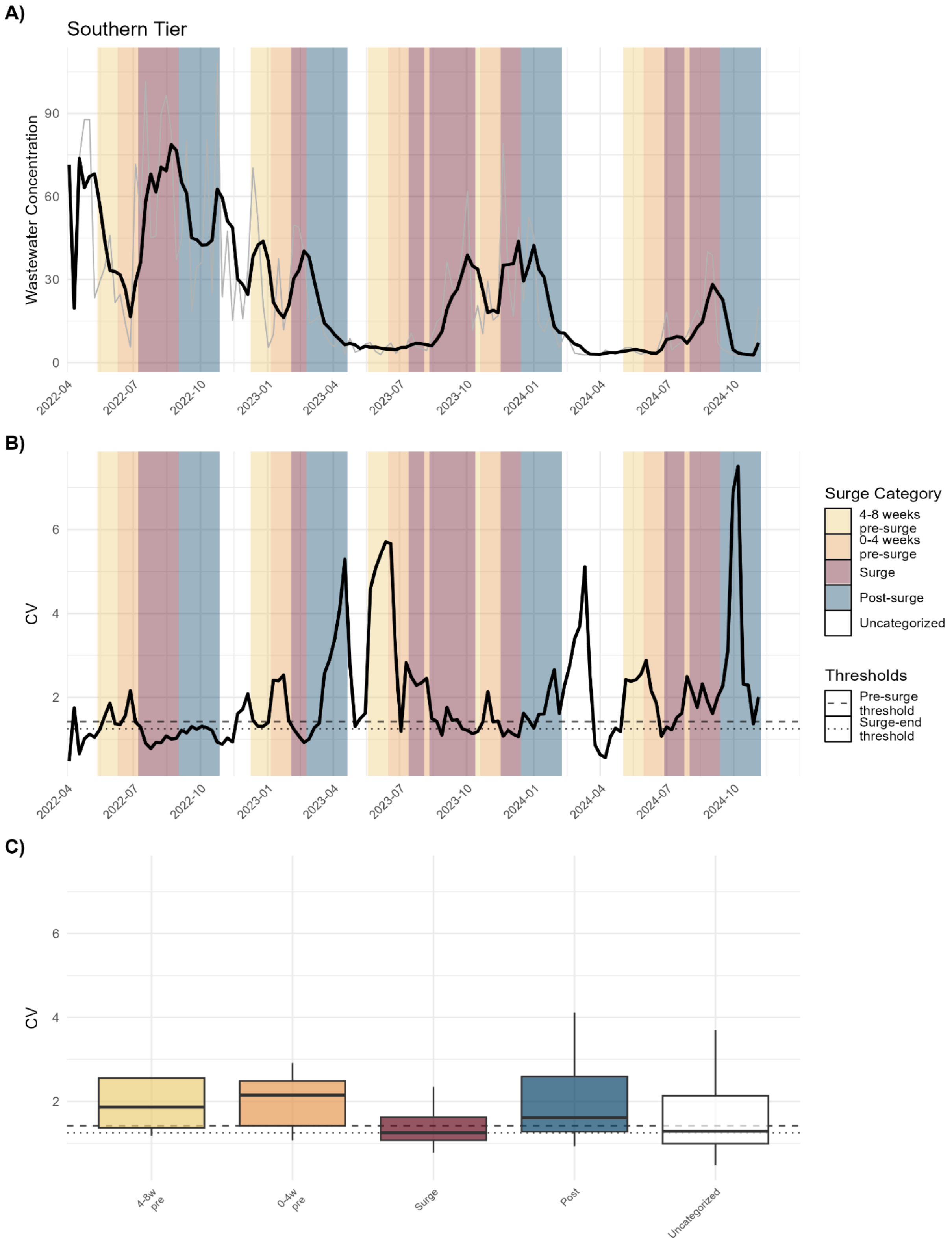


**Figure S12.** Wastewater CV of Southern Tier

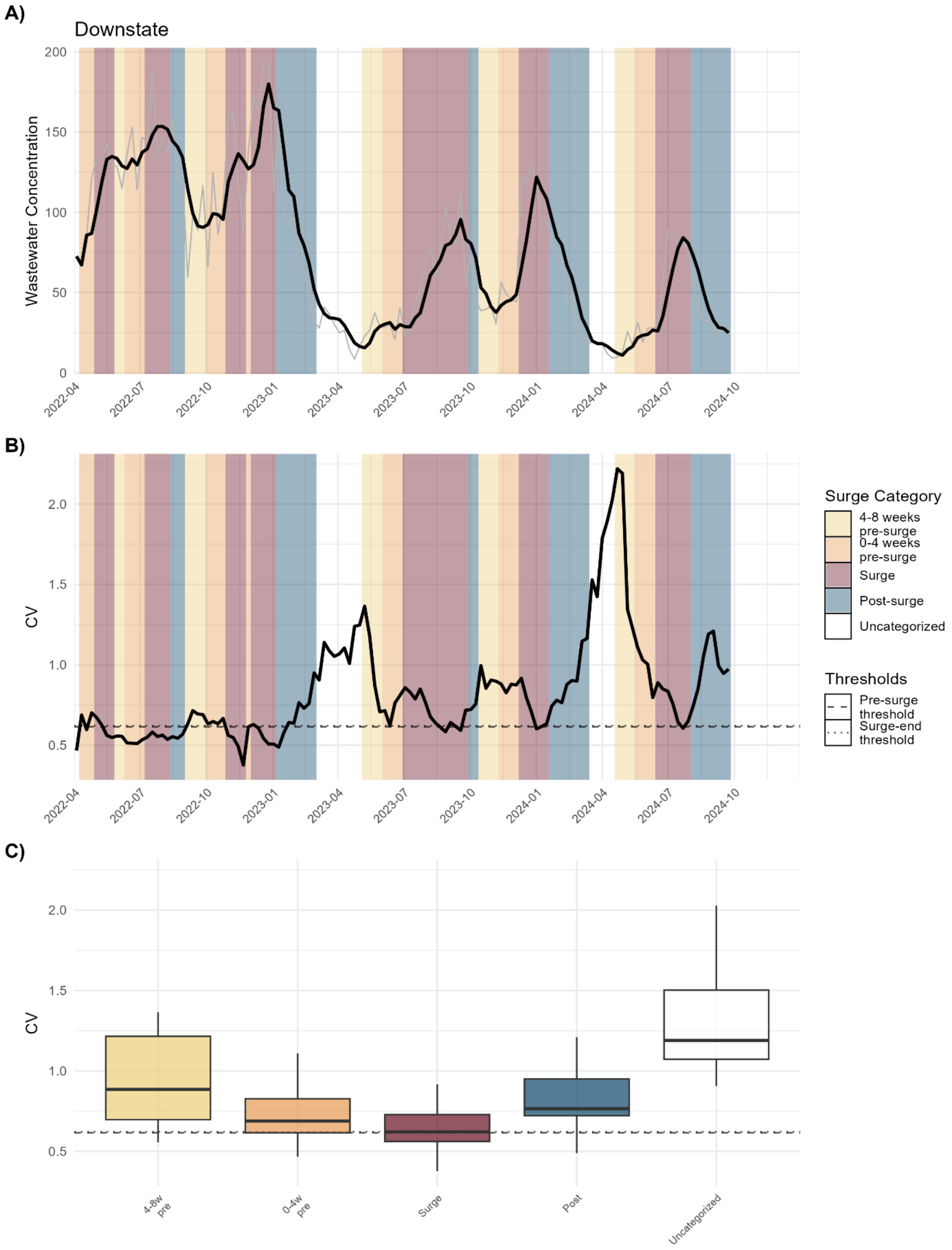


**Figure S13.** Wastewater CV of Downstate

#### Wastewater Kruskal-Wallis Pairwise Comparisons

These tables present pairwise Wilcoxon rank-sum test results comparing coefficient of variation (CV) across adjacent surge categories. Adjacent category comparison includes the two surge categories being compared, the number of weekly observations in each group, the median CV for each group, the unadjusted Wilcoxon rank-sum test p-value, and the Bonferroni-corrected p-value adjusted for multiple comparisons. Green highlighting indicates adjusted $p < 0.05$; orange indicates $0.05 \le$ adjusted $p < 0.10$.

**Table S3.** CV adjacent comparisons of all regions (wastewater)

| Category 1 | Category 2 | $n_1$ (weeks) | $n_2$ (weeks) | Median $_1$ | Median $_2$ | p-value | Adjusted p |
|---|---|---|---|---|---|---|---|
| Uncategorized | 4-8 weeks pre-surge | 373 | 160 | 1.203 | 1.168 | 0.688 | 1 |
| 4-8 weeks pre-surge | 0-4 weeks pre-surge | 160 | 211 | 1.168 | 1.204 | 0.540 | 1 |
| 0-4 weeks pre-surge | Surge | 211 | 371 | 1.204 | 0.969 | 1.14e-08 | 5.69e-08 |
| Surge | Post-surge | 371 | 262 | 0.969 | 1.218 | 5.86e-08 | 2.93e-07 |
| Post-surge | Uncategorized | 262 | 373 | 1.218 | 1.203 | 0.899 | 1 |

**Table S4.** CV adjacent comparisons of downstate (wastewater)

| Category 1 | Category 2 | $n_1$ (weeks) | $n_2$ (weeks) | Median 1 | Median 2 | p-value | Adjusted p |
|---|---|---|---|---|---|---|---|
| Uncategorized | 4-8 weeks pre-surge | 28 | 18 | 1.190 | 0.885 | 0.009618 | 0.048092 |
| 4-8 weeks pre-surge | 0-4 weeks pre-surge | 18 | 25 | 0.885 | 0.689 | 0.011367 | 0.056837 |
| 0-4 weeks pre-surge | Surge | 25 | 44 | 0.689 | 0.622 | 0.111873 | 0.559365 |
| Surge | Post-surge | 44 | 29 | 0.622 | 0.765 | 0.000184 | 0.000922 |
| Post-surge | Uncategorized | 29 | 28 | 0.765 | 1.190 | 8.92e-07 | 4.46e-06 |

**Table S5.** CV adjacent comparisons of upstate (wastewater)

### Supplemental Figures: Clinical Cases

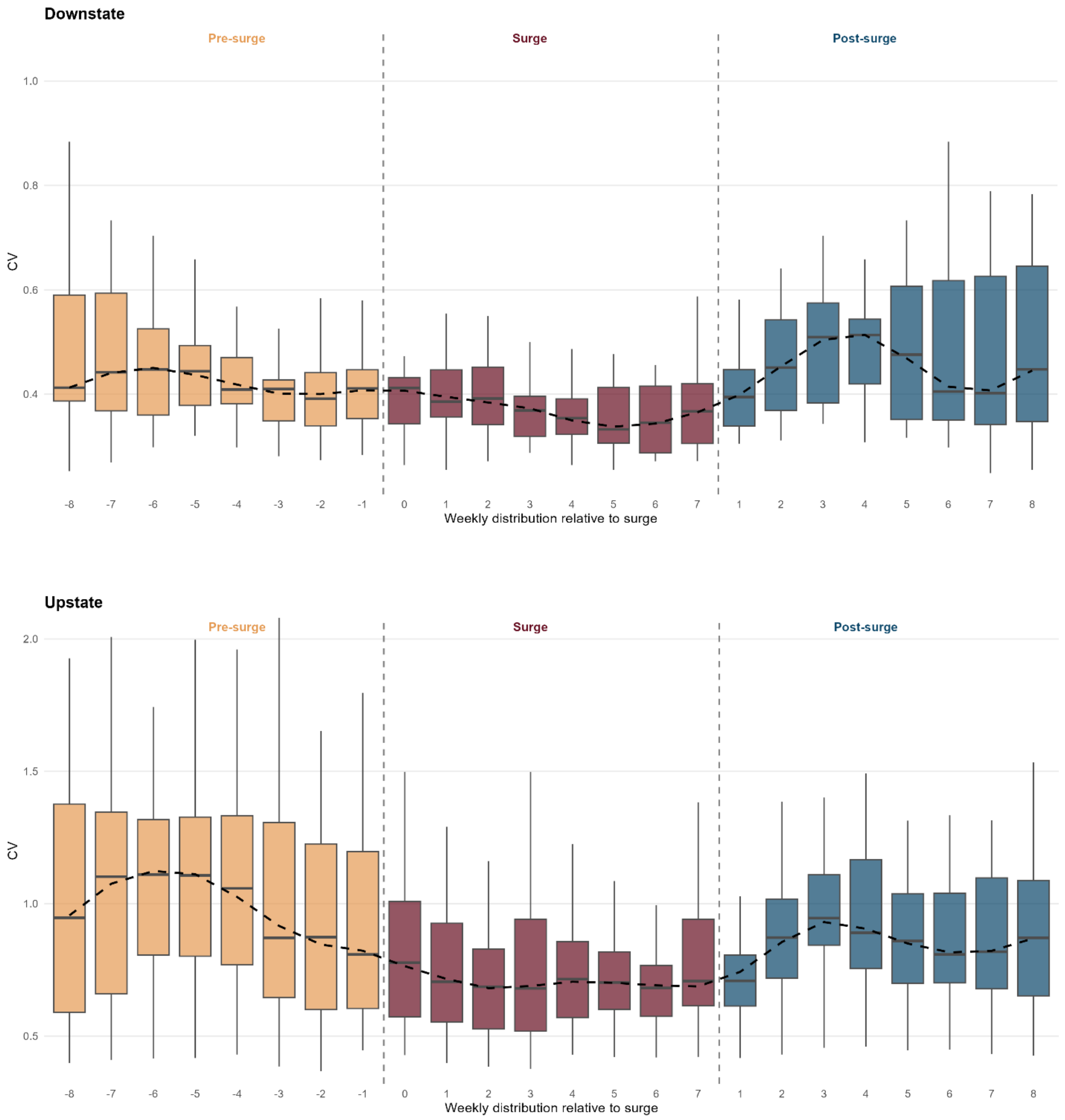


**Figure S14. Distribution of coefficient of variation (CV) by week relative to surge onset and termination for clinical cases for Downstate (top) and Upstate (bottom).** Boxplots show the distribution of CV values at each weekly position, pooled across all surges and regions, with weeks aligned relative to surge onset (left dashed line) and surge termination (right dashed line). Surge duration varies across events; the surge panel displays weeks 0 through 7 as a representative range encompassing observed surge lengths. Pre-surge weeks show elevated CV values peaking approximately 5-6 weeks before surge onset, followed by a decline into the surge. Surge weeks exhibit lower CV with a declining trend through active transmission. Post-surge weeks show a transient rise in CV during early fadeout before stabilizing. The dashed black line traces the median CV across weekly positions. This pattern illustrates the full bidirectional early warning signals present on either side of the surge.

Figures below show: Coefficient of variation analysis for clinical cases. Panels show: **A)** clinical case time series raw values (grey) with 3-week smoothing (black), **B)** coefficient of variation over time with 25th percentile threshold of pre-surge periods (dashed) and 50th percentile threshold of surge periods (dotted), and **C)** distribution of CV values by surge category and threshold lines. Background colors indicate epidemic phases, as shown in the legend.

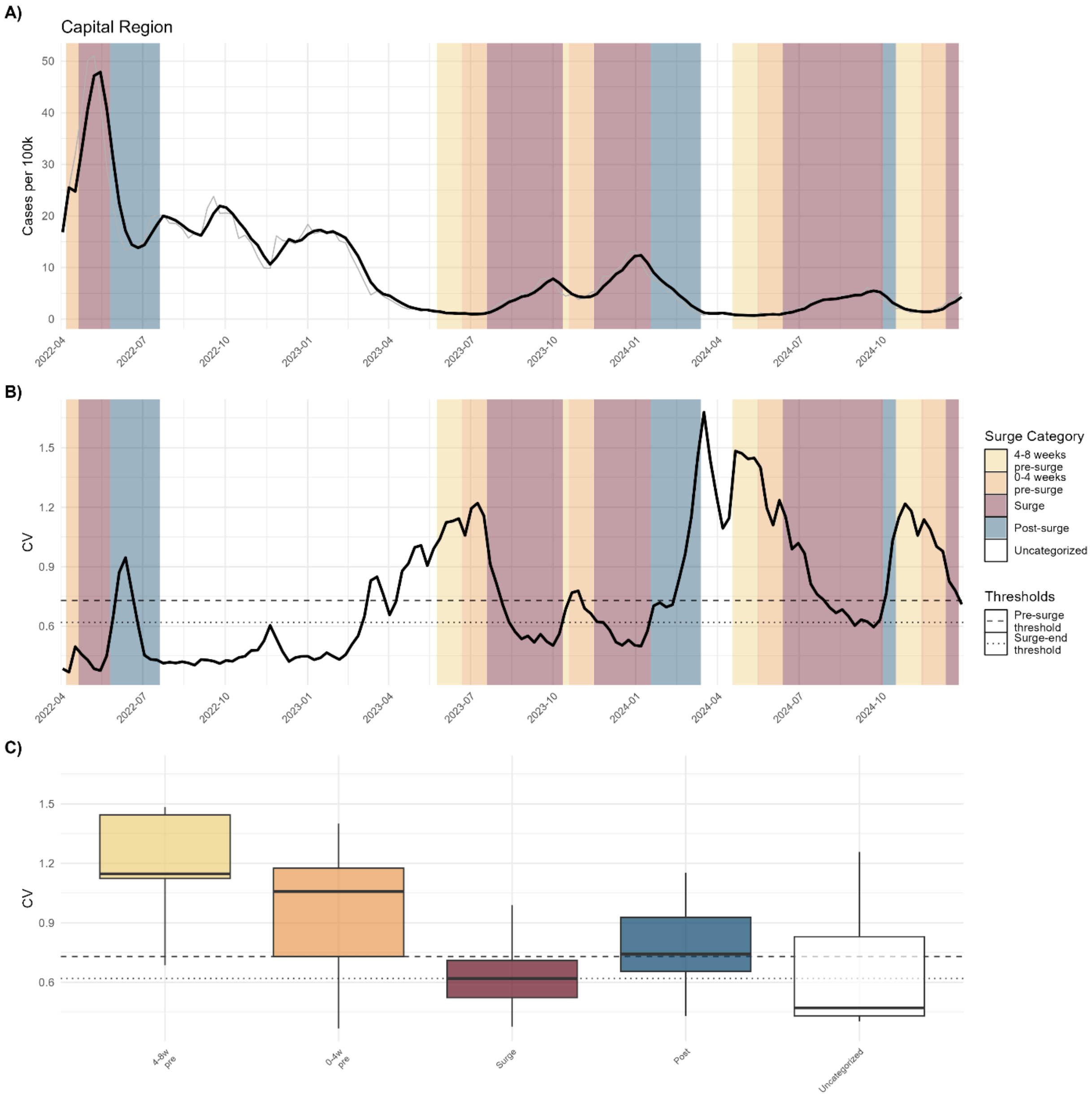


**Figure S15.** Cases CV of Capital Region

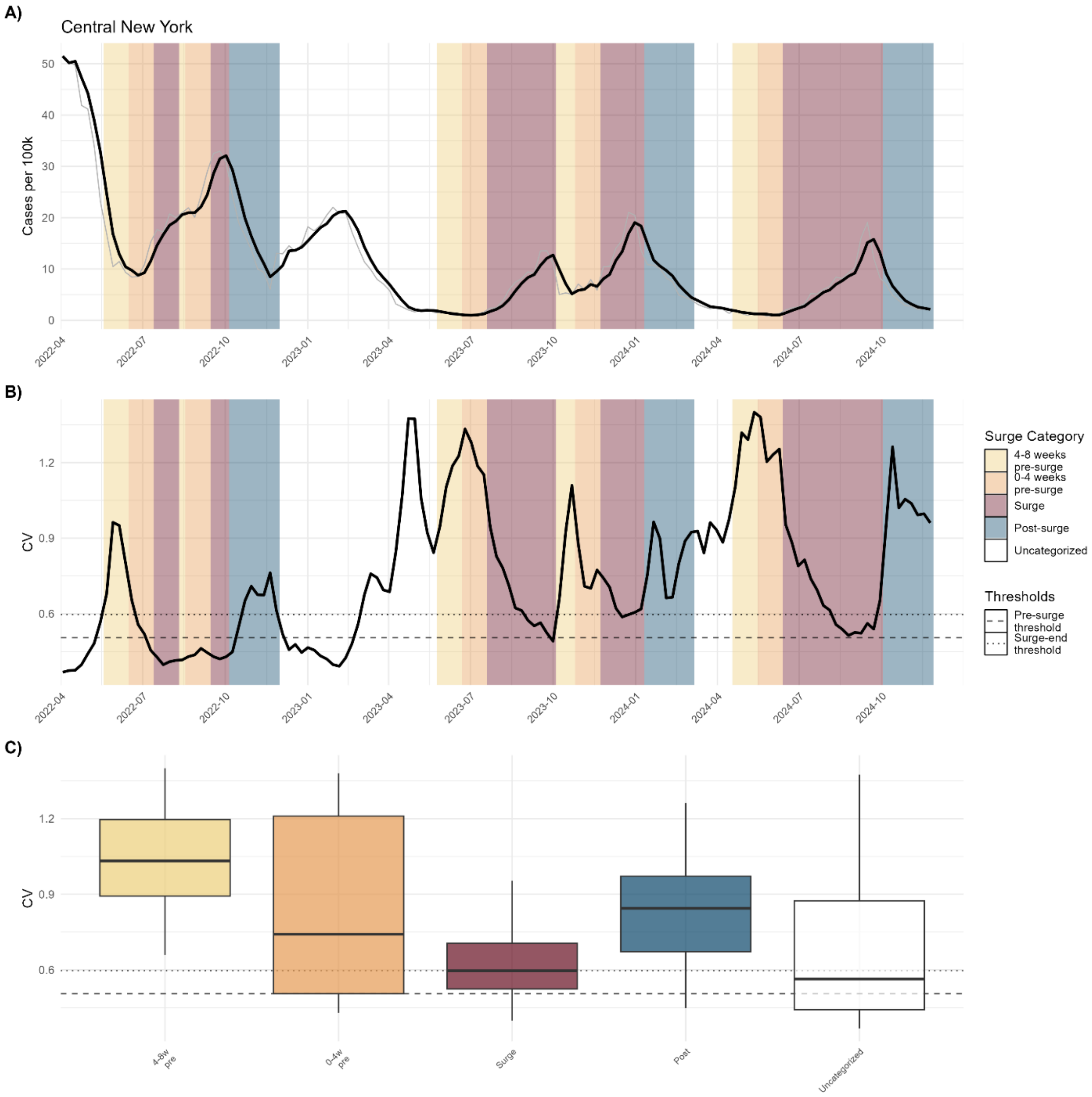


**Figure S16.** Cases CV of Central New York

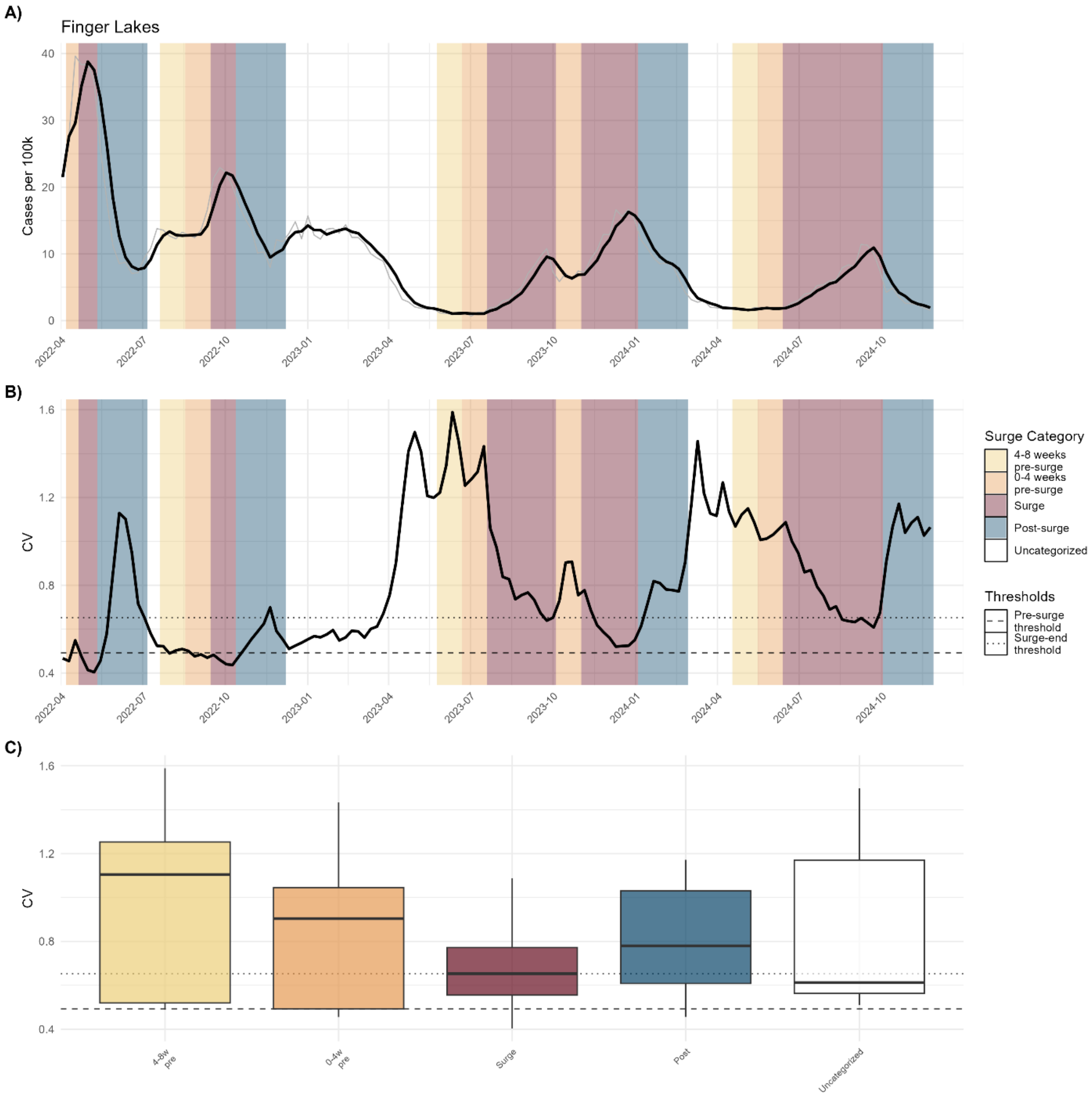


**Figure S17.** Cases CV of Finger Lakes

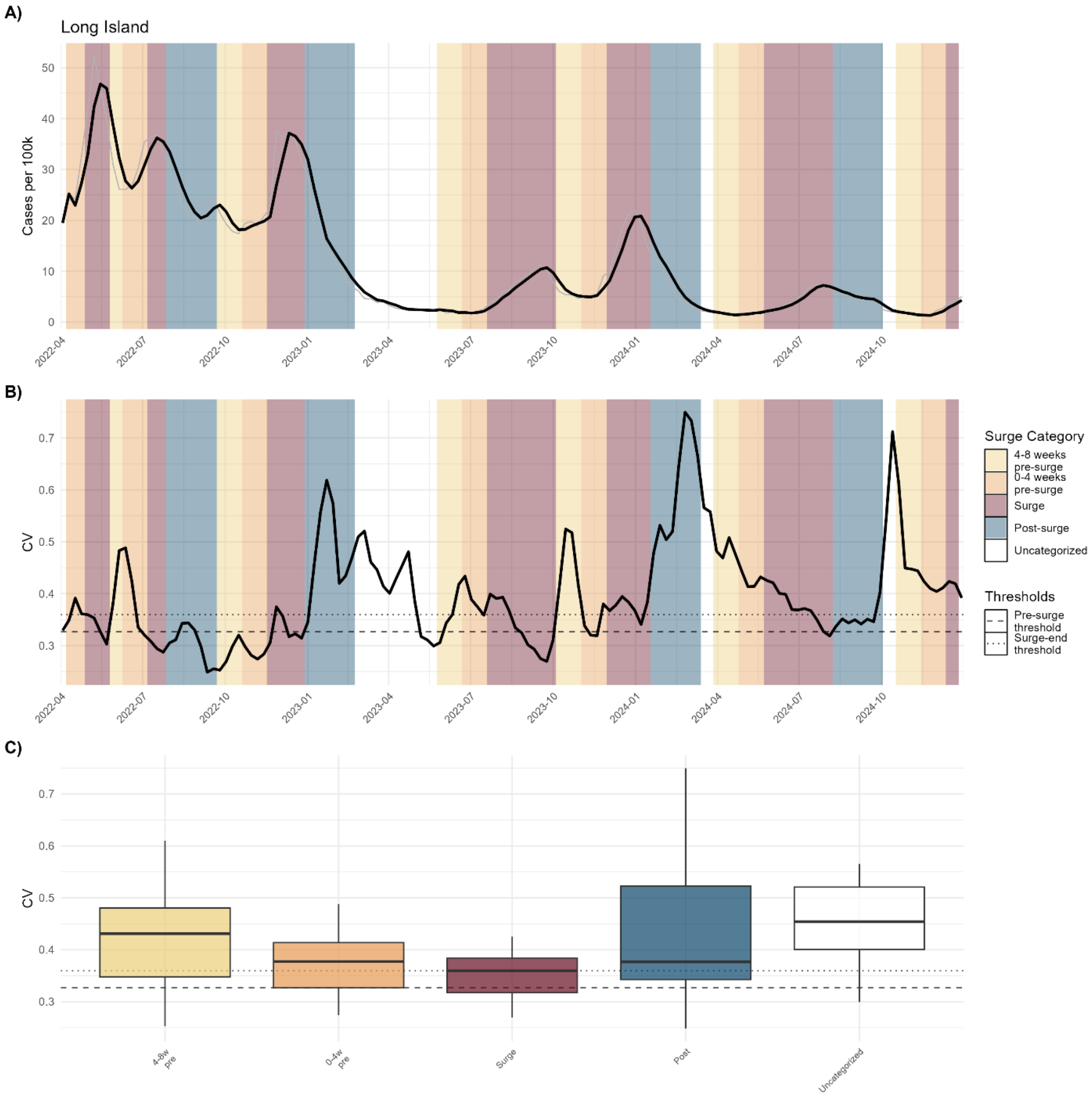


**Figure S18.** Cases CV of Long Island

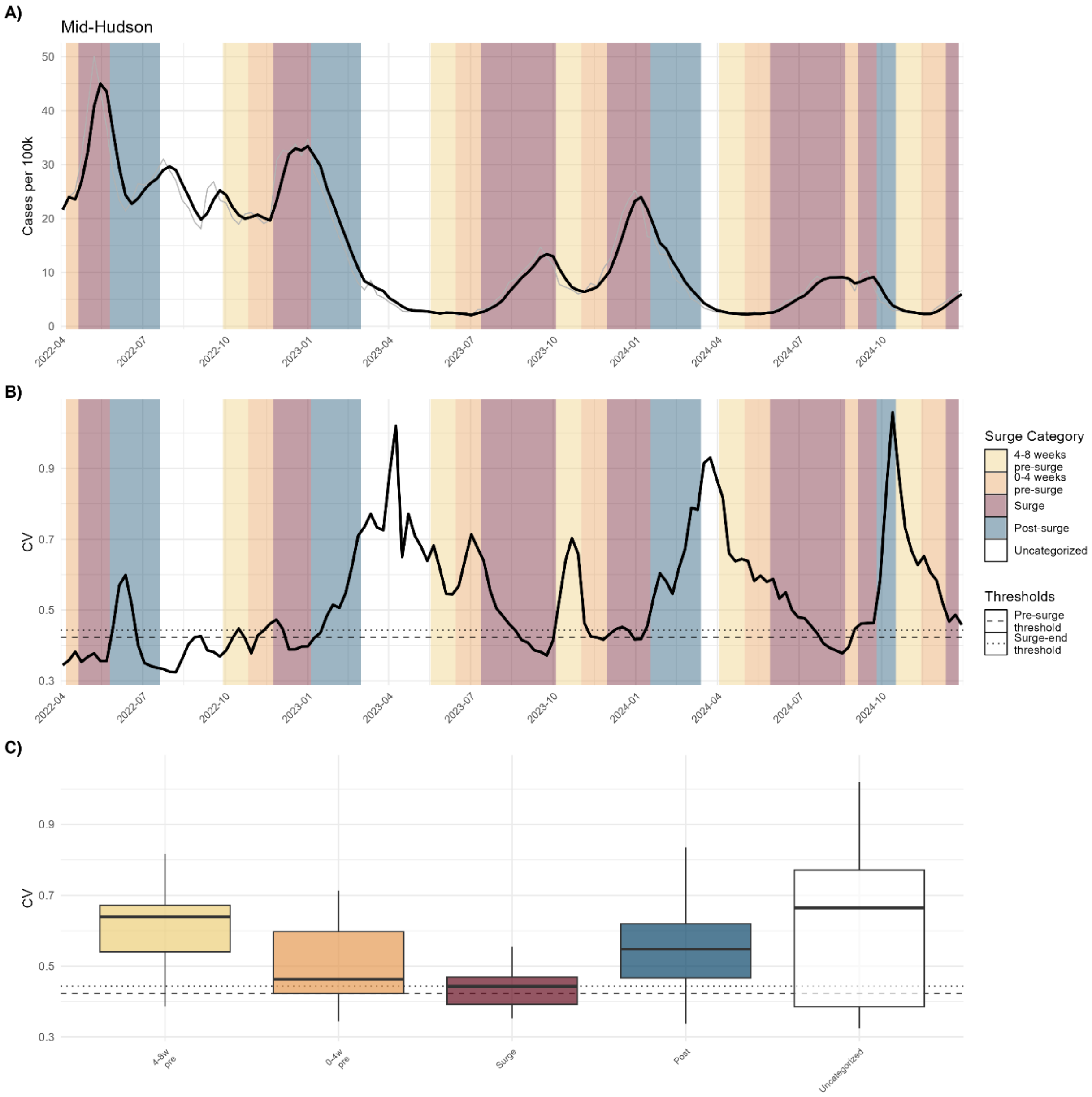


**Figure S19.** Cases CV of Mid-Hudson

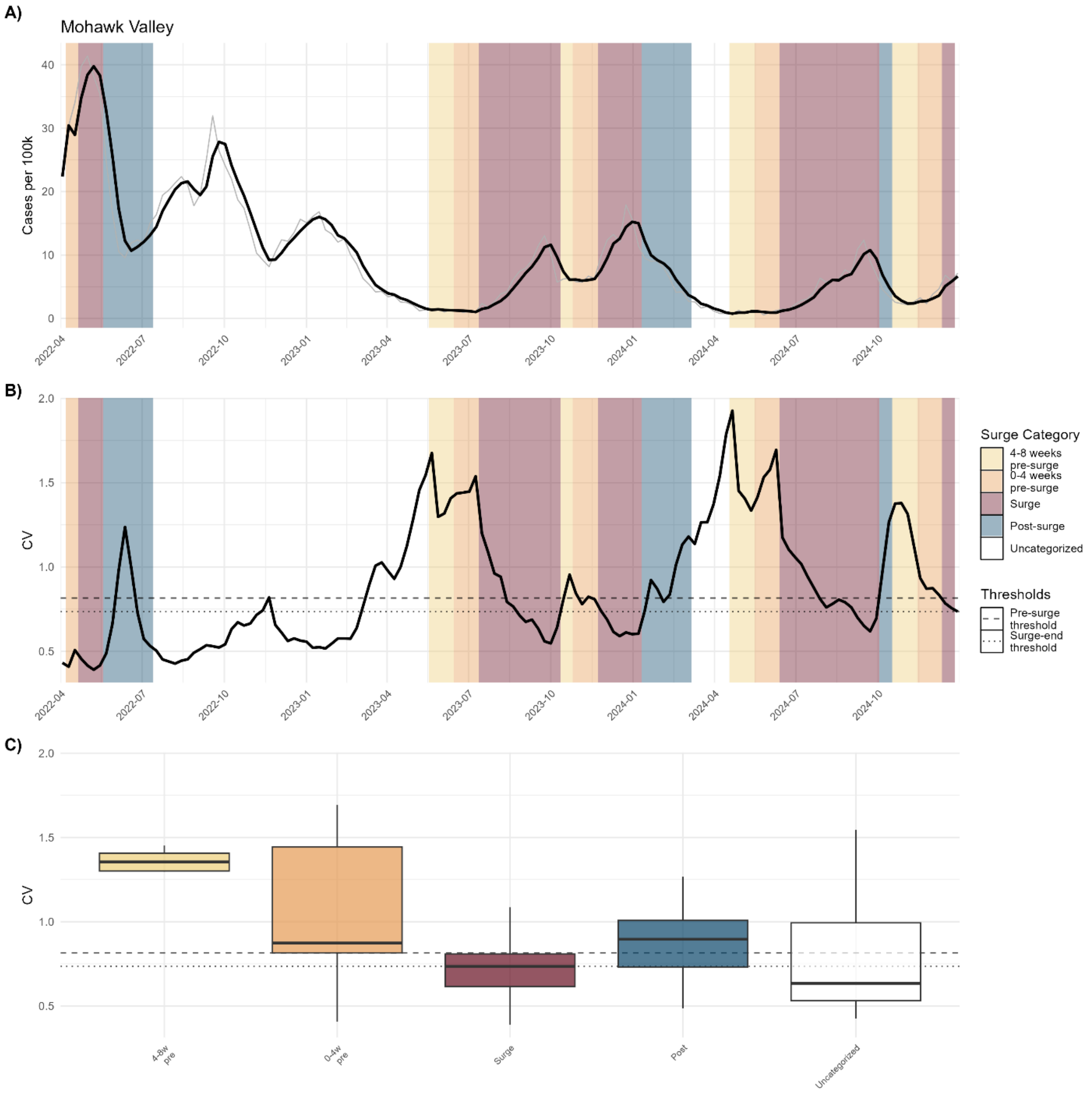


**Figure S20.** Cases CV of Mohawk Valley

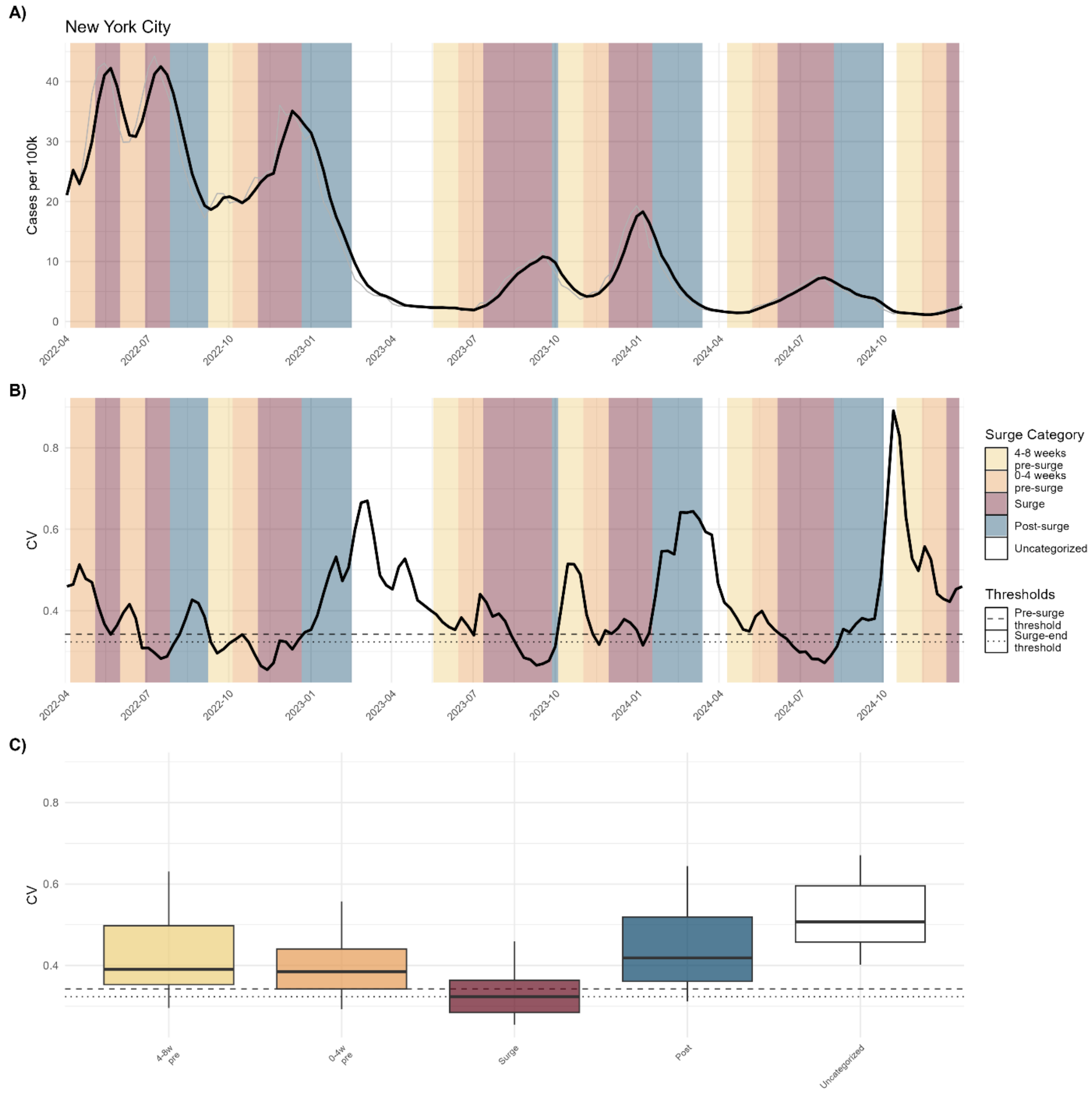


**Figure S21.** Cases CV of New York City

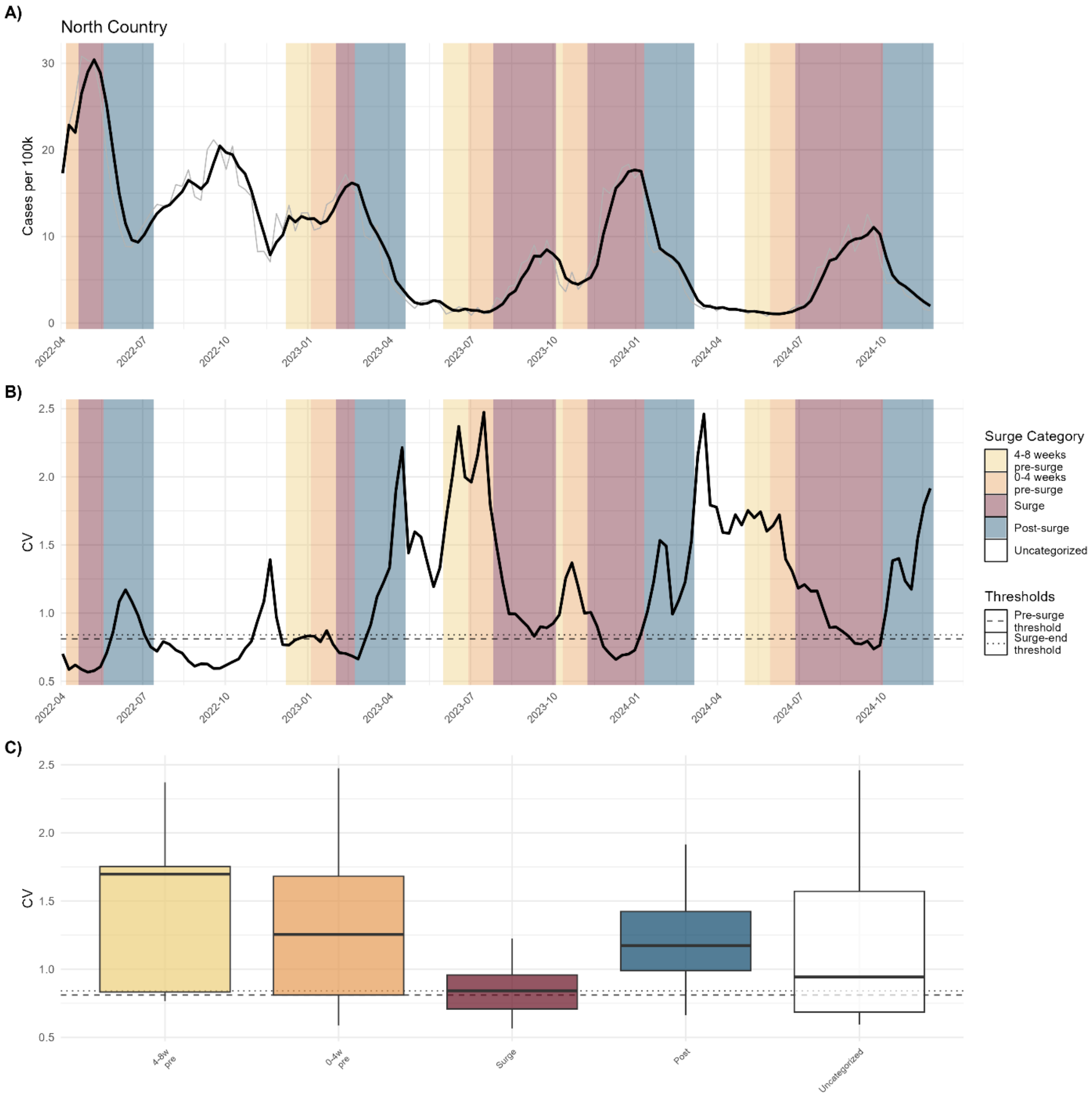


**Figure S22.** Cases CV of North Country

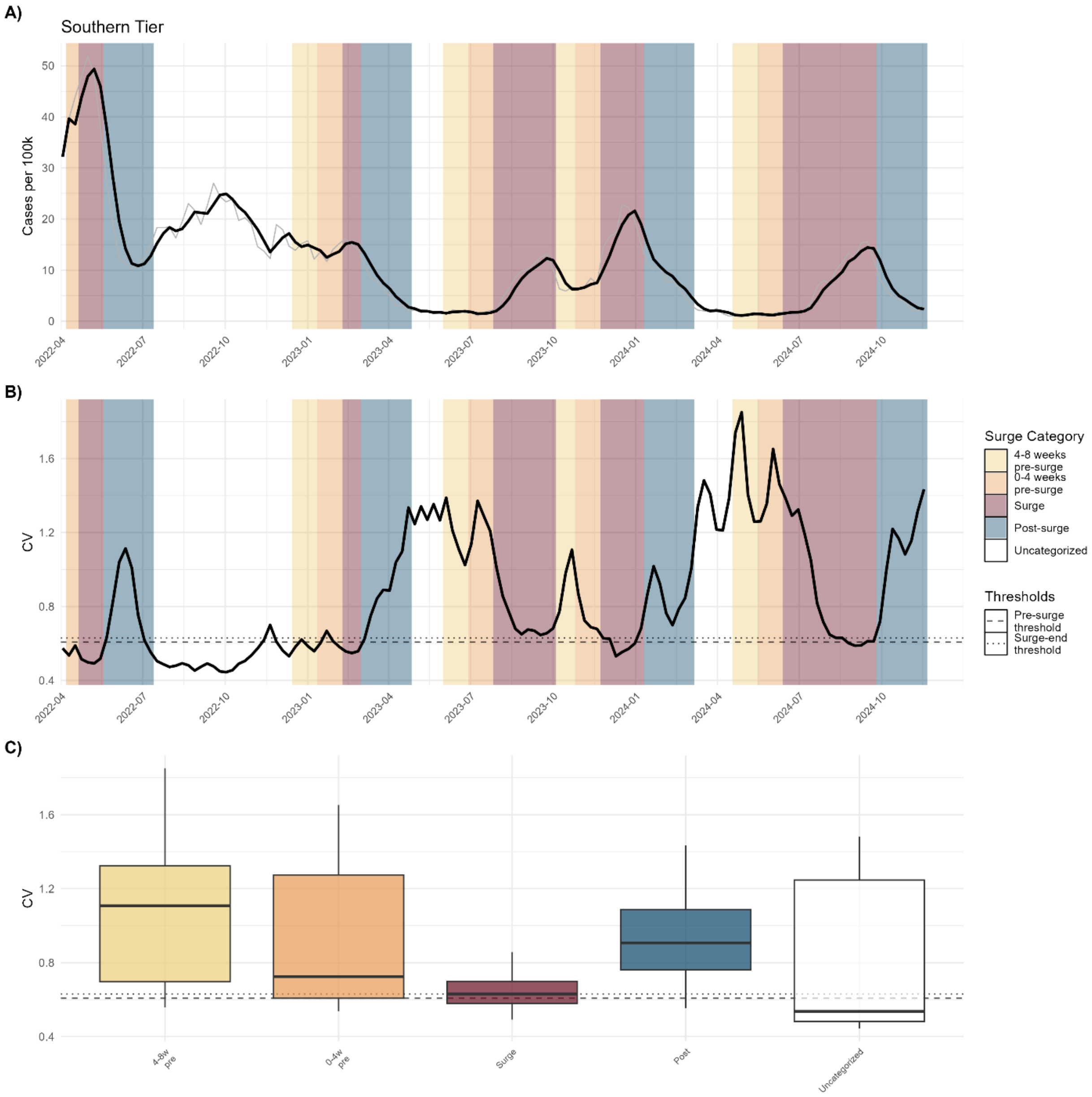


**Figure S23.** Cases CV of Southern Tier

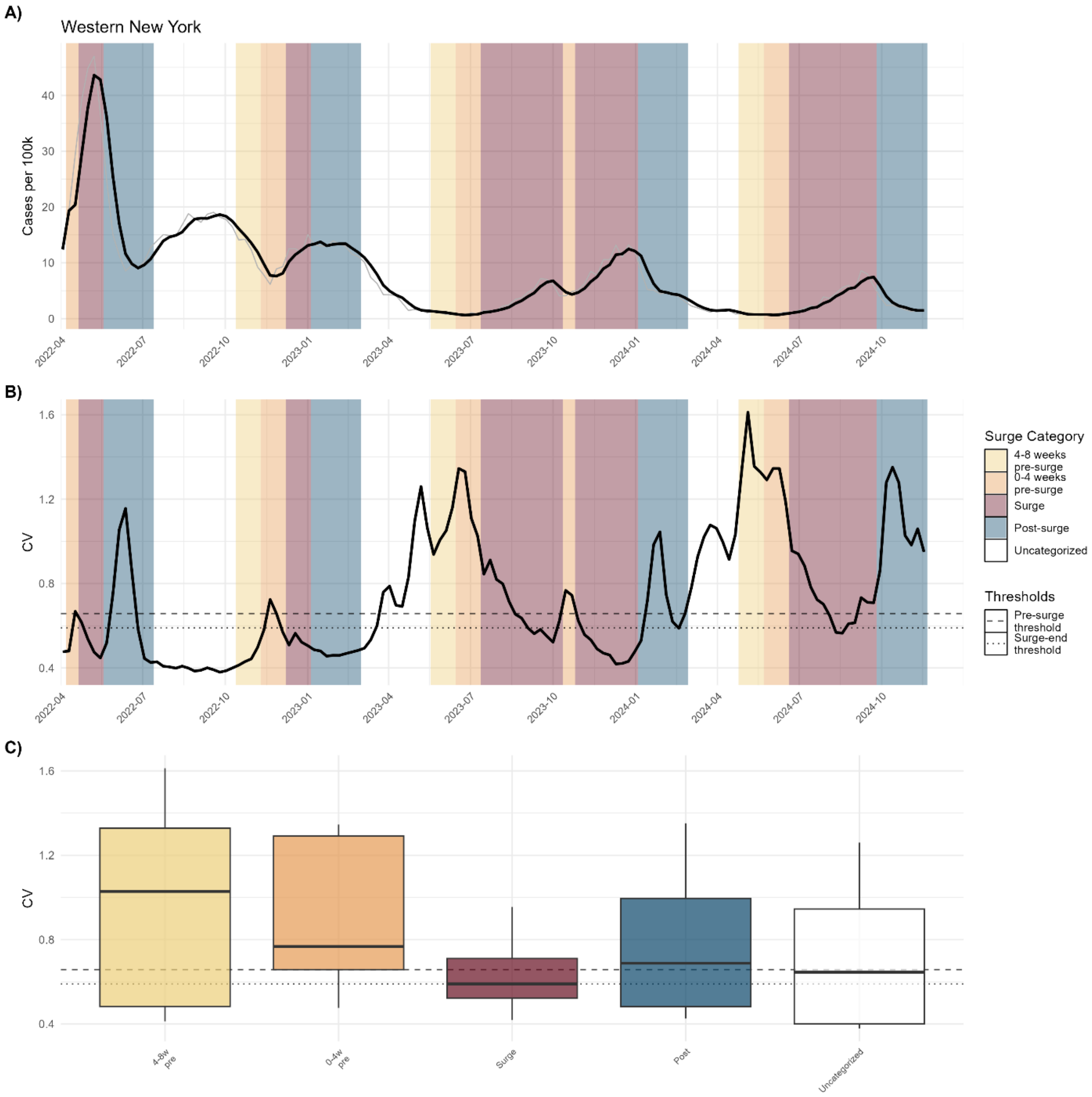


**Figure S24.** Cases CV of Western New York

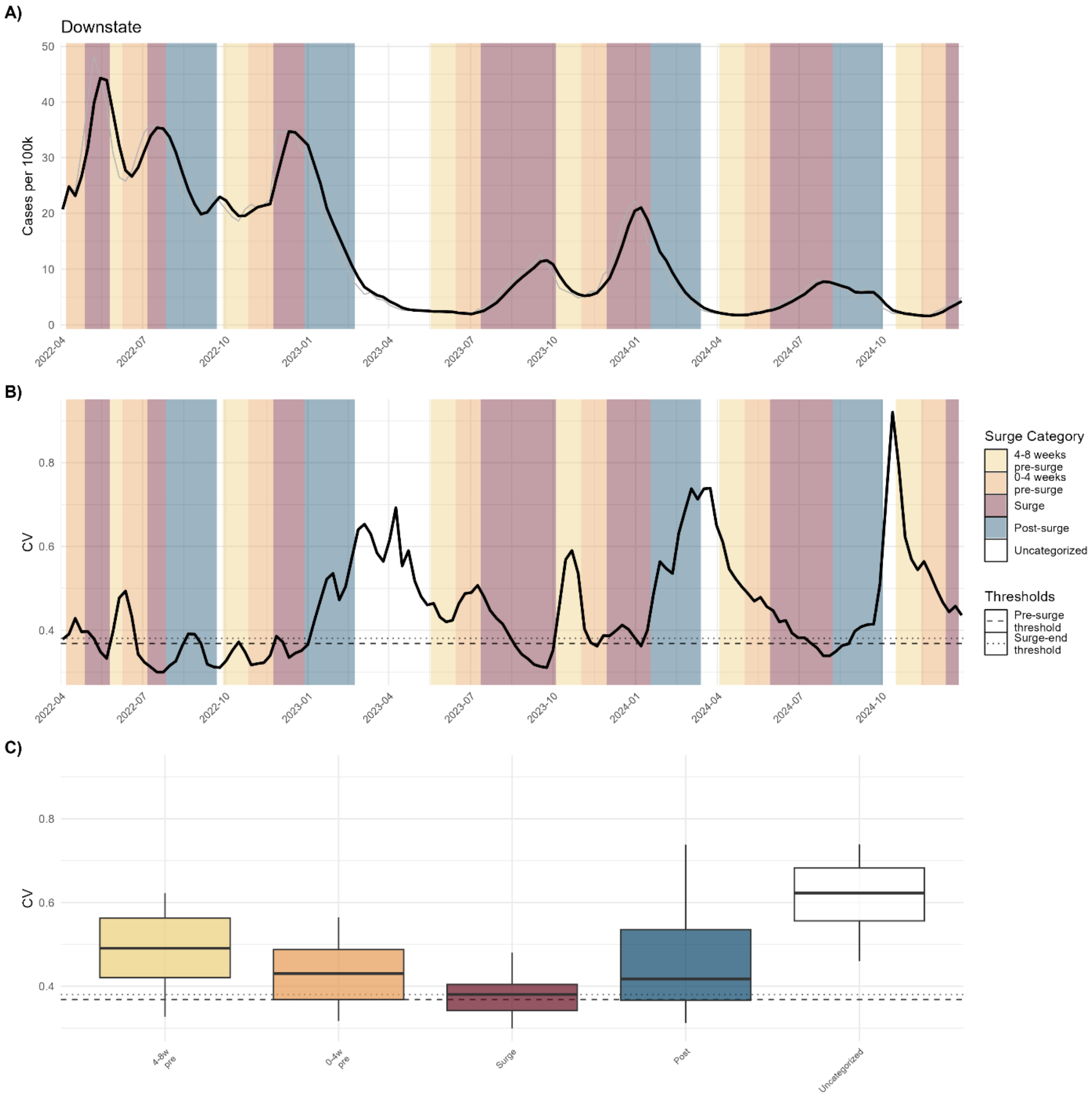


**Figure S25.** Cases CV of Downstate

#### Clinical Cases Kruskal-Wallis Pairwise Comparisons

These tables present pairwise Wilcoxon rank-sum test results comparing coefficient of variation (CV) across adjacent surge categories. Adjacent category comparison includes the two surge categories being compared, the number of weekly observations in each group, the median CV for each group, the unadjusted Wilcoxon rank-sum test p-value, and the Bonferroni-corrected p-value adjusted for multiple comparisons. Green highlighting indicates adjusted $p < 0.05$; orange indicates $0.05 \leq$ adjusted $p < 0.10$.

**Table S6.** CV adjacent comparisons of all regions (clinical)

| Category 1 | Category 2 | $n_1$ (weeks) | $n_2$ (weeks) | Median$_1$ | Median$_2$ | p-value | Adjusted p |
|---|---|---|---|---|---|---|---|
| Uncategorized | 4-8 weeks pre-surge | 357 | 158 | 0.638 | 0.803 | 0.022318 | 0.11159 |
| 4-8 weeks pre-surge | 0-4 weeks pre-surge | 158 | 213 | 0.803 | 0.613 | 0.011269 | 0.05634 |
| 0-4 weeks pre-surge | Surge | 213 | 434 | 0.613 | 0.565 | 0.000266 | 0.00133 |
| Surge | Post-surge | 434 | 278 | 0.565 | 0.710 | 8.18e-12 | 4.09e-11 |
| Post-surge | Uncategorized | 278 | 357 | 0.710 | 0.638 | 0.328618 | 1.00000 |

**Table S7.** CV adjacent comparisons of downstate (clinical)

| Category 1 | Category 2 | $n_1$ (weeks) | $n_2$ (weeks) | Median 1 | Median 2 | p-value | Adjusted p |
|---|---|---|---|---|---|---|---|
| Uncategorized | 4-8 weeks pre-surge | 18 | 22 | 0.623 | 0.491 | 0.00184 | 0.009200 |
| 4-8 weeks pre-surge | 0-4 weeks pre-surge | 22 | 28 | 0.491 | 0.431 | 0.01855 | 0.092758 |
| 0-4 weeks pre-surge | Surge | 28 | 44 | 0.431 | 0.381 | 0.00426 | 0.021313 |
| Surge | Post-surge | 44 | 32 | 0.381 | 0.418 | 0.00105 | 0.005229 |
| Post-surge | Uncategorized | 32 | 18 | 0.418 | 0.623 | 0.00019 | 0.000952 |

**Table S8.** CV adjacent comparisons of upstate (clinical)

| Category 1 | Category 2 | $n_1$ (weeks) | $n_2$ (weeks) | Median 1 | Median 2 | p-value | Adjusted p |
|---|---|---|---|---|---|---|---|
| Uncategorized | 4-8 weeks pre-surge | 56 | 10 | 0.589 | 1.364 | 4.38e-05 | 0.000219 |
| 4-8 weeks pre-surge | 0-4 weeks pre-surge | 10 | 15 | 1.364 | 1.288 | 0.11513 | 0.575668 |
| 0-4 weeks pre-surge | Surge | 15 | 39 | 1.288 | 0.661 | 0.00374 | 0.018702 |
| Surge | Post-surge | 39 | 24 | 0.661 | 0.960 | 9.00e-05 | 0.000450 |
| Post-surge | Uncategorized | 24 | 56 | 0.960 | 0.589 | 0.00585 | 0.029257 |

### Supplemental Figures: Hospitalizations

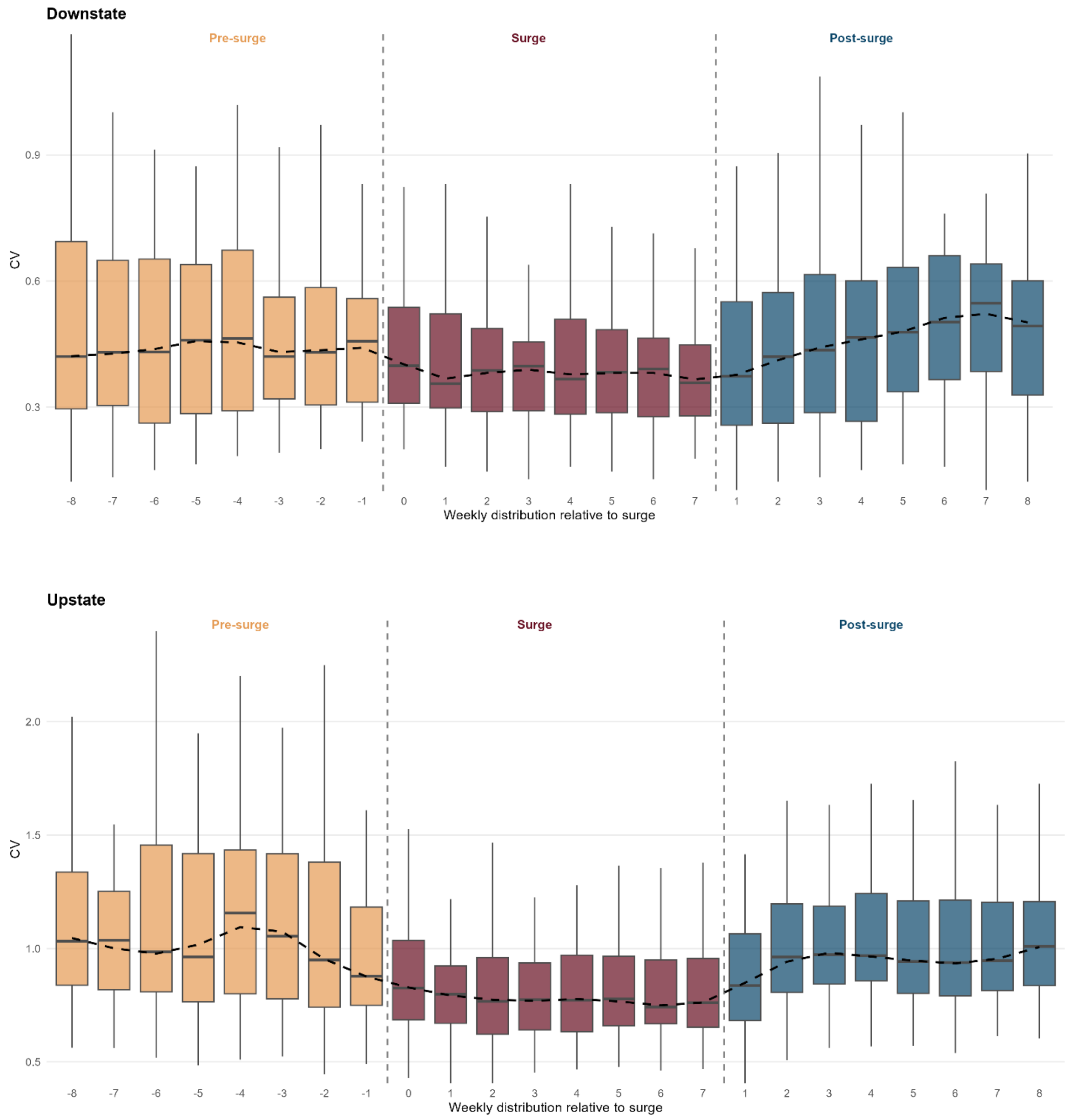


**Figure S26. Distribution of coefficient of variation (CV) by week relative to surge onset and termination for hospitalizations for Downstate (top) and Upstate (bottom).** Boxplots show the distribution of CV values at each weekly position, pooled across all surges and regions, with weeks aligned relative to surge onset (left dashed line) and surge termination (right dashed line). Surge duration varies across events; the surge panel displays weeks 0 through 7 as a representative range encompassing observed surge lengths. Pre-surge weeks show elevated CV values peaking approximately 4 weeks before surge onset, followed by a decline into the surge. Surge weeks exhibit lower CV with a declining trend through active transmission. Post-surge weeks show a transient rise in CV during early fadeout before stabilizing. The dashed black line traces the median CV across weekly positions. This pattern illustrates the full bidirectional early warning signals present on either side of the surge.

Figures below show: Coefficient of variation analysis for hospitalizations. Panels show: **A)** clinical case time series raw values (grey) with 3-week smoothing (black), **B)** coefficient of variation over time with 25th percentile threshold of pre-surge periods (dashed) and 50th percentile threshold of surge periods (dotted), and **C)** distribution of CV values by surge category and threshold lines. Background colors indicate epidemic phases, as shown in the legend.

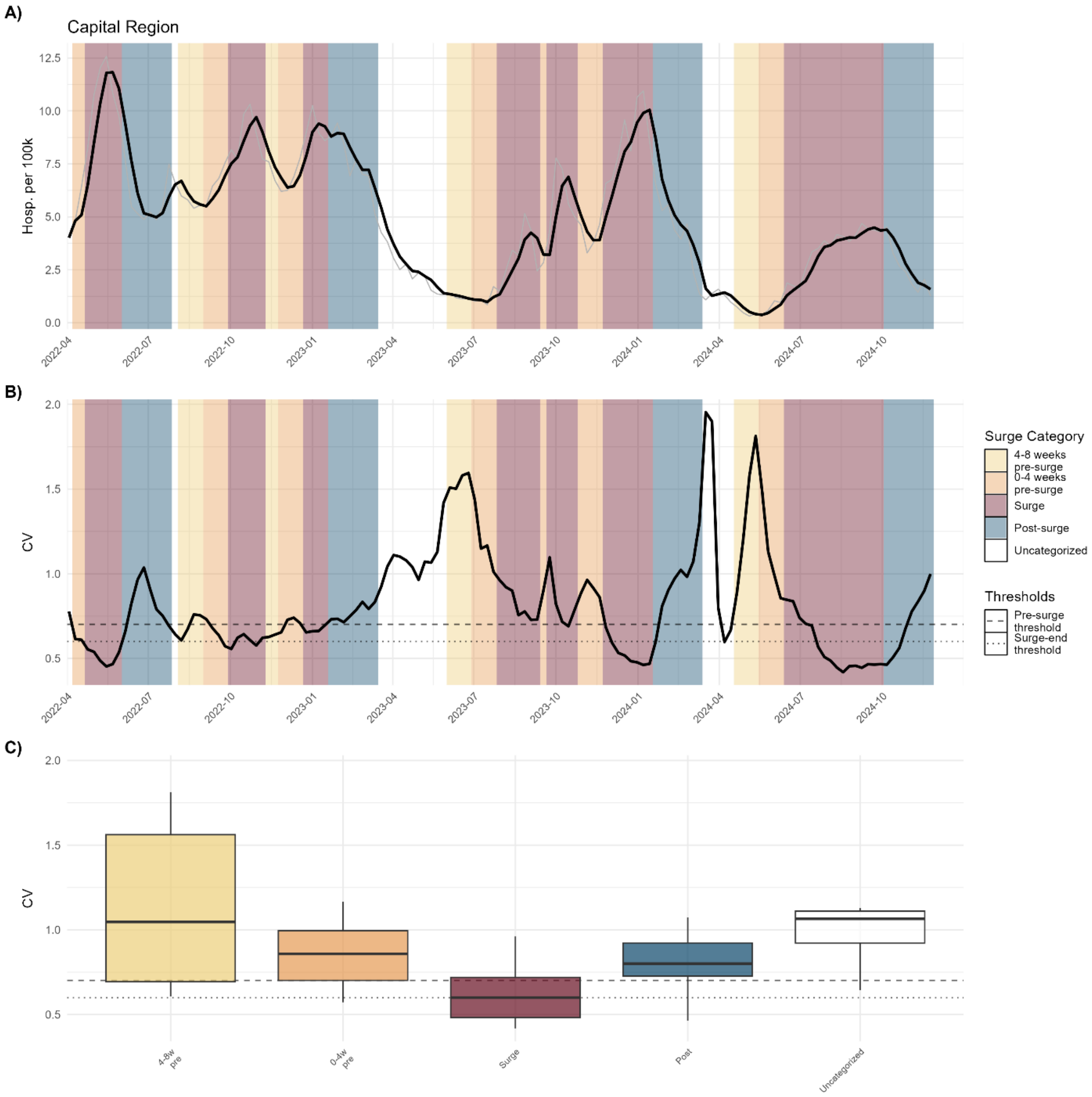


**Figure S27.** Hospitalizations CV of Capital Region

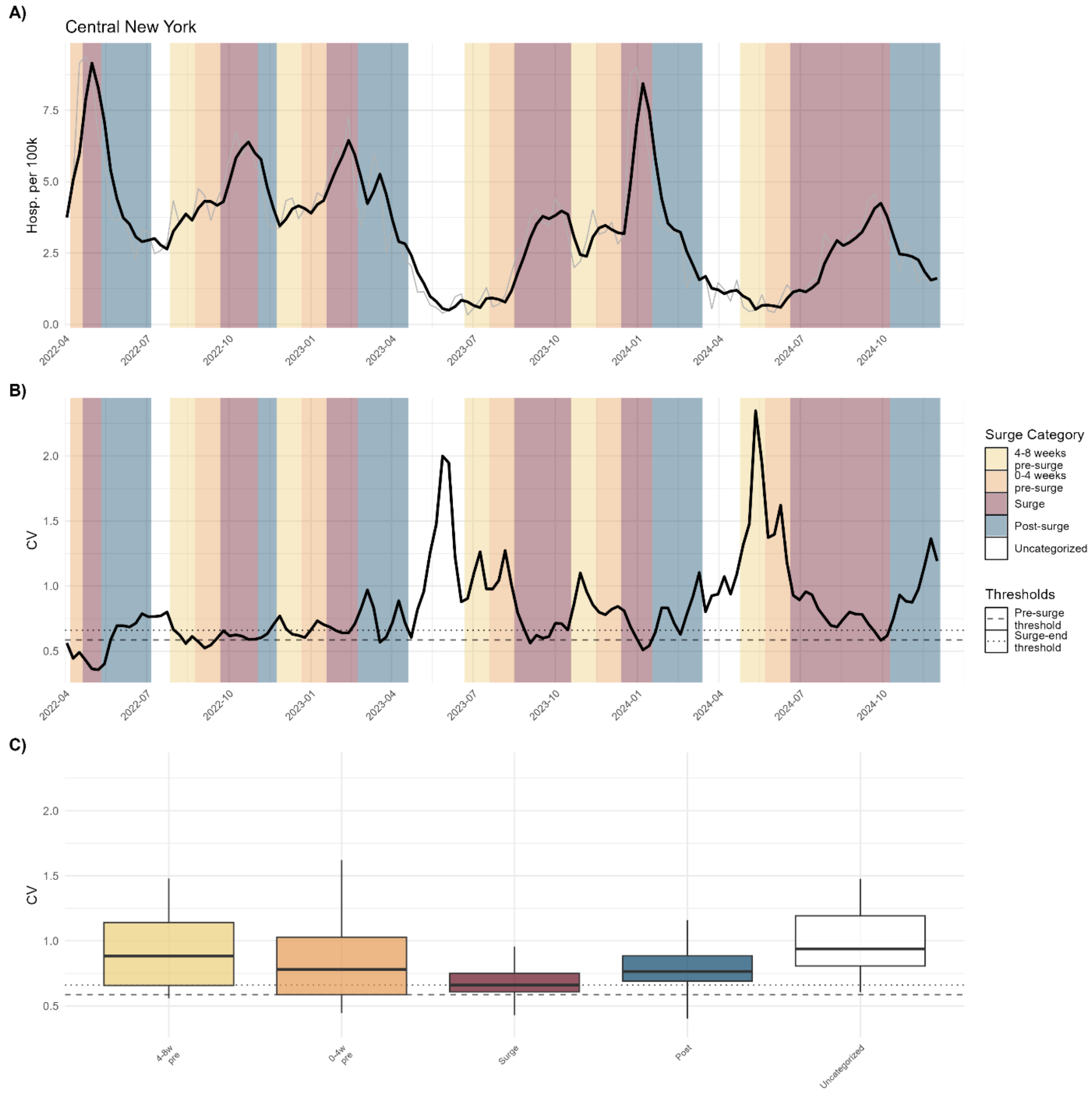


**Figure S28.** Hospitalizations CV of Central New York

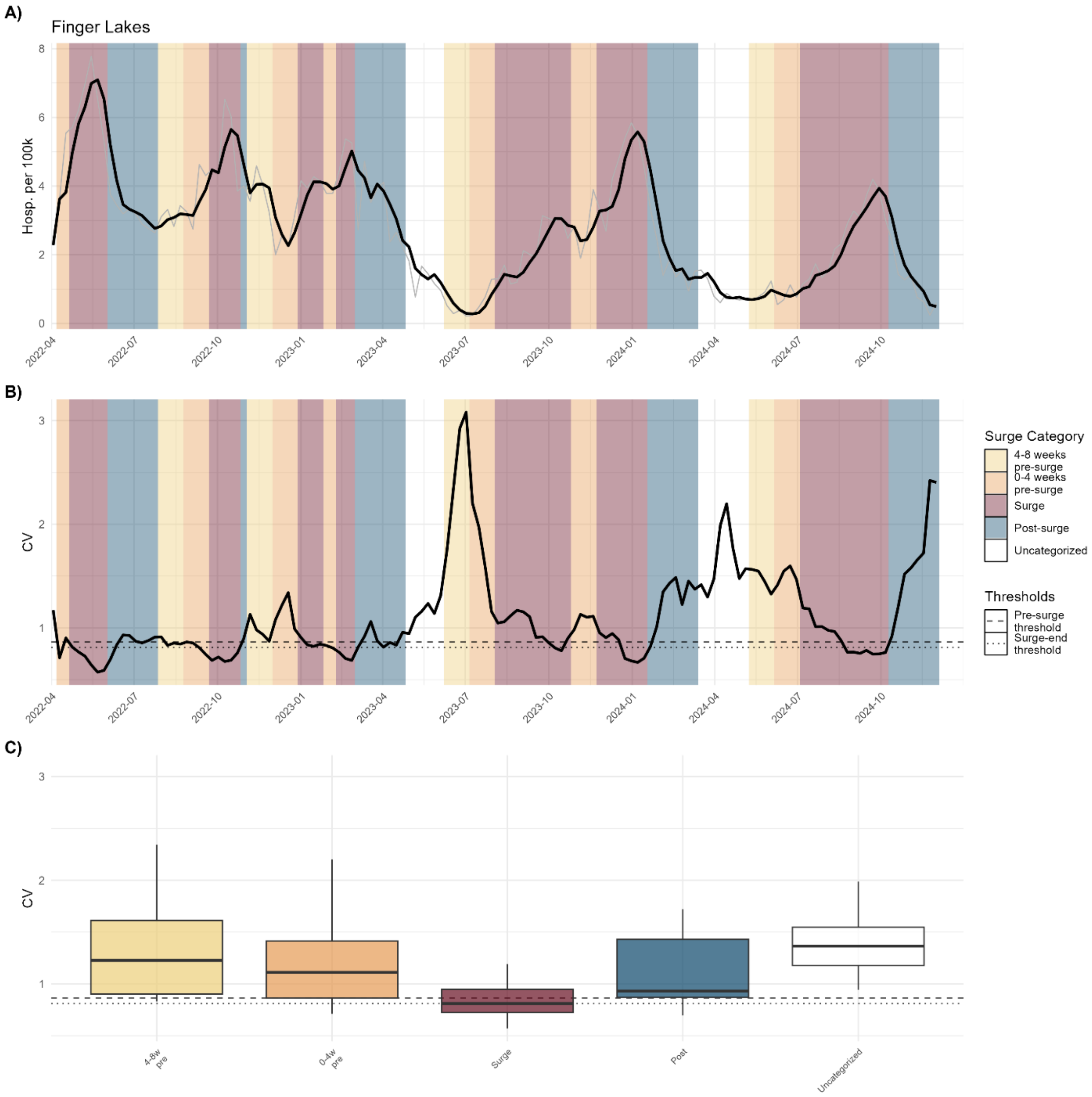


**Figure S29.** Hospitalizations CV of Finger Lakes

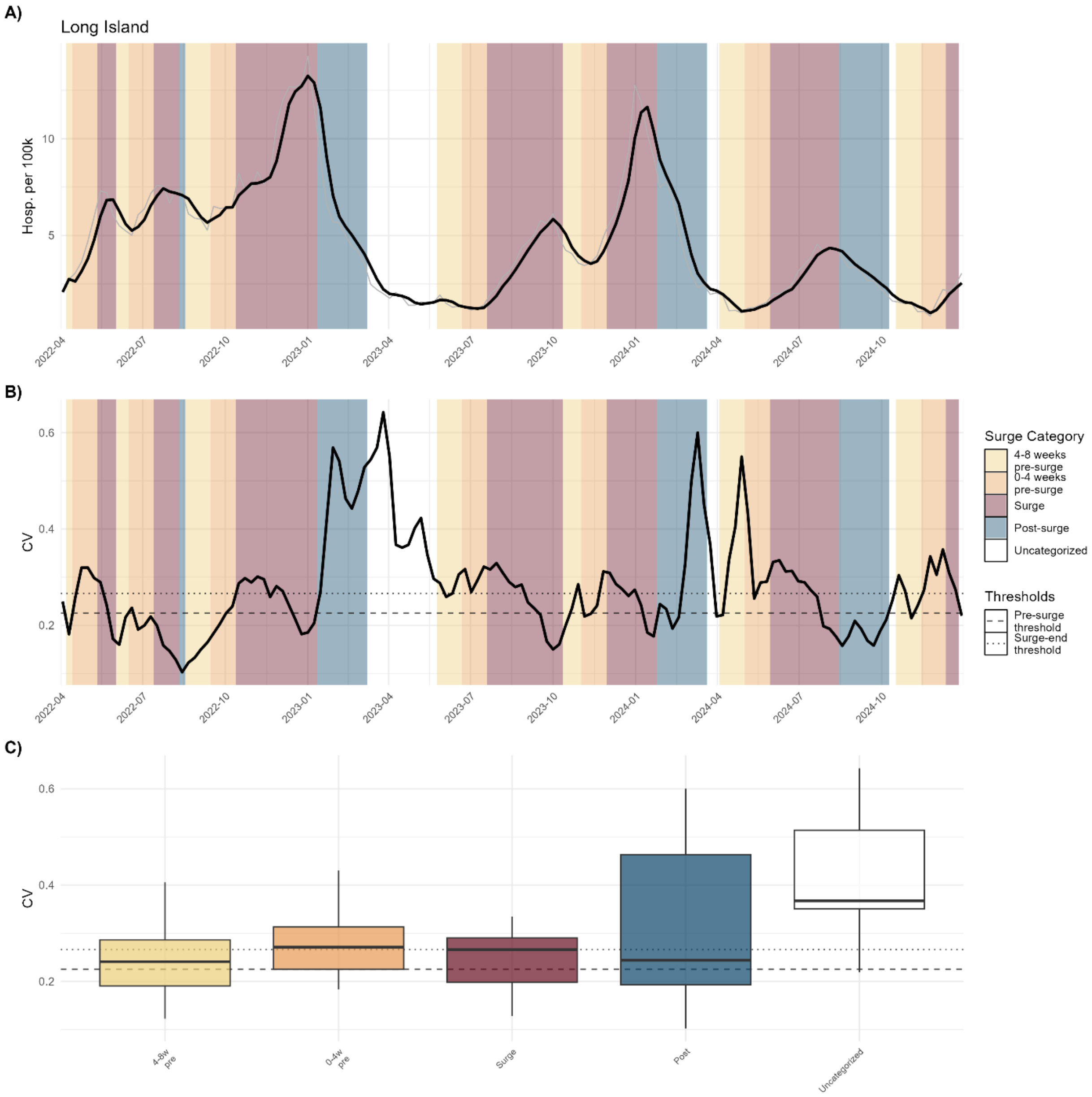


**Figure S30.** Hospitalizations CV of Long Island

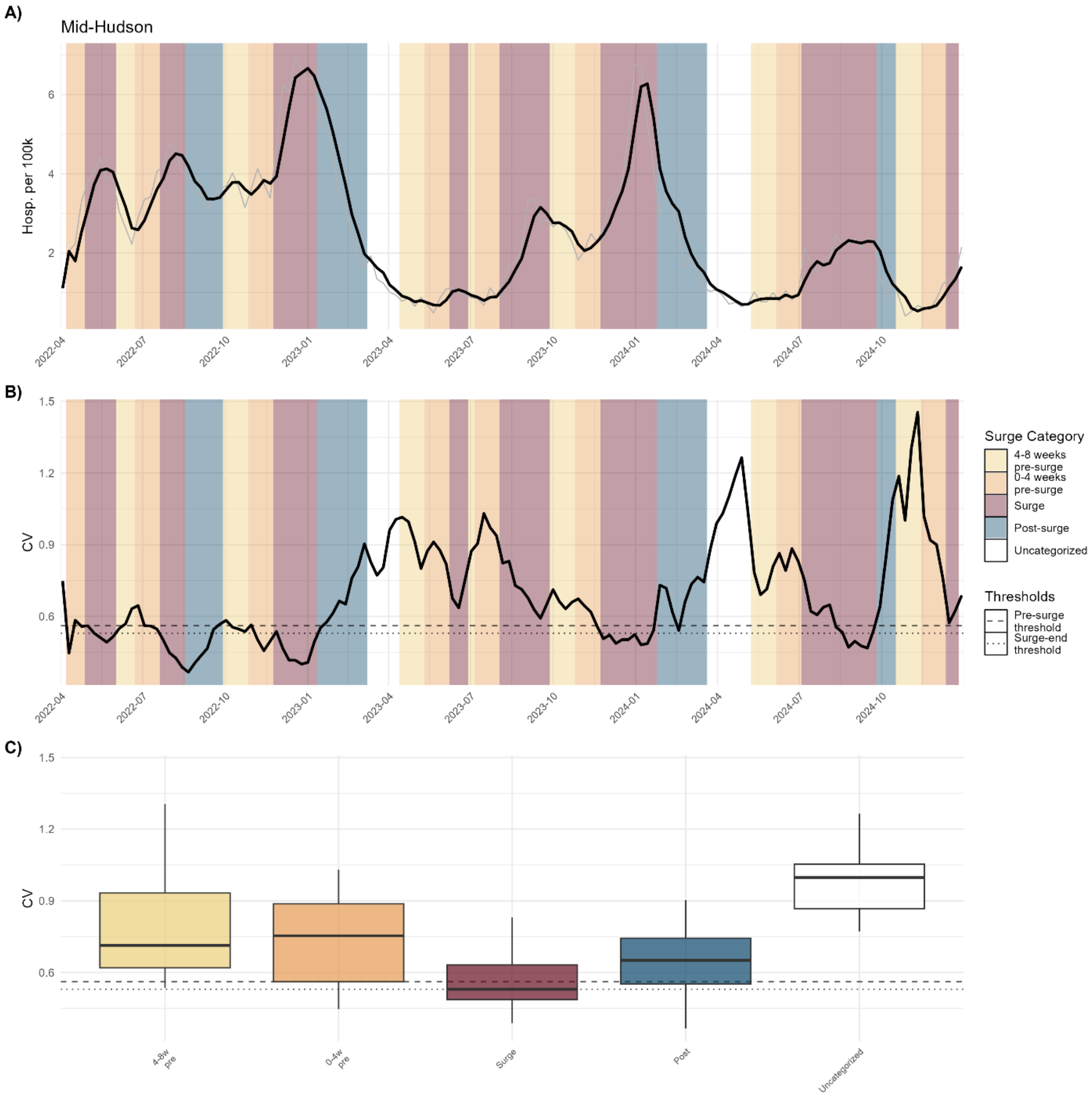


**Figure S31.** Hospitalizations CV of Mid-Hudson

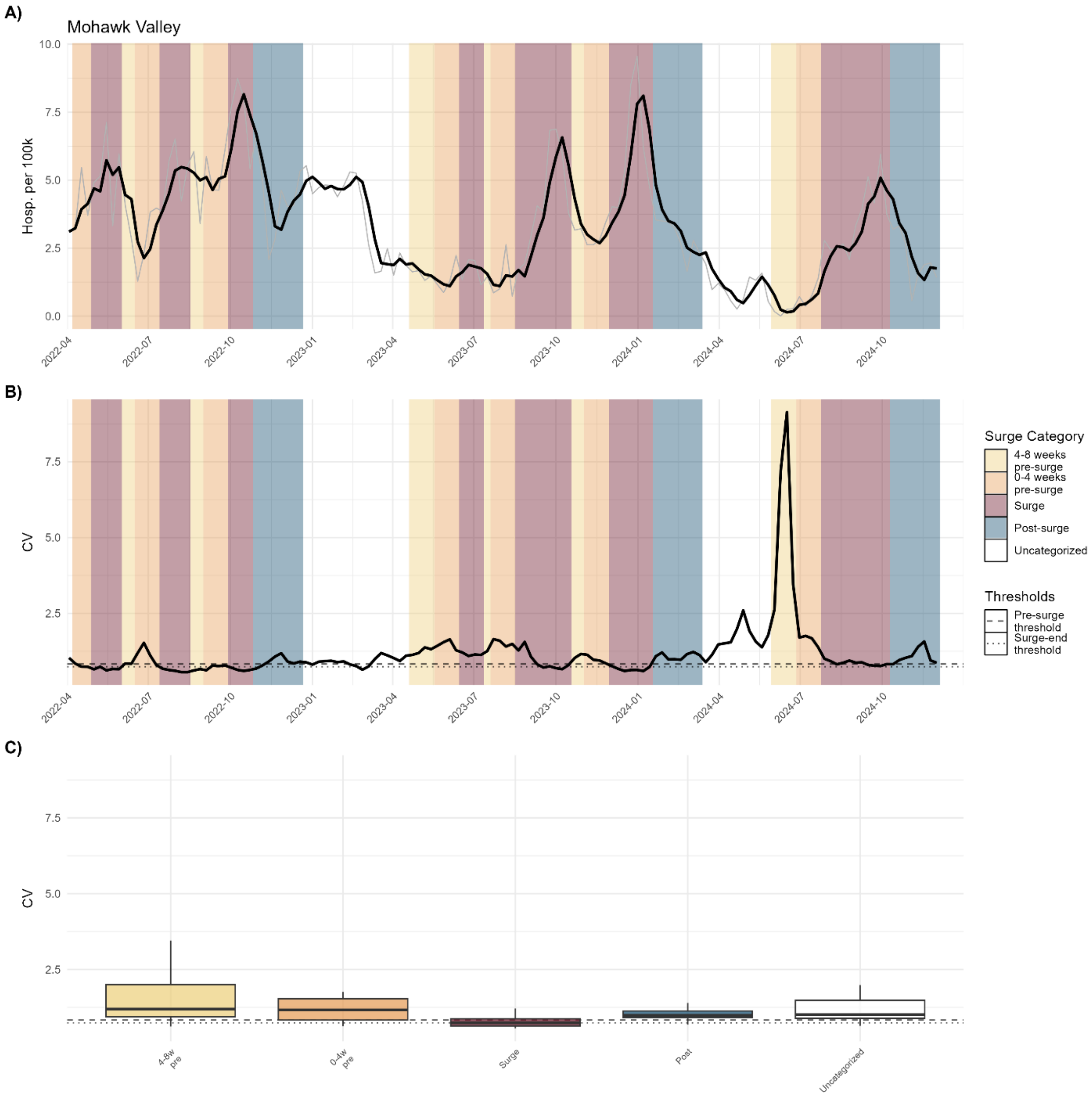


**Figure S32.** Hospitalizations CV of Mohawk Valley

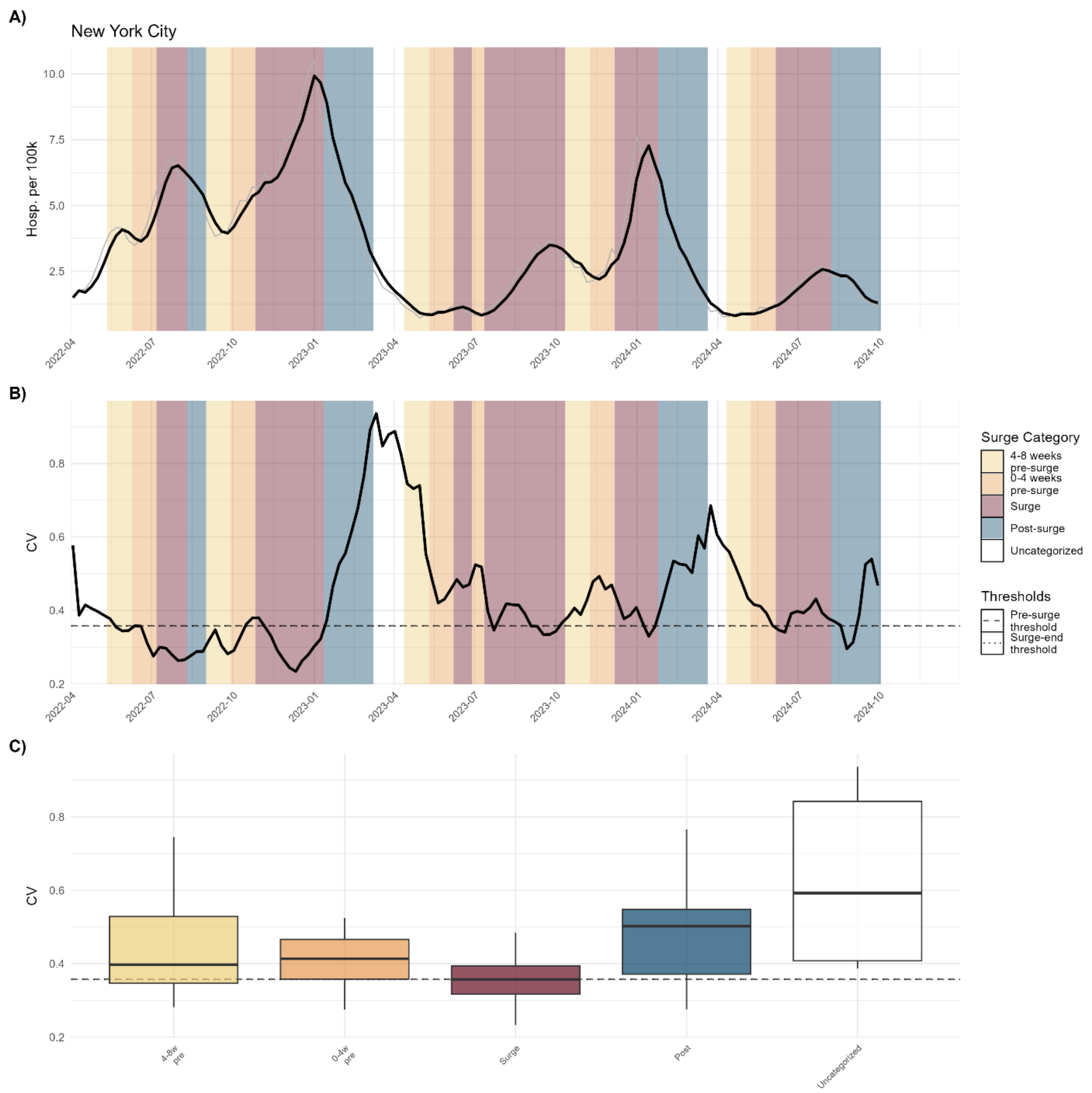


**Figure S33.** Hospitalizations CV of New York City

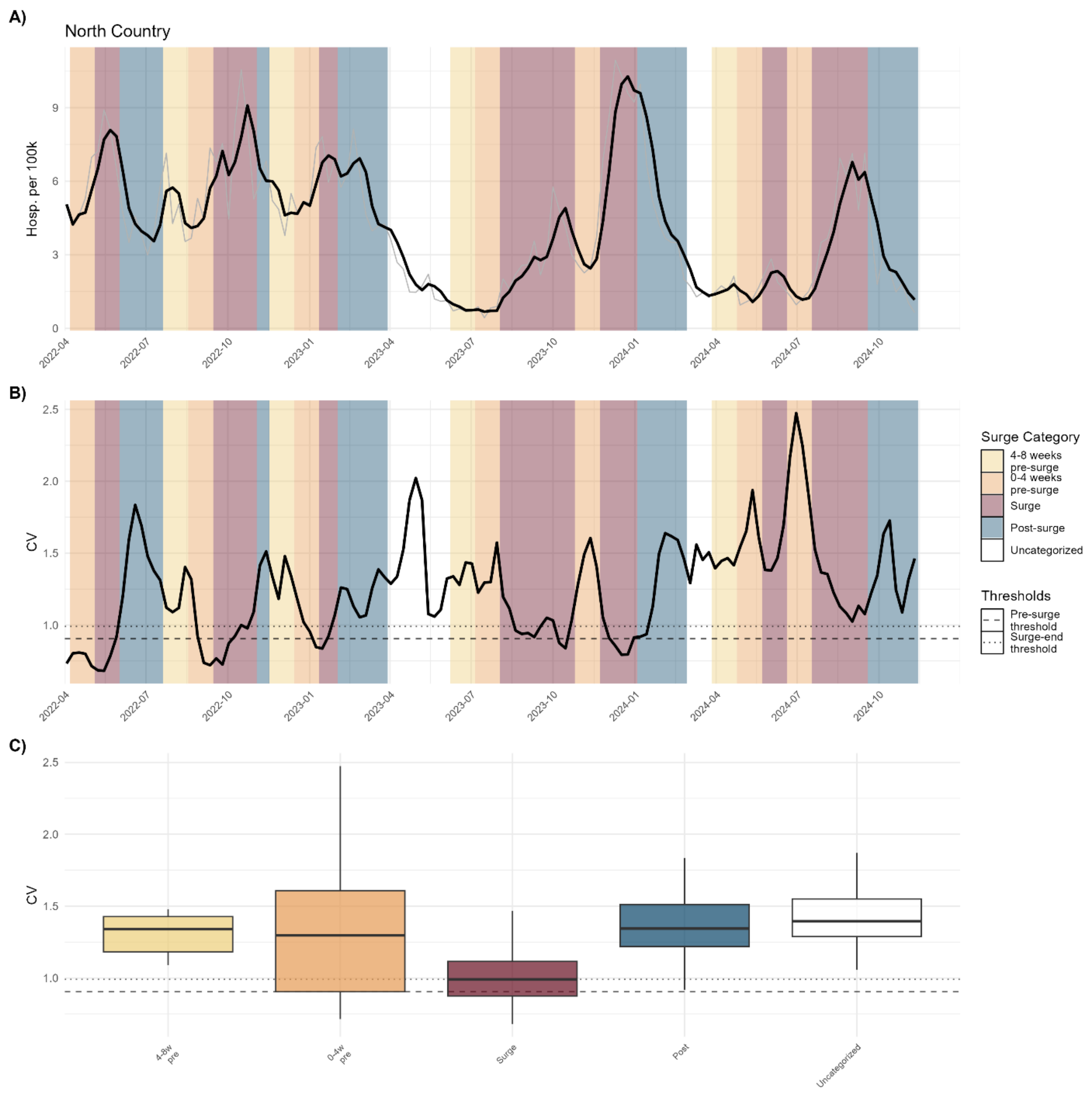


**Figure S34.** Hospitalizations CV of North Country

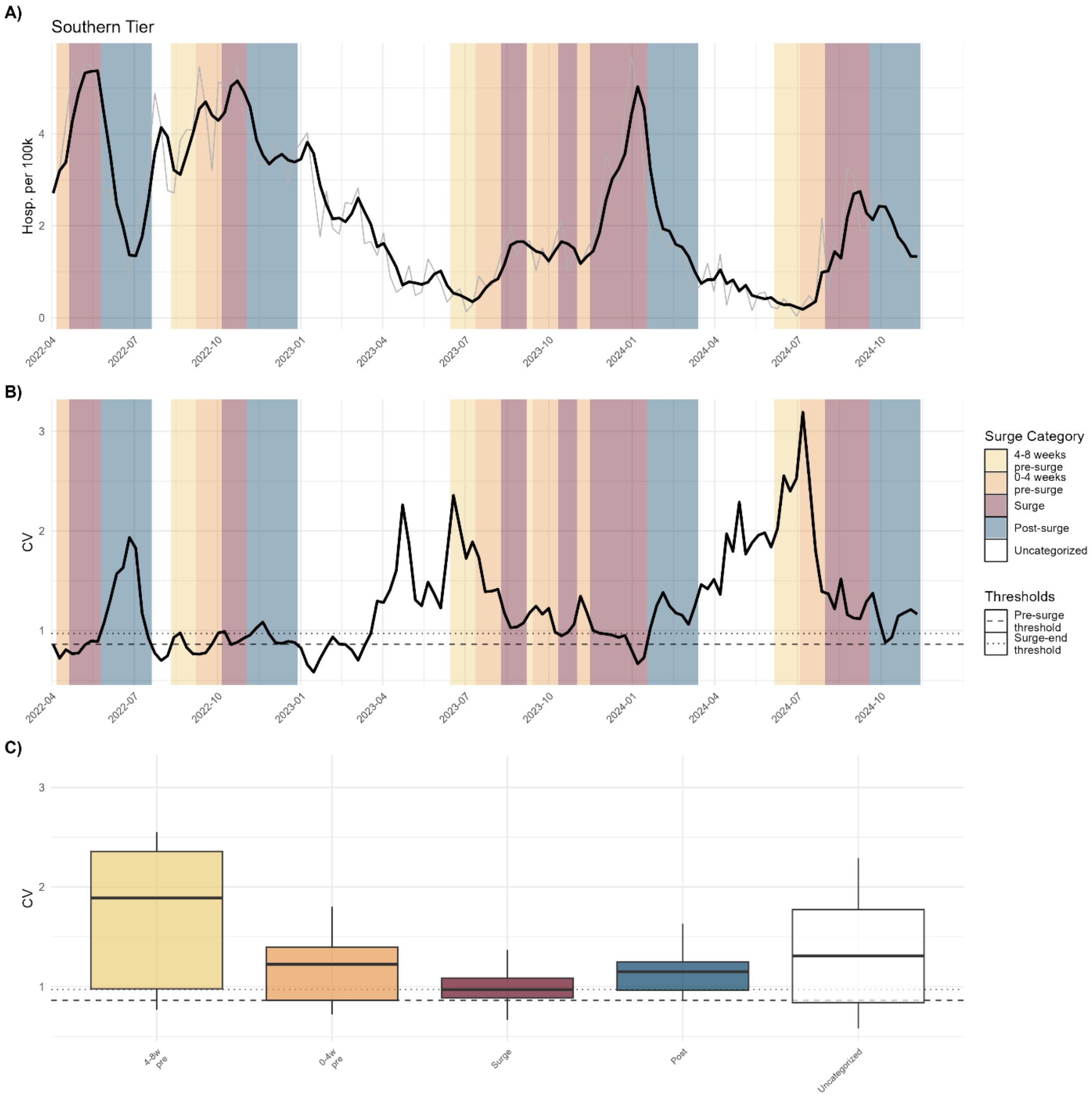


**Figure S35.** Hospitalizations CV of Southern Tier

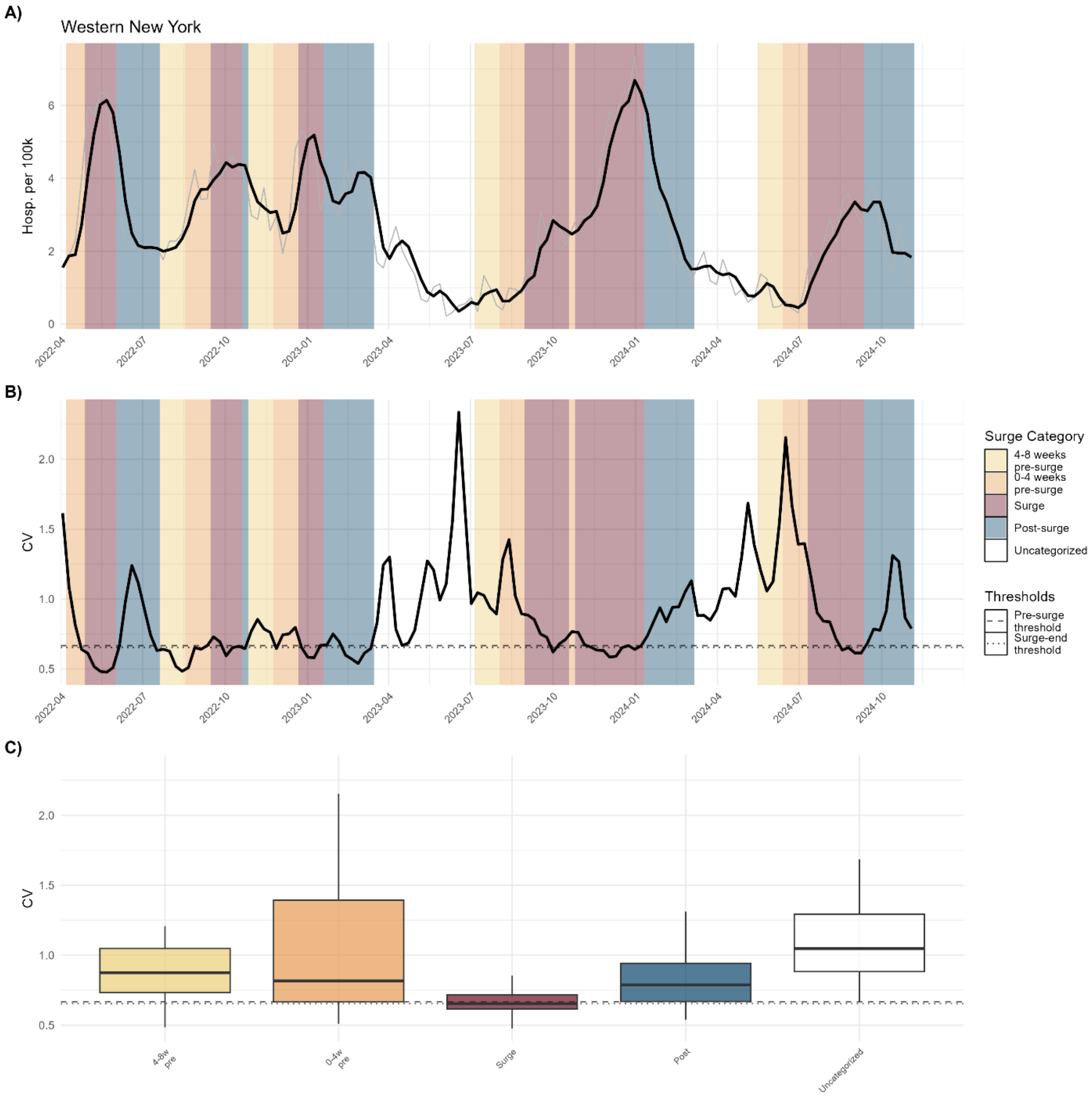


**Figure S36.** Hospitalizations CV of Western New York

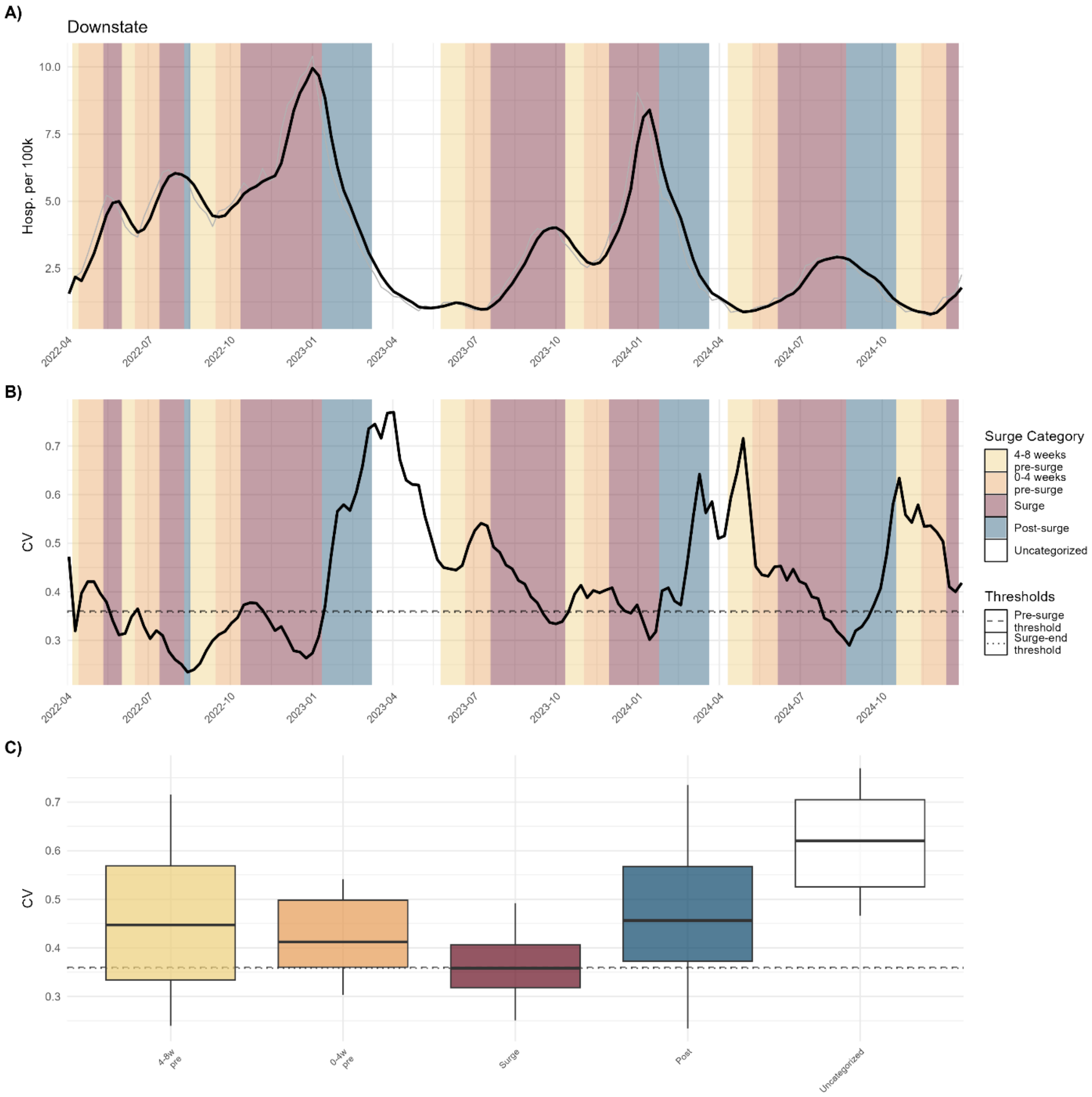


**Figure S37.** Hospitalizations CV of Downstate

#### Hospitalizations Kruskal-Wallis Pairwise Comparisons

These tables present pairwise Wilcoxon rank-sum test results comparing coefficient of variation (CV) across adjacent surge categories. Adjacent category comparison includes the two surge categories being compared, the number of weekly observations in each group, the median CV for each group, the unadjusted Wilcoxon rank-sum test p-value, and the Bonferroni-corrected p-value adjusted for multiple comparisons. Green highlighting indicates adjusted $p < 0.05$; orange indicates $0.05 \le$ adjusted $p < 0.10$.

**Table S9.** CV adjacent comparisons of all regions (hospitalizations)

| Category 1 | Category 2 | $n_1$ (weeks) | $n_2$ (weeks) | Median 1 | Median 2 | p-value | Adjusted p |
|---|---|---|---|---|---|---|---|
| Uncategorized | 4-8 weeks pre-surge | 248 | 178 | 1.013 | 0.848 | 0.000963 | 0.00481 |
| 4-8 weeks pre-surge | 0-4 weeks pre-surge | 178 | 252 | 0.848 | 0.825 | 0.752955 | 1.00000 |
| 0-4 weeks pre-surge | Surge | 252 | 464 | 0.825 | 0.656 | 1.33e-11 | 6.63e-11 |
| Surge | Post-surge | 464 | 298 | 0.656 | 0.858 | < 2e-16 | 2.57e-16 |
| Post-surge | Uncategorized | 298 | 248 | 0.858 | 1.013 | 5.49e-07 | 2.74e-06 |

**Table S10.** CV adjacent comparisons of downstate (hospitalizations)

| Category 1 | Category 2 | $n_1$ (weeks) | $n_2$ (weeks) | Median 1 | Median 2 | p-value | Adjusted p |
|---|---|---|---|---|---|---|---|
| Uncategorized | 4-8 weeks pre-surge | 14 | 23 | 0.620 | 0.447 | 0.000565 | 0.00282 |
| 4-8 weeks pre-surge | 0-4 weeks pre-surge | 23 | 28 | 0.447 | 0.412 | 0.481115 | 1.00000 |
| 0-4 weeks pre-surge | Surge | 28 | 54 | 0.412 | 0.358 | 0.001035 | 0.00518 |
| Surge | Post-surge | 54 | 25 | 0.358 | 0.456 | 0.000535 | 0.00267 |
| Post-surge | Uncategorized | 25 | 14 | 0.456 | 0.620 | 0.000985 | 0.00493 |

**Table S11.** CV adjacent comparisons of upstate (hospitalizations)

| Category 1 | Category 2 | $n_1$ (weeks) | $n_2$ (weeks) | Median 1 | Median 2 | p-value | Adjusted p |
|---|---|---|---|---|---|---|---|
| Uncategorized | 4-8 weeks pre-surge | 27 | 17 | 1.176 | 0.928 | 0.566808 | 1.00000 |
| 4-8 weeks pre-surge | 0-4 weeks pre-surge | 17 | 23 | 0.928 | 0.946 | 0.220995 | 1.00000 |
| 0-4 weeks pre-surge | Surge | 23 | 45 | 0.946 | 0.779 | 0.000980 | 0.00490 |
| Surge | Post-surge | 45 | 32 | 0.779 | 0.999 | 5.93e-07 | 2.97e-06 |
| Post-surge | Uncategorized | 32 | 27 | 0.999 | 1.176 | 0.000668 | 0.00334 |

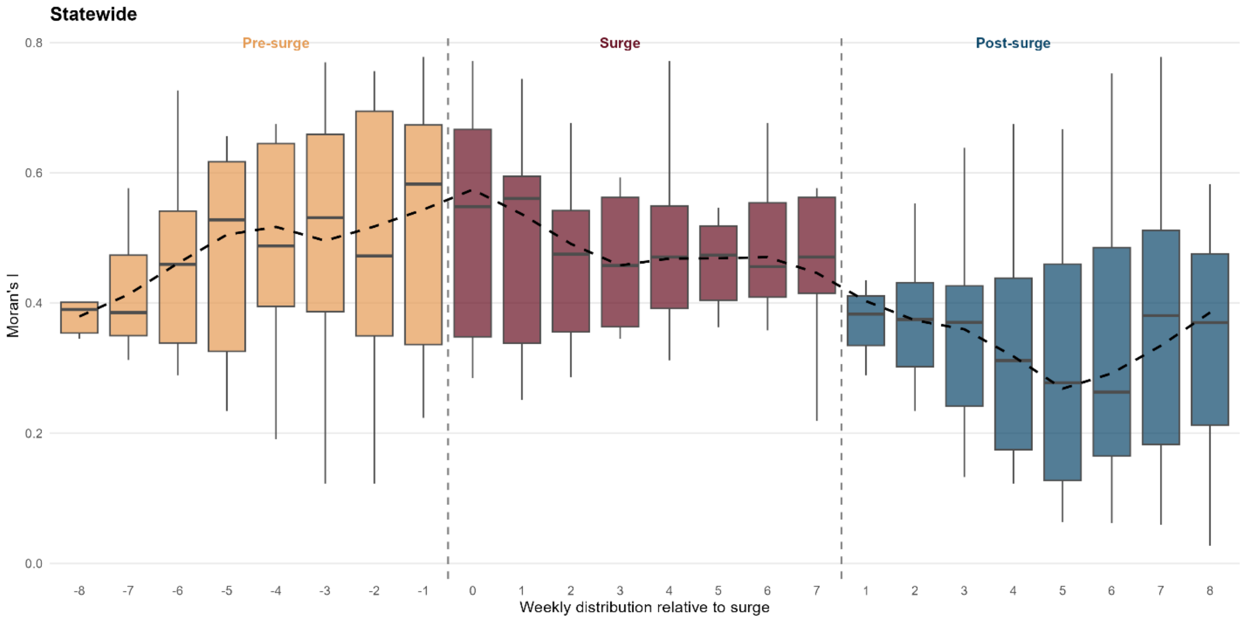


**Figure S38. Distribution of statewide global Moran's I by week relative to surge onset and termination for clinical cases.** Boxplots show the distribution of global Moran's I values at each weekly position, pooled across all surges and regions, with weeks aligned relative to surge onset (left dashed line) and surge termination (right dashed line). Surge duration varies across events; the surge panel displays weeks 0 through 7 as a representative range encompassing observed surge lengths. Pre-surge weeks show elevated CV values peaking just before surge onset, followed by a decline into the surge and post-surge period. The dashed black line traces the median CV across weekly positions. This pattern shows a more subtle bidirectional early warning signal as compared to CV.

#### Global Moran's I Kruskal-Wallis Pairwise Comparisons

These tables present pairwise Wilcoxon rank-sum test results that compare global Moran's I values for adjacent surge categories derived from clinical case data. The global Moran's I was calculated from county level clinical cases for the entire state. Tables for adjacent category comparison include the two surge categories being compared, the number of weekly observations in each group, the median CV for each group, the unadjusted Wilcoxon rank-sum test p-value, and the Bonferroni-corrected p-value adjusted for multiple comparisons. Green highlighting indicates adjusted $p < 0.05$; orange indicates $0.05 \leq$ adjusted $p < 0.10$.

**Table S12.** Global Moran's I Statistic statewide adjacent comparisons

| Category 1 | Category 2 | $n_1$ (weeks) | $n_2$ (weeks) | Median 1 | Median 2 | p-value | Adjusted p |
|---|---|---|---|---|---|---|---|
| Uncategorized | 4-8 weeks pre-surge | 23 | 19 | 0.2402 | 0.3995 | 9.23e-06 | 4.62e-05 |
| 4-8 weeks pre-surge | 0-4 weeks pre-surge | 19 | 26 | 0.3995 | 0.5195 | 0.249 | 1.000 |
| 0-4 weeks pre-surge | Surge | 26 | 45 | 0.5195 | 0.4217 | 0.151 | 0.756 |
| Surge | Post-surge | 45 | 24 | 0.4217 | 0.1749 | 1.41e-07 | 7.04e-07 |
| Post-surge | Uncategorized | 24 | 23 | 0.1749 | 0.2402 | 0.650 | 1.000 |
